# Lifecourse Genome-Wide Association Study Meta-Analysis Refines Understanding of the Critical Life Stages for the Influence of Adiposity on Breast Cancer Risk

**DOI:** 10.1101/2025.04.08.25325440

**Authors:** Grace M. Power, Laxmi Bhatta, Amanda Hughes, Carolina Medina-Gomez, Anne Richmond, Genevieve Leyden, Bethan Lloyd-Lewis, Eleanor Sanderson, Rebecca Richmond, Elizabeth C. Corfield, Daniel McCartney, Caroline Hayward, Irene Fontes Marques, Fernando Rivadeneira, Bjørn Olav Åsvold, Gibran Hemani, Janine F. Felix, Ben Brumpton, Alexandra Havdahl, George Davey Smith

## Abstract

Previous evidence suggests that higher prepubertal adiposity may protect against breast cancer risk; however, this protective effect does not appear to persist into later life. The specific age at which this effect diminishes remains unclear and has yet to be explored using causal inference methods.

This study examined the effect of body mass index (BMI) in nulliparous women during the early reproductive years on breast cancer risk. Using data from five large cohorts, we conducted genome-wide association studies (GWAS) on BMI from menarche to <40 years (N = 56,628), including three age sub-intervals: menarche to <20 years, 20 to <30 years, and 30 to <40 years. Results were meta-analysed, and consistency in genetic effects across age intervals was assessed. Two-sample Mendelian randomization (MR) within a lifecourse framework was applied to estimate the causal effect of genetically proxied BMI on overall breast cancer risk and seven subtypes using data from the Breast Cancer Association Consortium (N=up to 247,173).

Heterogeneity in genetic effects on BMI across early adulthood was observed, with 9 of the 45 discovery variants showing significant variation (Qhet < 0.05). Genome-wide genetic correlations suggested that BMI in early adulthood may be influenced by partially distinct genetic factors compared to other life stages (rG = 0.76 with prepubertal body size; rG = 0.85 with later-life body size). Univariable MR analyses provided strong evidence that higher genetically predicted BMI between menarche and <40 years reduced overall breast cancer risk as well as most subtypes except HER2-enriched breast cancer. These effects persisted after adjusting for later-life body size but attenuated when prepubertal body size was included in multivariable MR models.

Our findings suggest that while higher BMI in early adulthood is associated with reduced breast cancer risk, this effect may in part be attributable to adiposity accrued before puberty. These results refine our understanding of the timing of adiposity’s protective influence on breast cancer and highlight earlier life stages as critical windows for risk modulation.

**Teaser:** Improving knowledge of adiposity’s genetic architecture across the lifecourse refines insights into its role in breast cancer

## Introduction

Breast cancer is the most commonly diagnosed cancer in women worldwide, with over 2.3 million new cases and 685,000 deaths reported in 2020 (1). Nearly 99% of women diagnosed at the earliest stage survive for 5 years or more, compared to approximately 32% of those diagnosed at the most advanced stage (2). This underscores the importance of improving early detection and highlights the need to identify preventative strategies as key public health objectives.

Breast tissue is particularly vulnerable to exposures between menarche and first full-term pregnancy – a critical window of susceptibility (3). After menarche, the breast epithelium undergoes rapid cellular proliferation leading to an increased risk of acquiring mutation during puberty and until the terminal ductal differentiation that accompanies the first full-term pregnancy (4, 5). Evidence suggests that certain exposures such as physical activity (3), may alleviate the risk of breast cancer, while others, such as alcohol consumption (6), a known breast carcinogen (7), may exacerbate it, by acting during this period of tumorigenic susceptibility. By investigating whether known factors influencing breast cancer risk also exert effects during this timeframe, we may be better able to identify strategies for more effective breast cancer risk detection and potential avenues for prevention. Focusing on this period may additionally enable a more precise assessment by minimising the confounding effects of physiological changes associated with first birth.

Mendelian randomization (MR) is a technique that exploits the quasi-random distribution of genetic variants from parents to offspring, independent of the influence from other traits. Under specific assumptions, MR aims to estimate causal effects by reducing susceptibility to confounding factors, including confounding by undiagnosed existing disease and disease processes (reverse causation) (8, 9). Recent developments in lifecourse MR methodology include multivariable MR (MVMR) (10–12). This approach enables the direct estimation of the effects of an exposure measured at a specific life stage, controlling for the same exposure measured at another life stage, on later life outcomes. Previous MVMR analyses suggest that a larger prepubertal body size, employed as a proxy for BMI, is protective against breast cancer (13–15). This protective effect persists even after controlling for later life adult body size, which has a negligible effect on breast cancer risk when prepubertal body size is controlled for. These findings align with conventional analyses (16–18) and have been further supported through a proxy-genotype MR approach (19).

Although a larger body size in childhood has been shown to reduce breast cancer risk, it also leads to earlier menarche, a known risk factor for breast cancer (13). This dual effect suggests that the protective direct effect of prepubertal body size on breast cancer could be larger than its total observed effect, as it may be partially attenuated by an increased risk mediated through earlier menarche. This interplay highlights two potentially independent pathways that may interact in complex ways. Despite convincing evidence for the protective effects of prepubertal body size on breast cancer risk, the time period at which this protective effect diminishes is yet to be determined using causal inference methods.

We aimed to estimate the effect of higher BMI between menarche and first full-term pregnancy—or throughout early adulthood (<40 years) in individuals who remain nulliparous— on breast cancer risk later in life. This previously understudied period represents a critical window of susceptibility to exposures influencing breast cancer risk. We assess these effects on overall breast cancer and seven subtypes, including ER status (ER+/ER-) and five molecular subtypes: Luminal A, Luminal B1 (HER2+), Luminal B2 (HER2-), HER2-enriched, and triple-negative breast cancer using data from the Breast Cancer Association Consortium (BCAC) (20, 21), with the aim of exploring whether the effects of a higher BMI during this period differ in relation to menopausal status through associations with these breast cancer subtypes.

## Materials and methods

This study adheres to the Strengthening The Reporting of Observational Studies in Epidemiology Using Mendelian Randomisation (STROBE-MR) guidelines (Table S1).

### Data sources and study design

#### Inclusion criteria and participating cohorts

We analysed five large population-based prospective cohorts with genomic and phenotypic BMI data from nulliparous women between menarche and <40 years. These included the Avon Longitudinal Study of Parents and Children (ALSPAC), the Trøndelag Health Study (HUNT), the Norwegian Mother, Father and Child Cohort Study (MoBa), Generation R, and Generation Scotland. Participant selection is shown in Figure 1.

**Figure 1.**
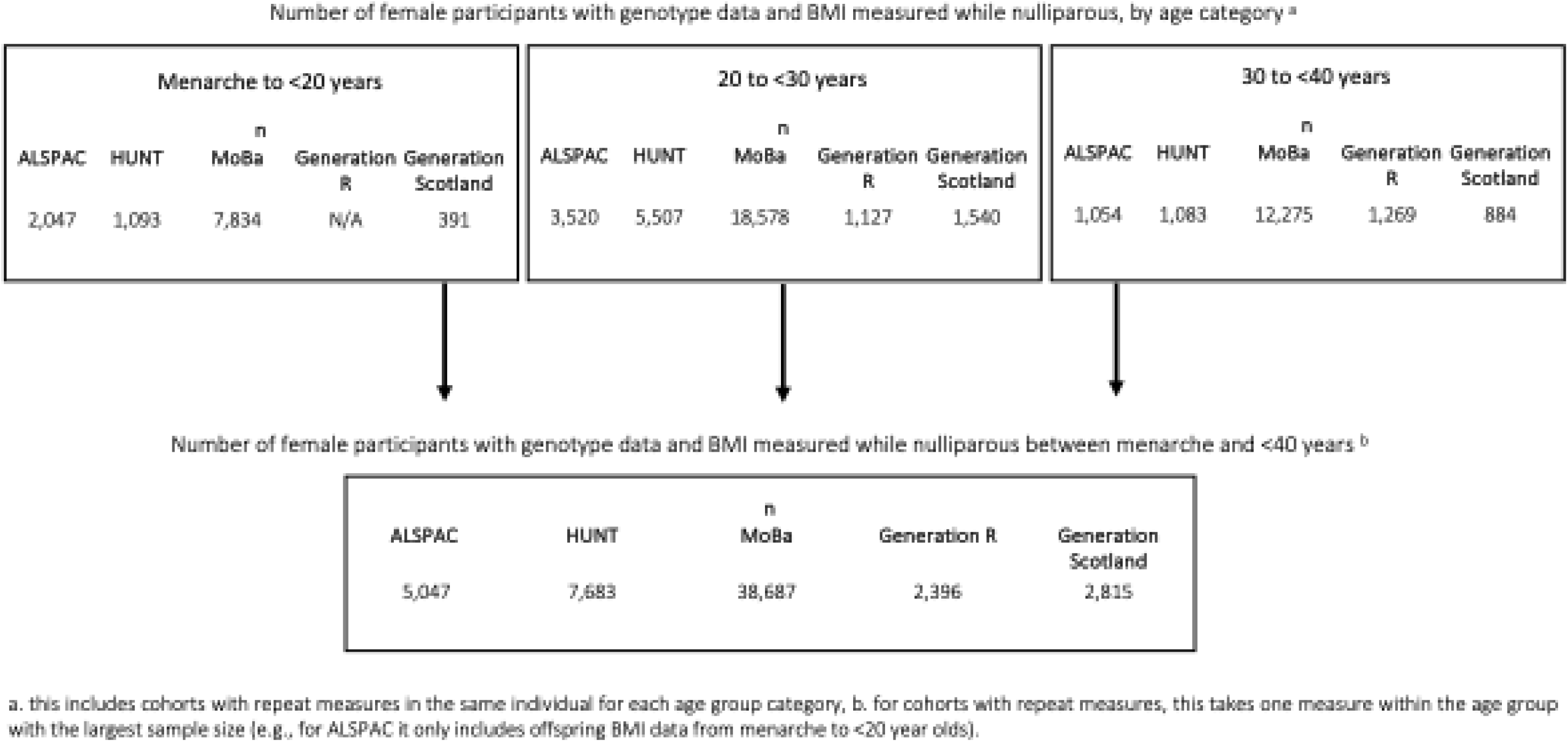
Overview of study participants used for this analysis

ALSPAC. ALSPAC is a birth cohort which enrolled 14,541 pregnant women living in Avon, England with expected dates of delivery between April 1, 1991, and December 31, 1992 (the estimated recruitment rate of those eligible was 80%) (22–24). Further enrolments after 1998 resulted in a baseline sample of 14,901 children alive at one year of age. Some study data were collected and managed using Research Electronic Data Capture (REDCap) electronic data capture tools hosted at the University of Bristol (25). Please note that the study website contains details of all the data that is available through a fully searchable data dictionary and variable search tool: http://www.bristol.ac.uk/alspac/researchers/our-data/. We used data collected on women prior to pregnancy and their daughters in the 17 and 24-year follow up groups that had information on genotype, menarche, BMI and age. We included 3,125 ALSPAC mothers and 3,496 daughters from the offspring generation in the age-stratified analysis. In the analysis focused on individuals between menarche and first birth (if pregnancy occurred within this timeframe), we included 3,125 ALSPAC mothers and 1,922 daughters.

HUNT. HUNT is a cohort study of the adult population in Trøndelag County, Norway, and comprises ∼230,000 individuals aged ≥20 years (26, 27). Individuals were recruited in four surveys from 1984–2019, of which the HUNT2 and HUNT3 surveys were included in the present study. Invitee participation rates were 69% and 54%, respectively. Data on perinatal outcomes are available via linkage to the Medical Birth Registry of Norway (MBRN) (28) for individuals born after 1967. The full recruited sample of mothers was 64,098. A total of 69,716 offspring were available in HUNT, with genotype and measured phenotype information, of which 36,938 were female (29, 30). A total of 7,683 individuals were used in this study.

MoBa. MoBa is a population-based pregnancy cohort study conducted by the Norwegian Institute of Public Health (31). Participants were recruited from all over Norway from 1999 to 2008, with 41% of all pregnant women invited consenting to participate. Blood samples were obtained from both parents during pregnancy and from mothers and children (umbilical cord) at birth (32). The first child was born in October 1999 and the last in July 2009. The cohort includes approximately 114,500 children, 95,200 mothers, and 75,200 fathers. Additionally, MoBa was linked to data from the Medical Birth Registry (MBRN), a national health registry containing information about all births in Norway from 1967 onwards (28). This analysis used a total of 67,690 MoBa mothers and 7,216 daughters from the offspring generation with information on genotype, menarche, BMI, and age.

Generation R. The Generation R Study is a population-based prospective cohort from fetal life onwards in Rotterdam, the Netherlands (33). Women with an expected delivery date between April 2002 and January 2006 living in Rotterdam were eligible to enrol the study, and written informed consent was obtained from all participants. A total 9,778 mothers, who gave birth to 9,749 live-born children, enrolled in the study. Genotype data were available for a total of 7,236 mothers, of which 3,450 were first-time mothers of singleton children and had information on pre-pregnancy BMI (34). Maternal age and pre-pregnancy weight were reported by the mother and height was measured at intake. A total of 2,479 women contributed to this GWAS.

Generation Scotland. Generation Scotland is a population-based cohort with ∼20,000 participants from across Scotland, aged between 18 and 98 years. During 2006 to 2010, potential participants (aged 35–65 years) were identified and invited to join at random from collaborating general medical practices in Scotland. Participants were asked to identify ≥1 first-degree relatives aged ≥18 years who would also be able to participate. Subsequently, participants attended a staffed research clinic, completed a health questionnaire, and had physical and clinical characteristics measured according to a standardized protocol. Fasting blood and urine samples were collected, according to standard operating procedures. The full recruited sample of female participants was 14,152, of which 3,059 were included in this study.

#### Genotyping, quality control and imputation in each cohort

Genotyping, quality control and imputation for each cohort are described in detail in Supplementary Text and elsewhere (30, 34–36). Briefly, participants were genotyped using genome-wide SNP arrays, and imputation was performed using standard reference panels following cohort-specific quality control (QC) protocols. ALSPAC mothers and children were genotyped using Illumina Human660K and HumanHap550 arrays, respectively, with imputation via Haplotype Reference Consortium (HRC) v1.1. HUNT samples were genotyped using three Illumina HumanCoreExome arrays, with imputation performed using Minimac3 and a customized HRC v1.1 reference panel (30, 35). MoBa sampels were genotyped across 24 batches with varying selection criteria, centers, and arrays. The European Genome-Phenome Archive HRC v1.1 (Study ID EGAS00001001710) was used for phasing and imputation (31, 36). Generation R mothers were genotyped using Illumina GSA-MD 2.0 and 3.0 arrays, with imputation against the 1000 Genomes Phase 3 v5 reference panel (33, 34). Generation Scotland samples were genotyped using the Illumina HumanOmniExpressExome-8v1 chip, with imputation via HRC.r1-1 on the Sanger Imputation Server.

### Statistical methods

#### Lifestage-stratified genome-wide association studies (GWAS) and meta-analysis

We aimed to establish lifestage-specific genetic effect profiles for BMI. This was to (i) assess the consistency of genetic variant effects on BMI across different age groups (including prepubertal and later-life adulthood, where body size was used as a proxy for BMI based on previously derived genome-wide association studies (GWAS) from UK Biobank data (14)) and (ii) applying these profiles in downstream lifecourse MR analyses. To achieve this, we conducted GWAS on lifestage-stratified BMI values across an age range that has not been previously investigated. Specifically, separate GWAS analyses were run on nulliparous women between menarche and <40 years. These analyses were further stratified into three age groups: <20 years, 20 to <30 years, and 30 to <40 years. For BMI measurements from mothers in birth cohorts, we extracted pre-pregnancy data from women who had not given birth previously. A complete-case approach was used, excluding individuals with missing values from the analysis. Due to the skewed distribution of BMI in the data, a natural log transformation was applied prior to running the GWAS analyses within each cohort. GWAS analyses were carried out independently by individual studies, each following their own quality control (QC) procedures (Supplementary Text and elsewhere (30, 34–36). All analyses were restricted post hoc to common genetic variants with a minor allele frequency (MAF) ≥ 0.01. Although mothers and daughters are related in several of the cohorts used, they were treated as independent samples in this study, with adjustments made to account for their relatedness..

We meta-analysed summary statistics from the five cohorts for each lifestage-specific GWAS separately, using a fixed effects model employed in METAL version 2020-05-05 (37). We carried out further standard quality control QC procedures (38) including tests for bias due to population structure (genomic control inflation factor [λ] and Linkage Disequilibrium Score Regression [LDSC] (39)). We also estimated SNP heritability and assessed genetic correlations between our meta-analysed GWAS and GWAS derived using UK Biobank data of prepubertal body size and adult body size (14). Analyses were performed using the LDSC software v1.0.0 (https://github.com/bulik/ldsc) employing default parameters. We used linkage disequilibrium (LD) scores from European participants of the 1000 genomes project that can be downloaded from the LDSC website. We generated Manhattan plots which contain horizontal lines drawn at –log10(1 × 10^−5^) for “suggestive associations” and –log10(5 × 10^−8^) for the ’genome-wide significant’ threshold and QQ plots (Figure S1-S4) (40).

We used LD clumping with an r^2^ threshold of 0.001 to select a set of independent instruments for BMI at each age group. This was performed using PLINK (41) and genotype data from European individuals from phase 3 v5 enrolled in the 1000 genomes project as a reference panel (42).

We visualised the age-specific effect trajectories of these SNPs at each age group (menarche to <20 years, 20 to <30 years and 30 to <40 years), with point size inversely proportional to the standard error of the effect estimate. We focused exclusively on SNPs derived from the age groups analysed in this study and did not include those from other studies due to differences in effect size scales. For example, body size measures for prepubertal and later adulthood stages, derived from UK Biobank data, were based on body size categories rather than BMI, making direct comparisons inconsistent (14).

#### Publicly available genome-wide association study (GWAS) data

We used publicly available breast cancer data from the Breast Cancer Association Consortium (BCAC) from 2017 (N= 228,951; ER+/ER- samples, extracted from OpenGWAS under IDs: ieu-a- 1127, ieu-a-1128) (20) and the latest release of BCAC from 2020 (N= 247,173; overall sample and five molecular subtypes: Luminal A, Luminal B1 (HER2+), Luminal B2 (HER2-), HER2- enriched, and triple-negative breast cancer (21) (details in Table S2). In the BCAC classification, Luminal B1 (HER2+) tumours are characterized by ER+, progesterone receptor positive (PR+), and HER2+ status, whereas HER2-enriched tumours are defined as HER2+, ER-, PR+ tumours. The cohort design and genotyping protocol details are described elsewhere (bcac.ccge.medschl.cam.ac.uk/bcac-groups/study-groups/, bcac.ccge.medschl.cam.ac.uk/bcacdata/). The study groups in the BCAC consortium do not include ALSPAC, HUNT, MoBa, Generation R, Generation Scotland or UK Biobank cohorts.

For comparative purposes, we utilised previously conducted GWASs of prepubertal and later life adult body size, used as a proxy for BMI, previously undertaken in the UK Biobank study on 246,511 females. These GWAS represent the largest available sample size for body size in these age groups. GWASs were adjusted for age and genotyping chip and genetic variants were selected based on P< 5 × 10^−8^ and r^2^ < 0.001 (14, 43, 44). UK Biobank data were collected between 2006 and 2010 on individuals aged between 40 and 69 years old at baseline, including data from clinical examinations, assays of biological samples, detailed information on self-reported health characteristics, and genome-wide genotyping, using a prospective cohort study design (44). The prepubertal body size measure utilised recall questionnaire data, involving responses from adult participants who were asked whether, compared to the average, they were ‘thinner’, ‘about average’ or ‘plumper’, when they were aged 10 years old. The adult body size variable was derived using clinically measured body mass index (BMI) data (mean age 56.5 years). It was then separated into a 3-tier variable using the same categories as the childhood body size measure for comparability; “thinner” (21.1 kg/m^2^-25 kg/m^2^), “about average” (25 kg/m^2^-31.7 kg/m^2^) and “plumper” (31.7 kg/m^2^-59.9 kg/m^2^) (14). These scores have been independently validated in three distinct cohorts, providing verification that these genetic instruments can reliably separate childhood and adult BMI (14, 45, 46).

To disentangle whether our results were influenced by age at menarche, we used publicly available data (Age when periods started (menarche)) from the UK Biobank cohort where female individuals were asked ‘How old were you when your periods started?’. This GWAS was conducted in 2014 (N=182,416, extracted from OpenGWAS under ID: ieu-a-1095)). To investigate the potential for collider bias resulting from restricting our analyses to nulliparous women we used publicly available parity data (Number of live births) from the UK Biobank cohort from 2018 (N= 250,782, extracted from OpenGWAS under ID: ukb-b-1209).

#### Lifecourse Mendelian randomization (MR) analysis

Our primary two-sample MR analyses used the inverse variance weighted (IVW) estimator, implemented in the TwoSampleMR R package (47). When genetic variants are used as instrumental variables in MR, the assumptions of instrumental variables must be met, i.e., the genetic variants used must (i) be strongly associated with the exposure of interest and relevant to the population studied, here pregnant women (“relevance”), (ii) not share common causes with the outcome (“independence”), and (iii) not affect the outcome other than through the exposure (“exclusion-restriction”) (48). We conducted sensitivity analyses, which relax the assumptions made about horizontal pleiotropy, including MR Egger regression (49), and the weighted median-based estimator (50). We ran IVW multivariable MR (MVMR), an extension of MR that employs multiple genetic variants associated with multiple measured risk factors, to calculate the direct and indirect effects of BMI between menarche and first birth on breast cancer outcomes. This analysis accounted for prepubertal and later-life adult body size separately (Figures 2A, B and C). These two additional time points were not included together in a single model due to the risk of weak instrument bias. Instrument strength was evaluated using the two-sample conditional *F*-statistic described elsewhere (51).

**Figure 2.**
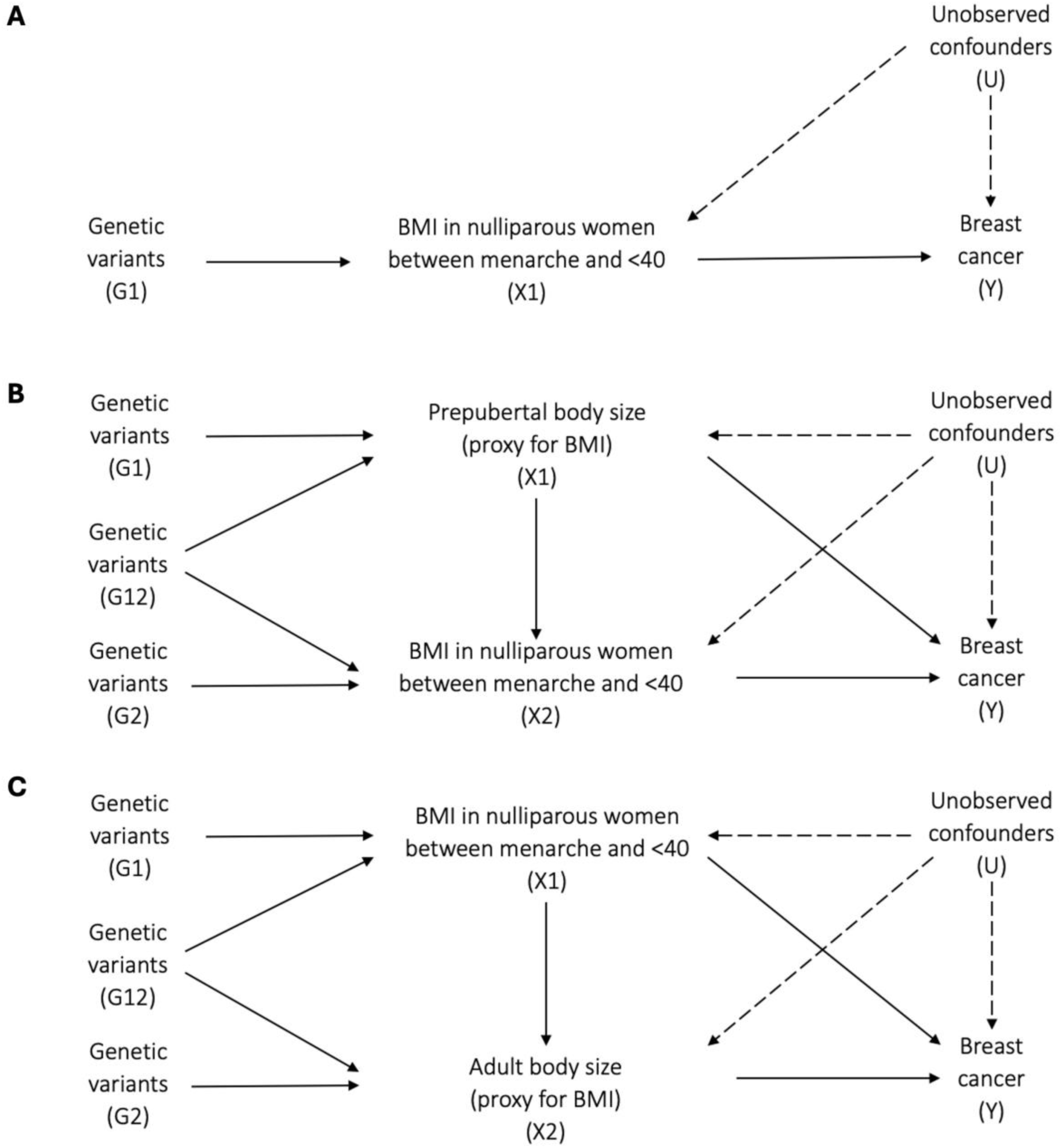
Directed acyclic graphs indicating three scenarios to explain the causal effect between BMI in nulliparous women between menarche and <40 years and breast cancer in later life. A. BMI in nulliparous women between menarche and <40 years has a total causal effect on breast cancer. B. BMI in nulliparous women between menarche and <40 years has a total causal effect on breast cancer, independent of prepubertal body size. C. BMI in nulliparous women between menarche and <40 years has a total causal effect on breast cancer, independent of adult body size.

#### Sensitivity analyses

To minimise pregnancy-related confounding, we restricted the BMI exposure GWAS to nulliparous women, enabling a clearer assessment of BMI’s direct influence across life stages. In contrast, the breast cancer outcome GWAS included both nulliparous and parous women. This discrepancy in selection mechanisms introduces potential selection bias in the genetic effect estimates for BMI but not for breast cancer. Specifically, conditioning on parity in the BMI GWAS may induce bias in the estimated SNP effects on BMI (𝛽^_𝐺𝑥_), whereas breast cancer estimates (𝛽^_𝐺𝑦_), remain unaffected as parity was not conditioned on in the outcome GWAS (Equation 1).

In this two-sample MR setting, the MR estimate is given by:

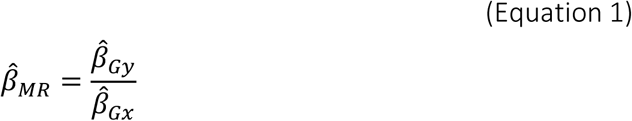

Where:

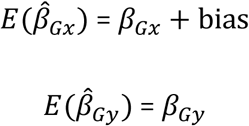

Since 𝛽^_𝐺𝑦_ is not subject to the same selection mechanism and there is no reason to assume that the selection-induced bias in 𝛽_𝐺𝑥_ is correlated with 𝛽^_𝐺𝑦_, a major spurious association in the MR analysis is unlikely. However, selection bias in the BMI GWAS could distort the MR estimate, with the direction and magnitude depending on whether and how BMI influences parity within the restricted sample. In an extreme scenario it could induce false positive associations with BMI that arise due to collider bias though this is unlikely at current sample sizes.

To empirically assess this, we performed univariable MR analyses estimating the effect of parity on BMI in nulliparous women across different life stages (menarche to <40 years, <20 years, 20 to <30 years, and 30 to <40 years). While parity itself cannot causally influence BMI in nulliparous women, genetic variants associated with parity may exhibit pleiotropic effects on BMI through shared metabolic and reproductive pathways. If these parity-associated variants also influence BMI within our restricted sample, it would suggest selection bias related to reproductive behaviour, implying that conditioning on parity in the BMI GWAS may have introduced collider bias.

In addition, a later onset of menarche has been linked to a reduced risk of breast cancer (52, 53). Childhood body size has been shown to accelerate the timing of menarche, whilst an earlier menarche increases the likelihood of increased body size in adulthood (54, 55). With these traits sharing a complex and interconnected relationship, we conduct sensitivity analyses estimating the effect of BMI in the lifestages analysed in nulliparous women on overall breast cancer accounting for age at menarche in MVMR analyses. Age at menarche is treated as a confounder, as it may influence both BMI and breast cancer risk through hormonal and metabolic pathways. Including it in the MVMR model ensures that observed BMI effects are not simply a reflection of differences in pubertal timing.

We conducted statistical analyses in R version 4.3.3 (56).

## Results

Measurements of body mass index (BMI) between menarche and age <40 years were available for 56,863 nulliparous women across five cohorts: the Avon Longitudinal Study of Parents and Children (ALSPAC), the Trøndelag Health Study (HUNT), the Norwegian Mother, Father and Child Cohort Study (MoBa), Generation R, and Generation Scotland. Corresponding phenotype sample sizes and cohort characteristics for age and BMI are presented in Table 1. Within the primary <40 year group, mean ages at measurement ranged from 23.9 years in ALSPAC to 29.4 years in Generation R, while mean BMI values varied between 22.8 kg/m² in ALSPAC and 24.5 kg/m² in Generation Scotland. To explore patterns across narrower age intervals within this broader period, we also conducted analyses stratified into three groups: menarche to <20 years, 20 to <30 years, and 30 to <40 years. Phenotypic sample sizes and summary measures of age and BMI for the full <40 group and each stratum are shown in Table 1.

**Table 1.** Descriptive statistics for nulliparous women between menarche and <40 years, menarche and <20 years, 20 and <30 years and 30 and <40 years, across all cohorts.

|  | Cohort | N | Age (years) |  | BMI (kg/m<sup>2</sup>) |  |
| --- | --- | --- | --- | --- | --- | --- |
|  |  |  | Mean (SD) | Median (Min-Max) | N | Mean (SD) |
| ALL (MENARCHE TO <40 YEARS) | ALSPAC | 5047 | 23.92 (6.01) | 24.00 (14.00-39.00) | 5047 | 23.92 (6.01) |
|  | HUNT | 7683 | 24.40 (5.04) | 23.7 (12.80-39.90) | 7683 | 24.40 (5.04) |
|  | MoBa | 38687 | 25.75 (6.68) | 27.00 (14.00-39.00) | 38687 | 25.75 (6.68) |
|  | Generation R | 2396 | 29.44 (4.71) | 30.13 (15.96-39.95) | 2396 | 29.44 (4.71) |
|  | Generation Scotland | 3059 | 26.29 (5.95) | 26.00 (14.00-39.00) | 3059 | 26.29 (5.95) |
| MENARCHE TO <20 YEARS | ALSPAC | 2047 | 17.78 (0.42) | 17.75 (14.00-19.33) | 2047 | 17.78 (0.42) |
|  | HUNT | 1093 | 16.90 (2.01) | 17.00 (12.80-19.90) | 1093 | 16.90 (2.01) |
|  | MoBa | 7834 | 14.74 (1.22) | 14.00 (14.00-19.00) | 7834 | 14.74 (1.22) |
|  | Generation R |  |  |  |  |  |
|  | Generation Scotland | 430 | 18.15 (0.94) | 18.00 (14.00-19.00) | 430 | 18.15 (0.94) |
| 20 TO <30 YEARS | ALSPAC | 3520 | 24.91 (2.12) | 24.00 (20.00-29.00) | 3520 | 24.91 (2.12) |
|  | HUNT | 5507 | 24.18 (2.69) | 23.70 (20.00-29.90) | 5507 | 24.18 (2.69) |
|  | MoBa | 18578 | 25.86 (2.51) | 26.00 (20.00-29.00) | 18578 | 25.86 (2.51) |
|  | Generation R | 1127 | 26.08 (2.86) | 26.62 (20.01 - 29.99) | 1127 | 26.08 (2.86) |
|  | Generation Scotland | 1745 | 24.41 (2.81) | 24.00 (20.00-29.00) | 1745 | 24.41 (2.81) |
| 30 TO <40 YEARS | ALSPAC | 1054 | 32.65 (2.45) | 32.00 (30.00-39.00) | 1054 | 32.65 (2.45) |
|  | HUNT | 1083 | 33.09 (2.60) | 32.30 (30.00-39.90) | 1083 | 33.09 (2.60) |
|  | MoBa | 12275 | 32.61 (2.38) | 32.00 (30.00-39.00) | 12275 | 32.61 (2.38) |
|  | Generation R | 1269 | 33.12 (2.25) | 32.76 (30.00 - 39.95) | 1269 | 33.12 (2.25) |
|  | Generation Scotland | 884 | 33.98 (2.85) | 34.00 (30.00-39.00) | 884 | 33.98 (2.85) |

### Lifestage-stratified genome-wide association studies (GWAS) and meta-analysis

Genome-wide association analyses (GWAS) for BMI were conducted in up to 56,628 nulliparous women with measures between menarche and age <40 years, and results were meta-analysed across cohorts. Stratified meta-analyses for the three life stages – menarche to <20 years, 20 to <30 years, and 30 to <40 years – included 11,365, 30,272 and 16,565, respectively (Figure 1). For cohorts with repeat measures within one life stage (e.g., menarche to <40 years), we retained the time point with the largest sample size (e.g., for ALSPAC, only BMI data from participants <20 years were included). Considering results across all GWAS time windows (menarche to <20, 20 to <30, 30 to <40, and the combined menarche to <40), we identified a total of 45 independent variants.

Overall, 31, 3, 12 and 5 independent genetic variants were associated with BMI in women between menarche to <40 years, menarche to <20 years, 20 to <30 years and 30 to <40 years prior to giving birth, respectively, based on conventional genome-wide corrections (i.e., P<5×10^−08^; Table S3).

Cochran’s Q statistic was used to evaluate heterogeneity among the 31 instrumental variables across the five cohort studies included in the primary analysis. To account for multiple comparisons, a Bonferroni correction was applied. After adjustment, none of the SNPs exhibited evidence of heterogeneity (*Q_bonf_*<0.05), suggesting consistent effect estimates across studies. This reinforces their validity in downstream MR analyses, reducing concerns that study-level differences may confound associations with the outcome.

We visualised the age-specific effect trajectories of these SNPs at each age group (menarche to <20 years, 20 to <30 years and 30 to <40 years), with point size inversely proportional to the standard error of the effect estimate (Figure S5). This revealed two notable patterns of heterogeneity: (i) SNP effects in the 20 to <30-year age group differ markedly from those in the menarche to <20-year and 30 to <40-year age groups, and (ii) some SNPs show a reversal in effect after age 30. Among 45 independent variants, nine showed substantial heterogeneity across the three age groups (*Q_het_*<0.05) (Figure 3). Given that the proportion of individuals from each cohort is comparable between the 20 to <30-year and 30 to <40-year age groups, the swing in effect estimates observed in the 20 to <30-year group is unlikely to be driven by a single cohort. These distinct patterns underscore age-specific heterogeneity that would likely be overlooked by linear meta-regression, highlighting the complexity of SNP effects on BMI over time.

**Figure 3.**
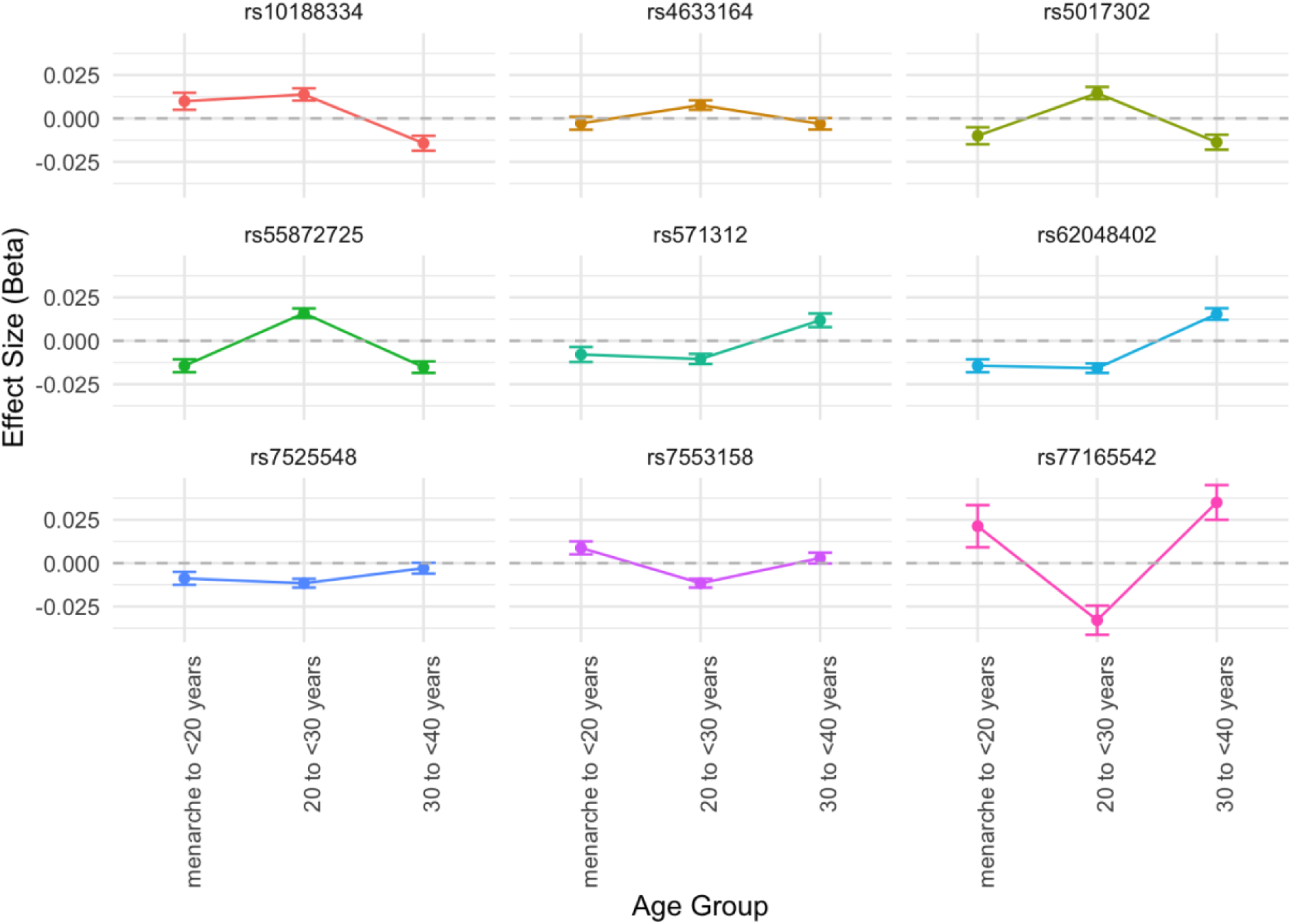
Plot illustrating the age-specific effect trajectories of individual SNPs with significant heterogeneity (QFDR < 0.05) on BMI across three defined age groups: menarche to <20 years, 20 to <30 years, and 30 to <40 years, in nulliparous women.

*Each plot represents a SNP exhibiting significant heterogeneity (QFDR < 0.05), illustrating its effect size (beta) across age intervals. Error bars denote 95% confidence intervals*.

### Heritability, LD score regression and genetic correlation analysis

We quantified the proportion of variance of the four meta-analyses undertaken on BMI in nulliparous women at different ages of European ancestry that can be explained by our SNP sets using Linkage Disequilibrium (LD) Score regression (57). The GWAS summary statistic SNP-based heritability ranged from 0.11 (SE = 0.01) for BMI in nulliparous women between 30 and <40 year olds, to 0.23 (SE = 0.02) for BMI in nulliparous women between 20 to <30 year olds (Table 2). These results indicate that genetic factors play an important role in BMI, particularly in the younger age groups. The number of GWAS hits aligns with expectations, closely tracking with heritability (ℎ^2^) and sample size, suggesting consistent polygenicity across age groups.

**Table 2.** Summary of meta-analysed genome-wide association results by age group categories.

| Metric | Age group category |  |  |  |
| --- | --- | --- | --- | --- |
|  | BMI in nulliparous women between menarche and <40 years | BMI in nulliparous women between menarche and <20 years | BMI in nulliparous women between 20 and <30 years | BMI in nulliparous women between 30 and <40 years |
|  | Estimate (SE) | Estimate (SE) | Estimate (SE) | Estimate (SE) |
| Heritability (Observed Scale $h^2$) | 0.19 (0.01) | 0.21 (0.03) | 0.23 (0.02) | 0.11 (0.01) |
| Genomic Inflation Factor (Lambda GC) | 1.27 | 1.07 | 1.17 | 1.09 |
| LD Score Regression Intercept | 1.01 (0.02) | 1.01 (0.01) | 1.00 (0.01) | 1.01 (0.01) |
| Proportion of Inflation (Ratio) | 0.03 (0.02) | 0.13 (0.08) | 0.01 (0.04) | 0.07 (0.06) |

The genetic correlations (*r_G_*) observed between prepubertal body size (used as a proxy for BMI) in females and BMI at different stages in nulliparous women from menarche to <40 years reveal consistently strong associations, as expected when investigating a similar phenotype (BMI and body size) across different life stages. The *r_G_* of 0.76 (SE = 0.02, P = 2.13x10^-223^) between prepubertal body size in females and BMI in nulliparous women <40 years indicates a high degree of shared genetic influence. This trend was mirrored at more granular age intervals, with *r_G_* values of 0.88, 0.74, and 0.71 for BMI in nulliparous women from menarche to <20, 20 to <30, and 30 to <40 years, respectively (Figure 4). These results suggest prepubertal body size has a slightly stronger genetic correlation in younger age groups, particularly for BMI measured before age 20. Similarly, body size (used as a proxy for BMI) in later life adult women shows strong genetic overlap with BMI measured in nulliparous women at various age periods, with *r_G_*values ranging from 0.68 (for BMI in nulliparous women from menarche to <20 years) to 0.95 (for BMI in nulliparous women aged 30 to <40 years). The consistently high *r_G_* values across all comparisons, particularly the near-perfect correlation between body size measured in later life adult women and BMI in nulliparous women aged 30 to <40 (*r_G_* = 0.95, SE = 0.05), may reflect an enduring genetic influence of BMI from adolescence into later reproductive years (Figure 4; Table S4). To generate the genetic correlation heatmap, we calculated the *r_G_* for measures with fully overlapping sample sizes. This overlap led to inflation of the *r_G_*estimate to be slightly greater than 1, and we capped the *r_G_* at a maximum value of 1.

**Figure 4.**
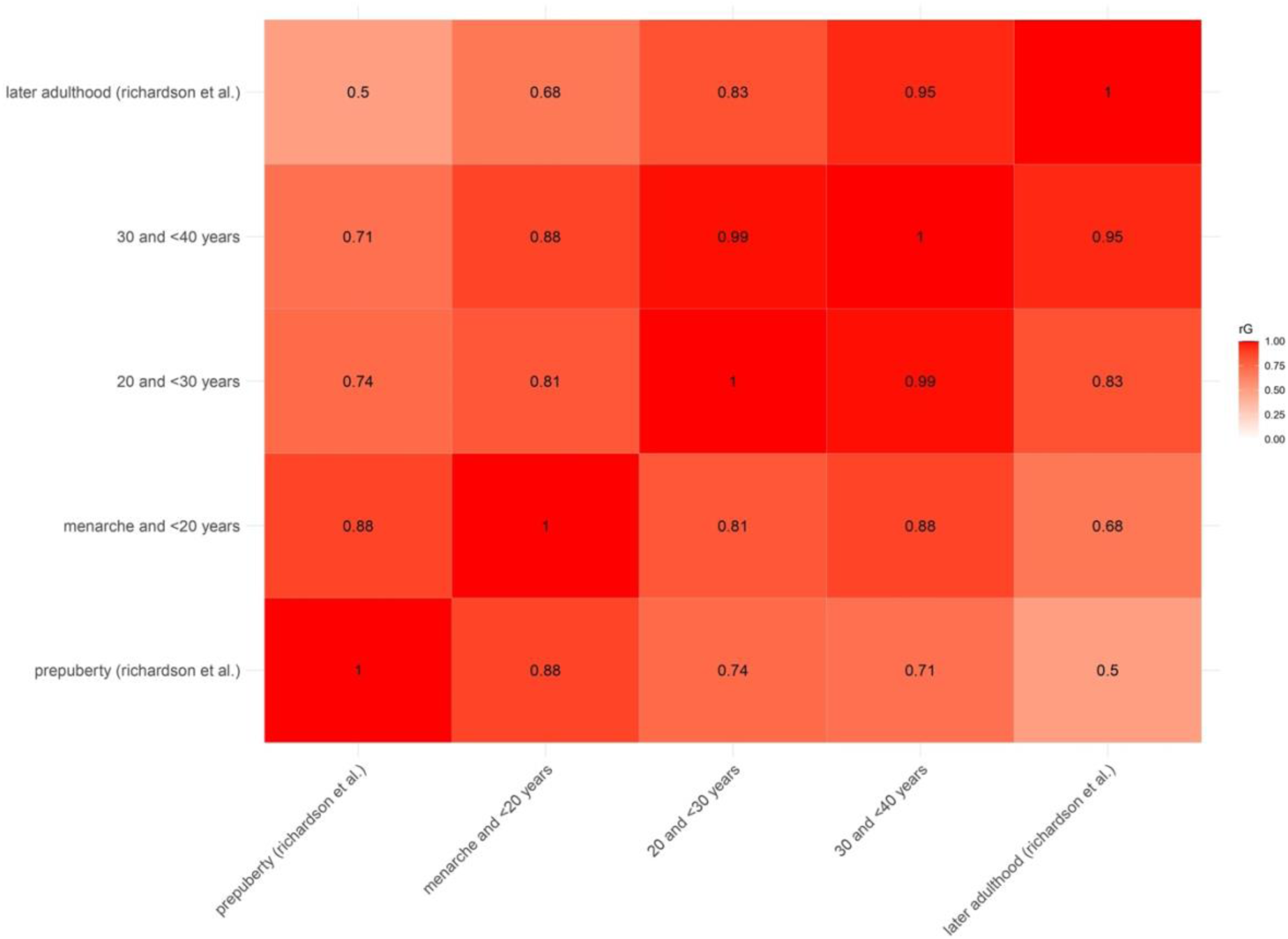
Genetic correlation heatmap for body mass index (BMI) and body size (used as a proxy for BMI) across various age groups in females, spanning from prepuberty through menarche and first birth to later adulthood.

### Lifecourse Mendelian randomization (MR) analysis

Univariable MR analyses using the 31 identified SNPs indicated strong evidence that higher genetically predicted BMI in nulliparous women between menarche and <40 years reduced the risk of overall breast cancer (IVW odds ratio (OR), 95% CI: 0.76, 0.67 to 0.86, P=1.27x10^-5^) (Figure 5; Table S5). Higher genetically predicted BMI in nulliparous women between menarche and <40 years additionally reduced the risk of most breast cancer subtypes, with the exception of the HER-2 enriched subtype. In multivariable MR accounting for adult body size, there remained evidence that higher genetically predicted BMI in nulliparous women between menarche and <40 years reduced the risk of overall breast cancer and most breast cancer subtypes, with the exception of the HER-2 enriched subtype. Adjustment for prepubertal body size led to a marked, though not complete, attenuation of these effects, consistent with part of the protection being shared with childhood adiposity.

**Figure 5.**
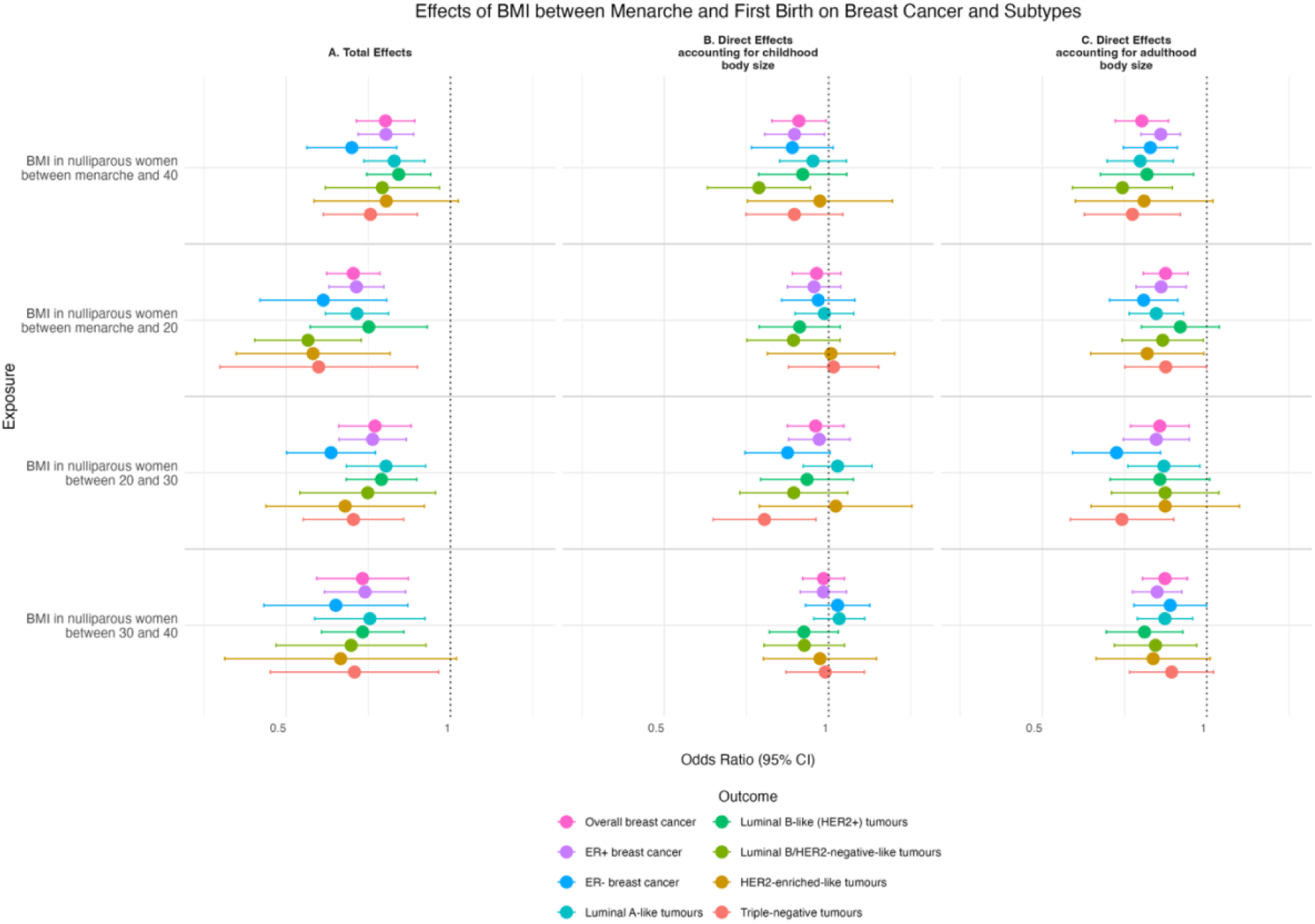
Univariable and multivariable Mendelian randomization (MR) for BMI between menarche and first birth on breast cancer (overall and subtype samples). A. The total effects (univariable MR). B. The direct effect accounting for childhood body size, and C. The direct effect accounting for adulthood body size (multivariable MR (MVMR)). The plots present the odds ratio of breast cancer per standard deviation (SD) increase in natural log-transformed BMI. Error bars indicate 95% confidence intervals around the point estimate from inverse- variance weighted (IVW) MR and IVW-MVMR analyses.

In the narrower life-stage periods obtained by partitioning the broader fertile-window measure, univariable MR analyses indicated evidence that higher genetically predicted BMI in nulliparous women between menarche to <20, 20 to <30, and 30 to <40 years reduced the risk of overall breast cancer and most subtypes, although the strength and precision of effects varied by subtype and life-stage period. After accounting for later life adult body size, protective effects were largely retained, with most estimates still supporting a reduction in risk. In contrast, adjusting for prepubertal body size removed much of the apparent protection in these narrower periods (Figure 5; Table S5). Consistent patterns of associations were observed using the weighted median method employed for robustness. In addition, results using the MR-Egger method did not provide evidence that horizontal pleiotropy was responsible for the estimates derived (Table S5).

Multivariable MR results should be interpreted with caution since conditional F-statistic was <10 for each of the BMI exposures included in these models, indicating potential for weak instrument bias (Table S5).

### Sensitivity analyses

Our analyses to investigate potential collider bias indicated very little evidence that genetically proxied parity associated with BMI in nulliparous women between menarche and <40 years, menarche and <20 years, and 20 and <30 years. There was some evidence that genetically proxied parity associated with lower BMI in nulliparous women between 30 and <40 years (Table S6). These results suggest collider bias due to selecting nulliparous women is likely to be of little concern within this study.

After adjusting for age at menarche, the estimated effect of BMI in nulliparous women between menarche and <40 years on overall breast cancer remains largely consistent with the UVMR analyses (Table S7). Age at menarche itself showed little direct effect on overall breast cancer or most subtypes once BMI was accounted for.

## Discussion

In this study, we undertook GWAS and MR analyses within a lifecourse framework to (i) assess the consistency of the genetic effects of BMI across different life stages, and (ii) investigate the effects of genetically proxied BMI in nulliparous women between menarche and <40 years on the risk of overall breast cancer and seven subtypes. This work builds on previous research to further the investigation into the influence of BMI during critical periods on breast cancer risk (13, 14, 58). Motivated by recent findings suggesting that a larger prepubertal body size, used as a proxy for BMI, may offer protection against breast cancer risk, while later life body size shows very little effect, we aimed to better understand the interval between puberty and later life. Specifically, we focused on the period between menarche and first full-term pregnancy, a crucial window of vulnerability for later life breast cancer development.

We observed considerable variation in genetic effects on BMI across different life stages for nine of the 45 discovery variants identified in this study. In addition, genome-wide cross- lifestage genetic correlations demonstrated variation, e.g., when comparing prepubertal body size and later-life body size with BMI from menarche to 40 years. In the univariable MR analyses, higher BMI in nulliparous women from menarche to <40 years was found to consistently reduce risk of breast cancer, including BMI measured in the separate age periods within this life stage (menarche to <20 years, 20 to <30, and 30 to <40 years), and risk of overall breast cancer and its subtypes. These protective effects largely remained upon accounting for later life adult body size. In contrast, adjusting for prepubertal body size in multivariable MR led to greater attenuation, indicating that part of the protective effect of BMI in early reproductive life may be explained by body size before puberty. The attenuation was more pronounced for the narrower life-stage periods than for the full fertile-window measure, reflecting both biological and methodological influences. Biologically, part of the apparent effect across all reproductive-age windows overlaps with childhood adiposity, so adjustment for prepubertal body size reduces the estimates. Methodologically, the narrower age-specific GWASs have substantially smaller sample sizes than the childhood GWAS. In multivariable MR, this imbalance can magnify the degree of attenuation by limiting power to detect effects independent of childhood size. By contrast, the broader reproductive-age measure is based on a larger GWAS sample and therefore shows less attenuation, despite adjustment for the same childhood body size data. Importantly, no such attenuation was observed when adjusting for later-life body size, even though similar sample size imbalances exist. This indicates that differential power alone may not fully explain the observed differences. Future methodological work is needed to examine how sample size disparities across life-stage GWASs influence estimates in multivariable MR and to develop strategies for accounting for this in lifecourse applications.

There is evidence suggesting two distinct pathways contributing to altered breast cancer risk during the prepubertal and pubertal periods: one associated with greater body size in earlier life and the other with earlier age at menarche (13). While a larger body size in childhood is protective, it may also accelerate age at menarche, which is itself a risk factor for breast cancer, suggesting that these pathways are not only independent but may also interact in complex ways. Furthermore, increased prepubertal body size has a decreasing effect on breast density, while age at menarche has been linked to higher breast density, which is another important risk factor for breast cancer (58, 59). This distinction between pathways highlights the need to examine potential mediating factors, which may help clarify the mechanisms at play. A deeper investigation into these separate but intersecting pathways could reveal novel insights into breast cancer risk.

The relationship between body size and breast cancer risk is complex. In conventional epidemiological settings, higher BMI has been linked to an increased risk of breast cancer in postmenopausal women and a decreased risk of breast cancer in premenopausal women (60–63). A plausible explanation includes the differing levels of oestrogen exposure between women experiencing overweight and normal-weight (64). Premenopausal women experiencing overweight tend to have longer anovulatory cycles, which reduces their exposure to ovarian hormones, potentially lowering their breast cancer risk. After menopause, fat tissue serves as another source of oestrogen production, which increases breast cancer risk among overweight women (65–68). Whilst our study did not specifically examine the relationship between overweight status and breast cancer pre and post menopause, effect estimates showed minimal variation between increased BMI and any of the breast cancer subtypes investigated in our study, with the exception of HER-2 enriched. Specifically, increased BMI in the 20 to <30-year category shifted from having a protective effect in univariable MR analyses to an increased risk of the HER-2 enriched subtype after accounting for body size later in life, though evidence supporting this effect is limited. There are likely complex interactions between body fat distribution, metabolic health, and HER-2 driven cell growth, potentially explaining why the HER-2 enriched subtype exhibited distinct patterns in response to higher BMI at different ages. This pattern is not observed in Luminal B1 (HER2+/ER+) tumours, suggesting that there may be a unique effect of HER-2 expression specifically in non-hormonally regulated tumours.

This study is an important and novel analysis with multiple strengths. It focuses on a specific age range where the protective effect of BMI on breast cancer risk begins to wane —an insight not previously achieved using causal inference methods. By examining this critical period between menarche and first birth, we enhance understanding on the potential interaction of nulliparity and BMI on breast cancer development. Focusing on this timeframe also allowed us to obtain an estimate that minimised the confounding effects introduced by the physiological changes associated with a first pregnancy. To strengthen the analysis, we integrated data from five large European longitudinal cohort studies, generating robust GWAS results on BMI. This approach allowed us to gain key insights into the consistency of BMI-related genetic effects across different stages of life. In addition, these data are not only useful for this particular study but also offer a valuable resource for future research into the effects of BMI in women at these life stages on other health outcomes.

This study has several limitations that should be considered when interpreting the findings. First, low conditional F-statistics were observed in the MVMR analysis, indicating potential for weak instrument bias. As a result, our MVMR findings should be interpreted with some caution. Second, while we combined data from five large European longitudinal cohorts, the sample size remains smaller (N=56,628) than that available for comparable measures of prepubertal and later-life body size derived using in UK Biobank data (N=246,511). This reduced sample size may limit statistical power for detecting smaller effect sizes. Third, used a prepubertal GWAS based on reported body size rather than measured BMI, as it offered a much larger sample size than any other GWAS of BMI at this life stage. However, this choice may distort MVMR analysis due to measurement error or differences in variance of effect sizes, and future work to precisely estimate childhood body size genetic effects are warranted. Fourth, there is the possibility of participant overlap between the HUNT and MoBa cohorts. While HUNT recruited individuals from Trøndelag County, Norway, MoBa had a national recruitment strategy. Some participants in HUNT may have also participated in MoBa, which could introduce a degree of sample duplication. Lastly, as our analysis was restricted to individuals of European ancestry, the generalisability of our findings to other populations is limited. Further research is needed to confirm these findings in more diverse populations.

## Conclusion

This study offers important insights into the genetic influences on BMI across different life stages and its causal relationship with breast cancer risk, focusing on the period between menarche and under 40 years in nulliparous women as a key window of susceptibility. While higher BMI in this interval appeared strongly protective against breast cancer in univariable analyses, the effect substantially, though not entirely, attenuated after accounting for childhood body size, used as a proxy for BMI. This pattern suggests that the protection may arise from the combined influence of greater adiposity in both childhood and early adulthood. These results have important implications for breast cancer prevention, underscoring earlier life stages as critical periods for potential interventions.

## Data Availability

Data access for the Avon Longitudinal Study of Parents and Children (ALSPAC) operates via a managed open access system. Approved proposals are reviewed by the ALSPAC Executive Committee. Full details are provided in the ALSPAC Data Management Plan (www.bristol.ac.uk/alspac/researchers/data-access/documents/alspac-data-management-plan.pdf). Data from the Norwegian Mother, Father and Child (MoBa) Cohort Study is managed by the Norwegian Institute of Public Health. Access is provided upon approval from the Regional Committees for Medical and Health Research Ethics (REC), compliance with GDPR, and data owner approval. Participant consent does not allow individual-level data storage in repositories or journals. Researchers seeking access for replication must apply via www.helsedata.no. To request access to data from the Trondelag Health Study (HUNT), researchers affiliated with Norwegian research institutes can apply for the use of HUNT data and biological samples, subject to approval by the Regional Committee for Medical and Health Research Ethics. Researchers from other countries may also apply if collaborating with a Norwegian Principal Investigator. All applications are reviewed by the HUNT Data Access Committee, and successful applicants are required to enter into data access and/or material transfer agreements. Detailed information on the application process, ethical requirements, and available datasets can be found on the HUNT website (www.ntnu.edu/hunt/data). Data from Generation Scotland are available on application to an independent access committee. More information can be found on the Generation Scotland website (www.generationscotland.org). Data from the Generation R Study are available upon reasonable request to the director of the Generation R Study, subject to local, national and European rules and regulations. All genetic instruments derived in this study are in the supplementary tables. Genome-wide association study summary statistics will be made available on a public repository upon publication. Code provided to collaborators for conducting the GWAS of BMI between menarche and first birth is available at: https://github.com/gracemarionpower/Collaboration_MA_BMI_M2FB. None of the material has been published or is under consideration for publication elsewhere.

## Acknowledgements.

We are extremely grateful to all the families who took part in this study, the midwives for their help in recruiting them, and the whole ALSPAC team, which includes interviewers, computer and laboratory technicians, clerical workers, research scientists, volunteers, managers, receptionists, and nurses. The authors would like to thank the research participants of the HUNT study which is a collaboration between HUNT Research Centre (Faculty of Medicine and Health Sciences, NTNU Norwegian University of Science and Technology), Trøndelag County Council, Central Norway Regional Health Authority, and the Norwegian Institute of Public Health. The HUNT genotype quality control and imputation has been conducted by the K.G. Jebsen Center for Genetic Epidemiology, Department of Public Health and Nursing, Faculty of Medicine and Health Sciences, NTNU. We are grateful to all the families who took part, the general practitioners and the Scottish School of Primary Care for their help in recruiting them, and the whole Generation Scotland team, which includes interviewers, computer and laboratory technicians, clerical workers, research scientists, volunteers, managers, receptionists, healthcare assistants and nurses. The Norwegian Mother, Father and Child Cohort Study is supported by the Norwegian Ministry of Health and Care Services and the Ministry of Education and Research. We are grateful to all the participating families in Norway who take part in this on-going cohort study. We thank the Norwegian Institute of Public Health (NIPH) for generating genomic data as part of the HARVEST collaboration, supported by the Research Council of Norway (#229624). We also thank the NORMENT Centre for providing genotype data, funded by the Research Council of Norway (#223273), South East Norway Health Authorities and Stiftelsen Kristian Gerhard Jebsen, and conducted by deCODE Genetics. We further thank the Center for Diabetes Research, the University of Bergen for providing genotype data and performing quality control and imputation of the data funded by the ERC AdG project SELECTionPREDISPOSED, Stiftelsen Kristian Gerhard Jebsen, Trond Mohn Foundation, the Research Council of Norway, the Novo Nordisk Foundation, the University of Bergen, and the Western Norway Health Authorities. The MoBa analyses were performed on the TSD (Tjeneste for Sensitive Data) facilities, owned by the University of Oslo, operated and developed by the TSD service group at the University of Oslo, IT Department (USIT). The analyses were performed using resources provided by Sigma2 - the National Infrastructure for High-Performance Computing and Data Storage in Norway. The Generation R Study is conducted by Erasmus MC in close collaboration with the School of Law and Faculty of Social Sciences of the Erasmus University Rotterdam, the Municipal Health Service Rotterdam area, Rotterdam, the Rotterdam Homecare Foundation, Rotterdam, and the Stichting Trombosedienst & Artsenlaboratorium Rijnmond (STAR-MDC), Rotterdam. We gratefully acknowledge the contribution of children and parents, general practitioners, hospitals, midwives, and pharmacies in Rotterdam. The generation and management of GWAS genotype data for the Generation R Study was done at the Human Genomics Facility, HuGe-F, housed within the Laboratory for Population Genomics of the Department of Internal Medicine at Erasmus MC. Genetic Laboratory of the Department of Internal Medicine, Erasmus MC, The Netherlands. We thank Pascal Arp, Gaby van Dijk, Marijn Verkerk, Samuel Gathan, Dr. Linda Broer and Jard de Vries for their help in creating, managing and QCing the GWAS database.

## Research Ethics and Informed Consent

Informed consent was obtained from all participants and ethical approval was obtained from the Regional Committee for Medical and Health Research Ethics, Central Norway (REK Central application number 2018/2488) (HUNT), the ALSPAC Ethics and Law Committee and the Local Research Ethics Committees (ALSPAC), the Medical Ethical Committee of Erasmus MC, University Medical Center Rotterdam, approved the study (MEC 198.782/2001/31) (Generation R), and The Regional Committees for Medical and Health Research Ethics (REK application number 2016/1702) (MoBa). Written consent was obtained from all participants in Generation Scotland. All components of Generation Scotland received ethical approval from the NHS Tayside Committee on Medical Research Ethics (REC Reference Number: 05/S1401/89). Generation Scotland has also been granted Research Tissue Bank status by the East of Scotland Research Ethics Service (REC Reference Number: 20-ES-0021).

## Data and computing code availability

Data access for the Avon Longitudinal Study of Parents and Children (ALSPAC) operates via a managed open access system. Approved proposals are reviewed by the ALSPAC Executive Committee. Full details are provided in the ALSPAC Data Management Plan (www.bristol.ac.uk/alspac/researchers/data-access/documents/alspac-data-management-plan.pdf). Data from the Norwegian Mother, Father and Child (MoBa) Cohort Study is managed by the Norwegian Institute of Public Health. Access is provided upon approval from the Regional Committees for Medical and Health Research Ethics (REC), compliance with GDPR, and data owner approval. Participant consent does not allow individual-level data storage in repositories or journals. Researchers seeking access for replication must apply via www.helsedata.no. To request access to data from the Trøndelag Health Study (HUNT), researchers affiliated with Norwegian research institutes can apply for the use of HUNT data and biological samples, subject to approval by the Regional Committee for Medical and Health Research Ethics. Researchers from other countries may also apply if collaborating with a Norwegian Principal Investigator. All applications are reviewed by the HUNT Data Access Committee, and successful applicants are required to enter into data access and/or material transfer agreements. Detailed information on the application process, ethical requirements, and available datasets can be found on the HUNT website (www.ntnu.edu/hunt/data). Data from Generation Scotland are available on application to an independent access committee. More information can be found on the Generation Scotland website (www.generationscotland.org). Data from the Generation R Study are available upon reasonable request to the director of the Generation R Study, subject to local, national and European rules and regulations.

All genetic instruments derived in this study are in the supplementary tables. Genome-wide association study summary statistics will be made available on a public repository upon publication.

Code provided to collaborators for conducting the GWAS of BMI between menarche and first birth is available at: https://github.com/gracemarionpower/Collaboration_MA_BMI_M2FB.

None of the material has been published or is under consideration for publication elsewhere.

