## Supplementary Materials for "Lifecourse Genome-Wide Association Study Meta-Analysis Refines Understanding of the Critical Life Stages for the Influence of Adiposity on Breast Cancer Risk"

Science Advances
AAAS

### Supplementary Materials for

#### Lifecourse Genome-Wide Association Study Meta-Analysis Refines Understanding of the Critical Life Stages for the Influence of Adiposity on Breast Cancer Risk

Grace M. Power *et al.**

**This PDF file includes:**

Supplementary Text

Figs. S1 to S5

Tables S1 to S7

References (1 to 52)

*Supplementary Text. Further information on genotyping, quality control and imputation in each cohort*

ALSPAC mothers were genotyped using the Illumina Human660K quad single nucleotide polymorphism (SNP) chip, and ALSPAC children using the Illumina HumanHap550 quad genome-wide SNP genotyping platform (1). ALSPAC fathers were genotypes using the 1000 Genomes phase 1 panel (2). Genotype data for all groups were imputed using the Haplotype Reference Consortium v1.1 reference panel after applying QC filters that excluded SNPs with minor allele frequency (MAF) ≤1%, call rate ≤95%, and deviation from Hardy–Weinberg equilibrium (HWE). Samples were further excluded based on incorrect sex assignment, evidence of cryptic relatedness, and non-European ancestry.

HUNT samples were genotyped using one of three different Illumina HumanCoreExome arrays (HumanCoreExome12 v1.0, HumanCoreExome12 v1.1, and UM HUNT Biobank v1.0) (3, 4). Genomic positions, strand orientation, and reference alleles were determined by aligning probe sequences against the human genome (Genome Reference Consortium build 37 and revised Cambridge Reference Sequence of mitochondrial DNA; http://genome.ucsc.edu) using BLAT (5). Ancestry was inferred by projecting genotyped samples onto principal components derived from the Human Genome Diversity Project (HGDP) reference panel (938 unrelated individuals; downloaded from http://csg.sph.umich.edu/chaolong/LASER/) (6, 7) using PLINK v1.9039 (8). The resulting genotype data were phased using Eagle2 v2.340 (9). Imputation was performed on the samples of recent European ancestry using Minimac3 (v2.0.1, http://genome.sph.umich.edu/wiki/Minimac3)(10) with default settings (2.5 Mb reference based chunking with 500 kb windows) and a customized Haplotype Reference consortium release 1.1 (HRC v1.1) for autosomal variants and HRC v1.1 for chromosome X variants (11). SNPs with MAF <1% and call rate <95% were excluded, and samples were removed if they showed excess heterozygosity (±3 standard deviations from the mean) or were identified as ancestral outliers based on principal component analysis. Additionally, deviations from Hardy-Weinberg equilibrium ($p < 1\times10^{-6}$) led to SNP exclusion to maintain genotype quality. This process was explained previously (12).

MoBa samples were genotyped through several research projects, across 24 genotyping batches with varying selection criteria, genotyping centers, and genotyping arrays. Detailed information on batch selection criteria, genotyping, pre-imputation quality control (QC), phasing, imputation, and post-imputation QC are described in full elsewhere (13). The establishment of MoBa and initial data collection was based on a license from the Norwegian Data Protection Agency and approval from The Regional Committees for Medical and Health Research Ethics. The MoBa cohort is currently regulated by the Norwegian Health Registry Act.

Generation Scotland samples were genotyped using the Illumina HumanOmniExpressExome-8v1 chip, and genotype calling was performed with the Beadstudio-Gencall v3 algorithm. QC excluded SNPs with MAF <1%, call rate <98%, and deviation from HWE (p-value $<1\times10^{-6}$). Samples were excluded if they had a call rate <98%. Phasing was conducted using SHAPEIT2 with the duoHMM option, and imputation was performed using the Haplotype Reference Consortium (HRC.r1-1) reference panel via the Sanger Imputation Server. Post-imputation filtering removed SNPs with an imputation quality score <0.4, duplicate variants, and monomorphic SNPs.

Generation R mothers were genotyped using the Illumina GSA-MD 2.0 and 3.0 arrays (14). Genotype data were imputed against the 1000 Genomes Phase 3 v5 reference panel following QC filters that excluded SNPs with MAF <0.01%, call rate <99%, and deviation from HWE. Samples were further excluded for incorrect sex assignment, genetic duplicates, and non-European ancestry.

*Figure S1A. Manhattan plot displaying the results of the meta-analysed genome-wide association studies (GWAS) for BMI in nulliparous women between menarche and <40 years*

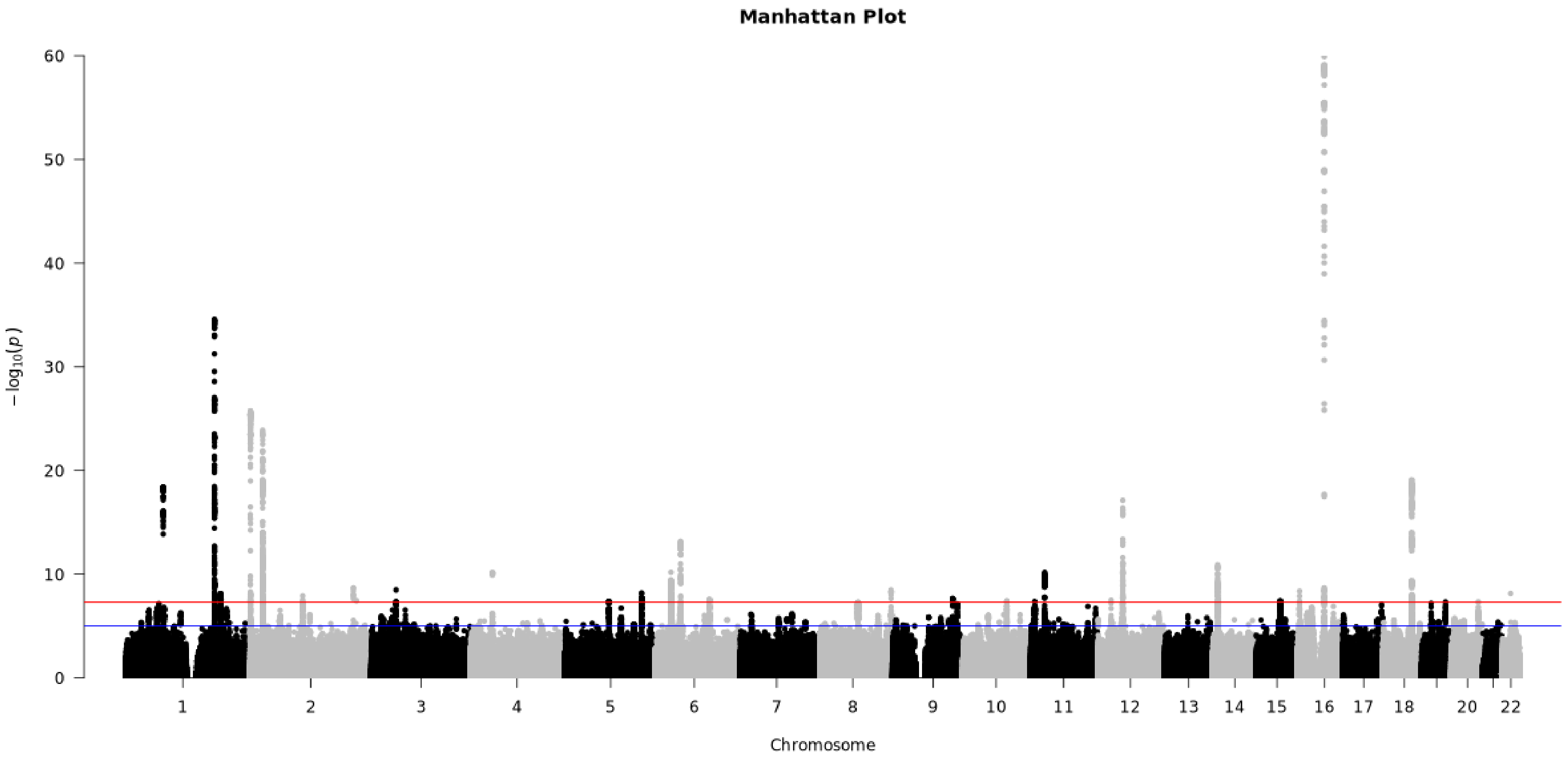

The x-axis represents the chromosomes, while the y-axis represents the -log10(p-values) of the SNPs. Horizontal lines are drawn at -log10(1 × $10^{-5}$) for "suggestive associations" in blue and -log10(5 × $10^{-8}$) for the "genome-wide significant" threshold in red.

*Figure S1B. QQ plot illustrating the observed vs. expected -log10(p-values) from the meta-analysed genome-wide association studies (GWAS) for BMI in nulliparous women between menarche and <40 years*

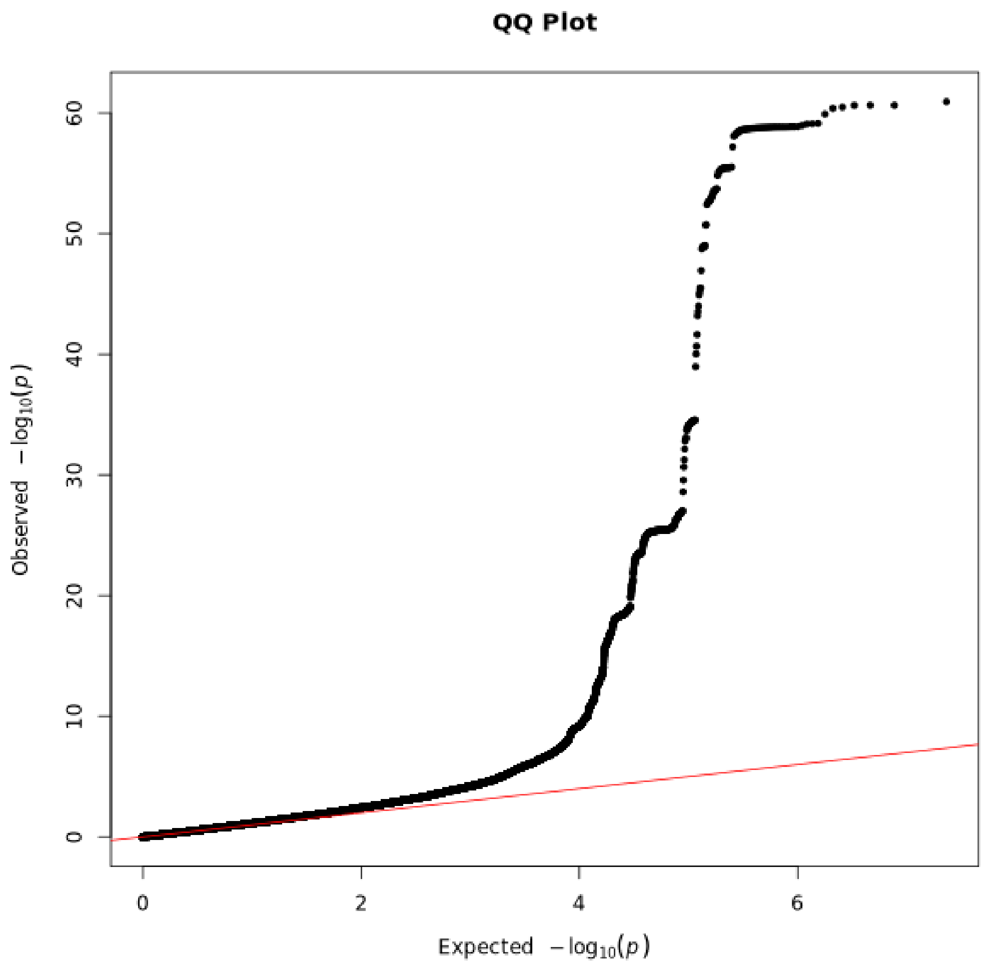

The red line represents the null hypothesis of no association.

*Figure S2A. Manhattan plot displaying the results of the meta-analysed genome-wide association studies (GWAS) for BMI in nulliparous women between menarche and <20 years*

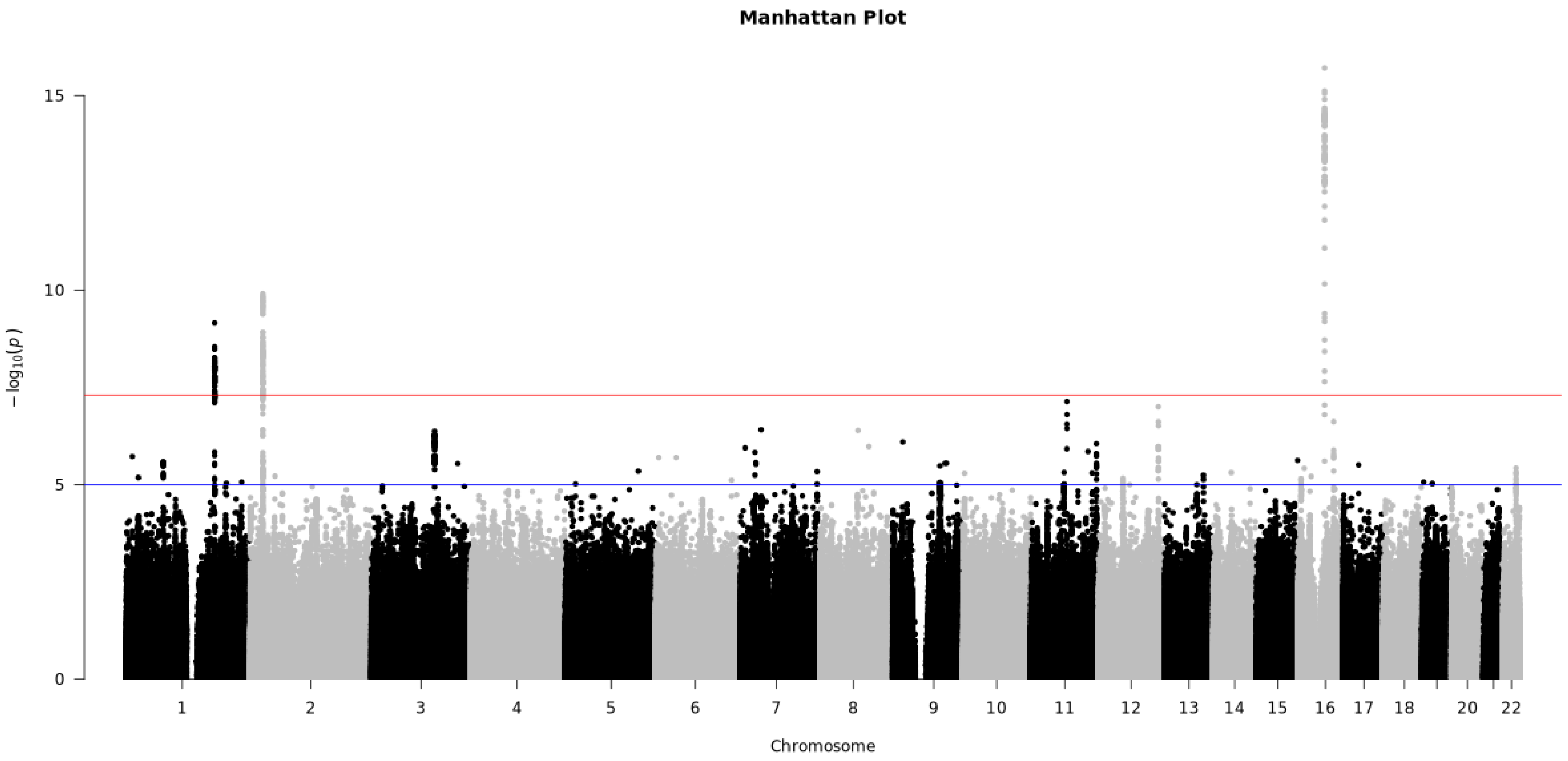

The x-axis represents the chromosomes, while the y-axis represents the -log10(p-values) of the SNPs. Horizontal lines are drawn at -log10($1 \times 10^{-5}$) for "suggestive associations" in blue and -log10($5 \times 10^{-8}$) for the "genome-wide significant" threshold in red.

*Figure S2B. QQ plot illustrating the observed vs. expected -log10(p-values) from the meta-analysed genome-wide association studies (GWAS) for BMI in nulliparous women between menarche and <20 years*

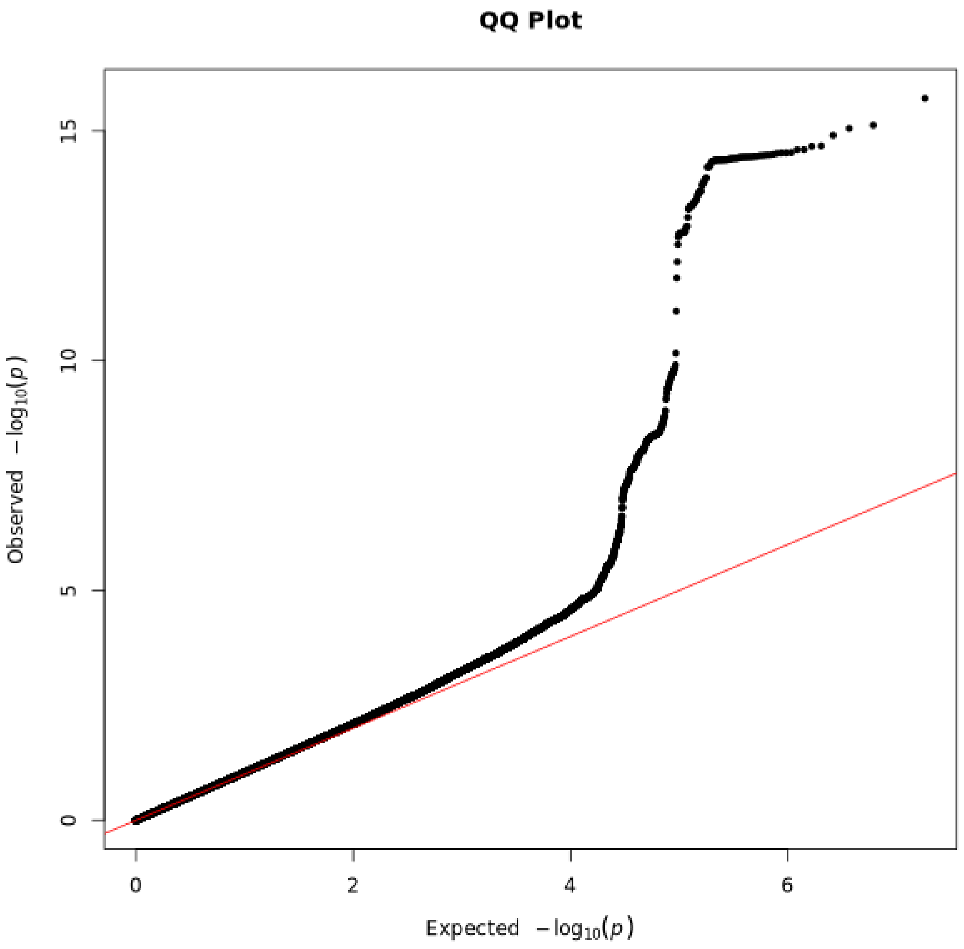

The red line represents the null hypothesis of no association.

*Figure S3A. Manhattan plot displaying the results of the meta-analysed genome-wide association studies (GWAS) for BMI in nulliparous women between 20 and <30 years*

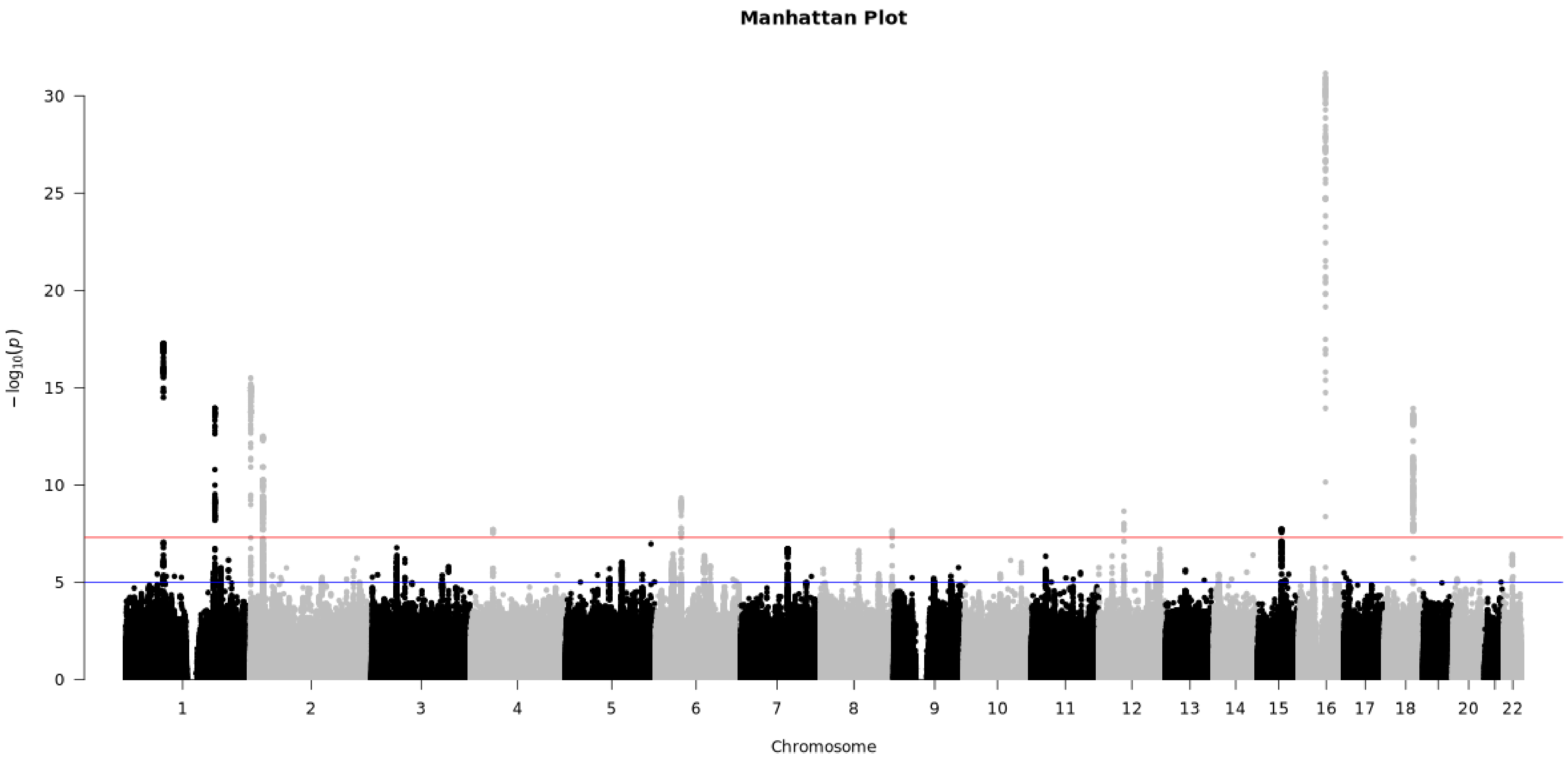

The x-axis represents the chromosomes, while the y-axis represents the -log10(p-values) of the SNPs. Horizontal lines are drawn at -log10($1 \times 10^{-5}$) for "suggestive associations" in blue and -log10($5 \times 10^{-8}$) for the "genome-wide significant" threshold in red.

*Figure S3B. QQ plot illustrating the observed vs. expected -log10(p-values) from the meta-analysed genome-wide association studies (GWAS) for BMI in nulliparous women between 20 and <30 years*

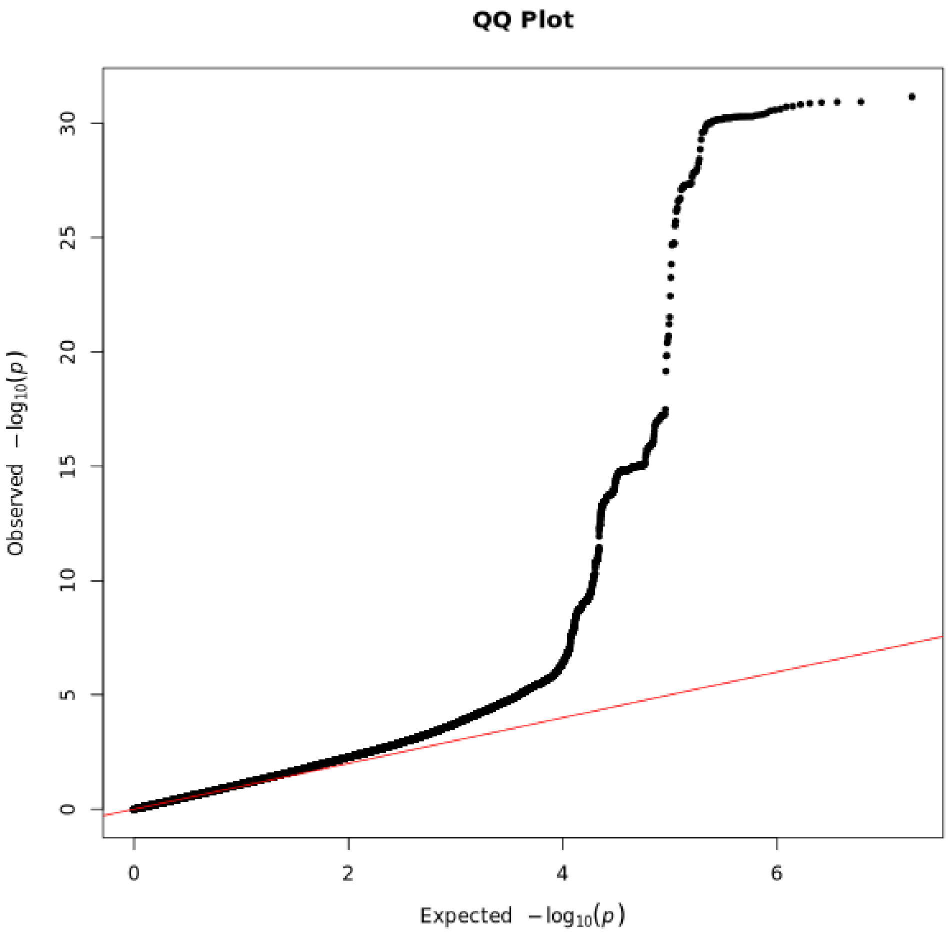

The red line represents the null hypothesis of no association.

*Figure S4A. Manhattan plot displaying the results of the meta-analysed genome-wide association studies (GWAS) for BMI in nulliparous women between 30 and <40 years*

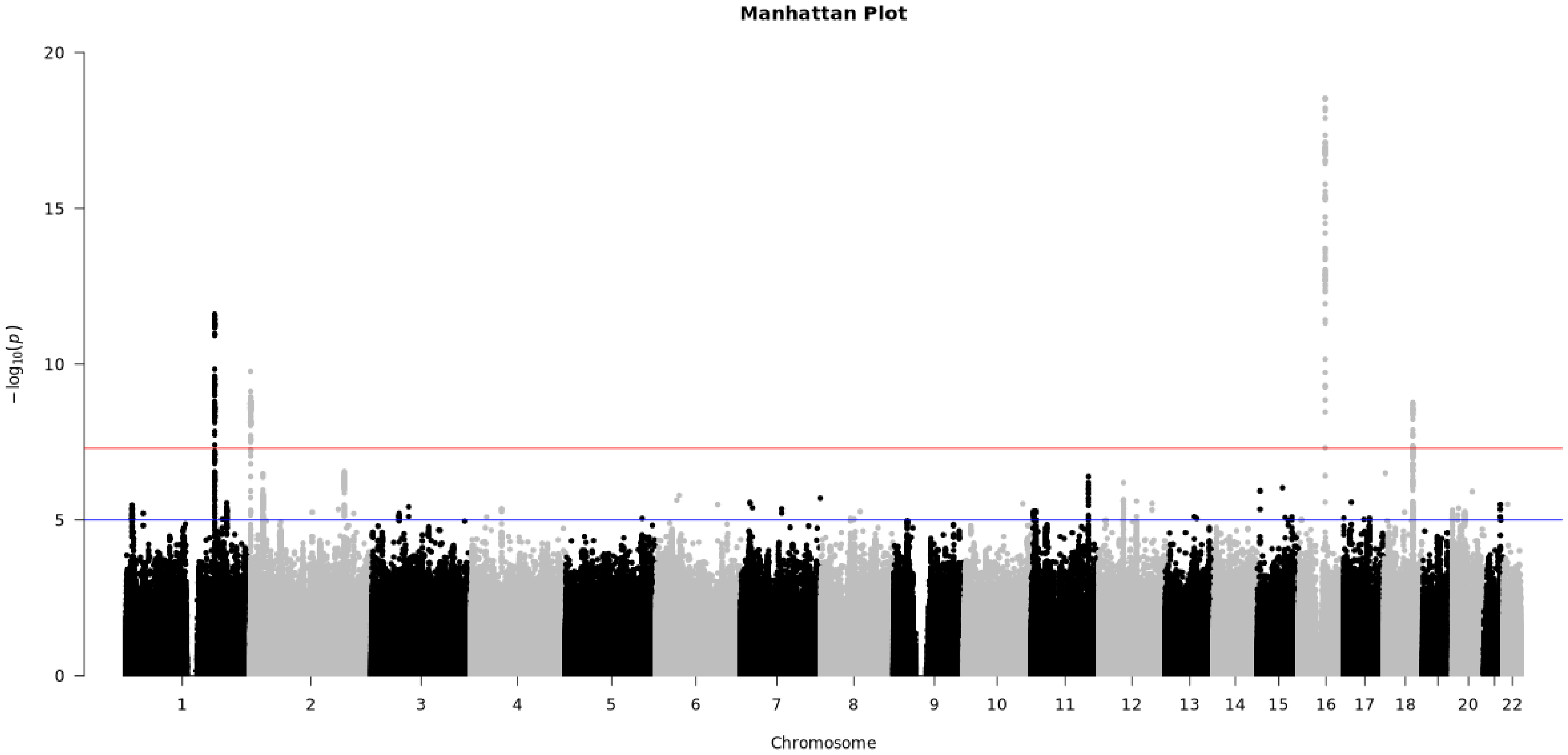

The x-axis represents the chromosomes, while the y-axis represents the -log10(p-values) of the SNPs. Horizontal lines are drawn at -log10(1 × $10^{-5}$) for "suggestive associations" in blue and -log10(5 × $10^{-8}$) for the "genome-wide significant" threshold in red.

*Figure S4B. QQ plot illustrating the observed vs. expected -log10(p-values) from the meta-analysed genome-wide association studies (GWAS) for BMI in nulliparous women between 30 and <40 years*

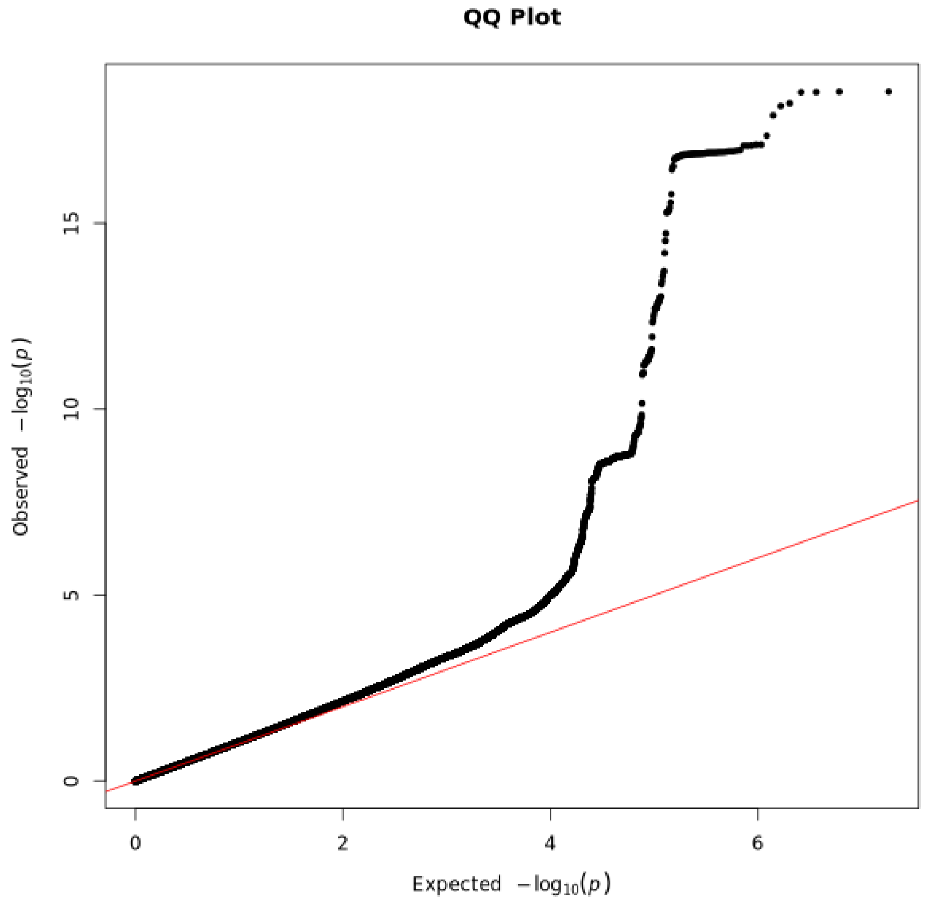

The red line represents the null hypothesis of no association.

*Figure S5. Plot illustrating the age-specific effect trajectories of individual SNPs on BMI across three defined age groups: menarche to <20 years, 20 to <30 years, and 30 to <40 years, in nulliparous women.*

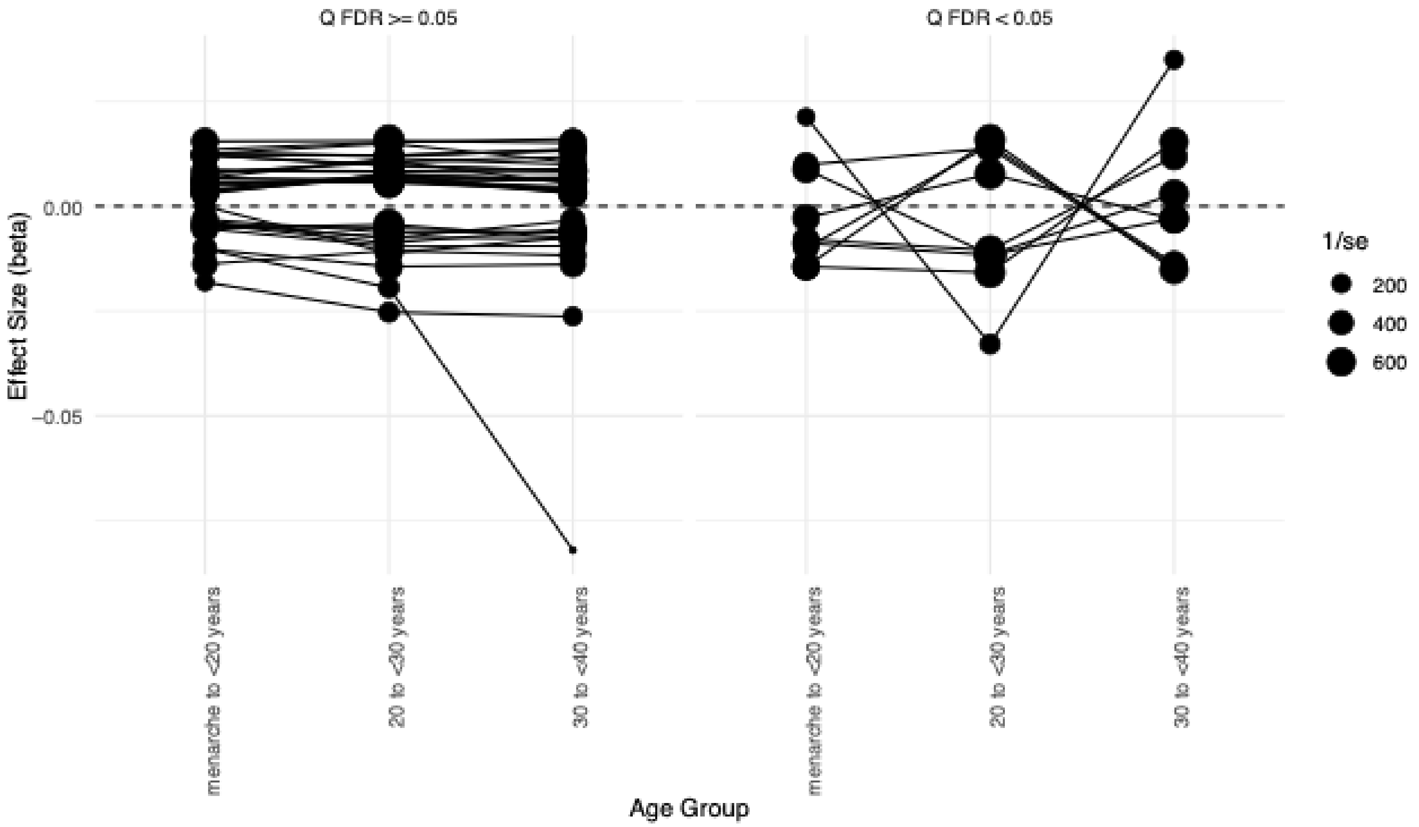

*Each line in the plot represents a single SNP, tracing its effect size (beta) across these age intervals. The point sizes are inversely proportional to the standard error of the effect estimates, meaning larger points indicate stronger associations with greater weight in the heterogeneity test. The plot facets divide SNPs based on whether they exhibit significant heterogeneity (p < 0.05, adjusted for multiple comparisons).*

*Table S1. STROBE-MR checklist of recommended items to address in reports of Mendelian randomization studies (15, 16).*

| Item No. | Section | Checklist item | Relevant text from manuscript |
|---|---|---|---|
| 1 | **TITLE and ABSTRACT** | Indicate Mendelian randomization (MR) as the study's design in the title and/or the abstract if that is a main purpose of the study | Abstract:<br><br>Results were meta-analysed, and two-sample Mendelian randomisation was applied within a lifecourse framework to assess the causal effect of BMI on breast cancer risk. |
| | **INTRODUCTION** | | |
| 2 | **Background** | Explain the scientific background and rationale for the reported study. What is the exposure? Is a potential causal relationship between exposure and outcome plausible? Justify why MR is a helpful method to address the study question | See entire Introduction and specifically:<br><br>We aimed to estimate the effect of higher BMI between menarche and first full-term pregnancy—or throughout early adulthood in individuals who remain nulliparous—on breast cancer risk later in life. This previously understudied period represents a critical window of susceptibility to exposures influencing breast cancer risk. We assess these effects on overall breast cancer and seven subtypes, including ER status (ER+/ER-) and five molecular subtypes: Luminal A, Luminal B1 (HER2+), Luminal B2 (HER2-), HER2-enriched, and triple-negative breast cancer using data from the Breast Cancer Association Consortium (BCAC) (17, 18), with the aim of exploring whether the effects of a higher BMI during this period differ in relation to menopausal status through associations with these breast cancer subtypes. |
| 3 | **Objectives** | State specific objectives clearly, including pre-specified causal hypotheses (if any). State that MR is a method that, under specific assumptions, intends to estimate causal effects | See above and the following:<br><br>Mendelian randomization (MR) is a technique that exploits the quasi-random distribution of genetic variants from parents to offspring, independent of the influence from other traits. Under specific assumptions, MR aims to estimate causal effects by reducing susceptibility to confounding factors, including confounding by undiagnosed existing disease and disease processes (reverse causation) (19, 20). Recent developments in lifecourse MR methodology include multivariable MR (MVMR) (21-23). This approach enables the direct estimation of the effects of an exposure measured at a specific life stage, controlling for the same exposure measured at another life stage, on later life outcomes. Previous MVMR analyses suggest that a larger prepubertal body size, employed as a proxy for BMI, is protective against breast cancer (24-26). This protective effect persists even after controlling for later life adult body size, which has a negligible effect on breast cancer risk when prepubertal body size is controlled for. These |

| | | | | |
|---|---|---|---|---|
| | | | | findings align with conventional analyses (27-29) and have been further supported through a proxy-genotype MR approach (30). |
| | **METHODS** | | | |
| 4 | **Study design and data sources** | | Present key elements of the study design early in the article. Consider including a table listing sources of data for all phases of the study. For each data source contributing to the analysis, describe the following: | See entire ‘Data sources and study design’ section, specifically:<br>We analysed five large population-based prospective cohorts with genomic and phenotypic BMI data from nulliparous women between menarche and <40 years. These included the Avon Longitudinal Study of Parents and Children, the Trøndelag Health Study, the Norwegian Mother, Father and Child Cohort Study, Generation R, and Generation Scotland. Participant selection is shown in Figure 1. |
| | | a) | Setting: Describe the study design and the underlying population, if possible. Describe the setting, locations, and relevant dates, including periods of recruitment, exposure, follow-up, and data collection, when available. | See entire ‘Data sources and study design’ section and Supplementary Text |
| | | b) | Participants: Give the eligibility criteria, and the sources and methods of selection of participants. Report the sample size, and whether any power or sample size calculations were carried out prior to the main analysis | See entire ‘Data sources and study design’ section and Supplementary Text |
| | | c) | Describe measurement, quality control and selection of genetic variants | See entire ‘Materials and methods’ section, specifically the ‘Genotyping, quality control and imputation in each cohort’ section and Supplementary text |
| | | d) | For each exposure, outcome, and other relevant variables, describe methods of assessment and diagnostic criteria for diseases | See entire ‘Data sources and study design’ section and Supplementary Text |
| | | e) | Provide details of ethics committee approval and participant informed consent, if relevant | Informed consent was obtained from all participants and ethical approval was obtained from the Regional Committee for Medical and Health Research Ethics, Central Norway (REK Central application number 2018/2488) (HUNT), the ALSPAC Ethics and Law Committee and the Local Research Ethics Committees (ALSPAC), the Medical Ethical Committee of Erasmus MC, University Medical Center Rotterdam, approved the study (MEC 198.782/2001/31) (Generation R), initial data collection on a license from the Norwegian Data Protection Agency and approval from The Regional Committees for Medical and Health Research Ethics (MoBa). The MoBa cohort is now based on regulations related to the Norwegian Health Registry Act. Written consent was obtained from all participants in Generation Scotland. All components of Generation Scotland received ethical approval from the NHS Tayside Committee on Medical Research Ethics (REC Reference Number: 05/S1401/89). Generation Scotland has also been granted Research Tissue Bank status by the East of Scotland Research Ethics Service (REC Reference Number: 20-ES-0021). |

| | | | |
|---|---|---|---|
| 5 | **Assumptions** | Explicitly state the three core IV assumptions for the main analysis (relevance, independence and exclusion restriction) as well assumptions for any additional or sensitivity analysis | See 'Lifecourse Mendelian randomization (MR) analysis' section, specifically:<br><br>Our primary two-sample MR analyses used the inverse variance weighted (IVW) estimator, implemented in the TwoSampleMR R package (31). When genetic variants are used as instrumental variables in MR, the assumptions of instrumental variables must be met, i.e., the genetic variants used must (i) be strongly associated with the exposure of interest and relevant to the population studied, here pregnant women ("relevance"), (ii) not share common causes with the outcome ("independence"), and (iii) not affect the outcome other than through the exposure ("exclusion-restriction") (32). We conducted sensitivity analyses, which relax the assumptions made about horizontal pleiotropy, including MR Egger regression (33), and the weighted median-based estimator (34). We ran IVW multivariable MR (MVMR), an extension of MR that employs multiple genetic variants associated with multiple measured risk factors, to calculate the direct and indirect effects of BMI between menarche and first birth on breast cancer outcomes. |
| 6 | **Statistical methods: main analysis** | Describe statistical methods and statistics used | See entire 'Statistical methods' section |
| | a) | Describe how quantitative variables were handled in the analyses (i.e., scale, units, model) | See 'Statistical methods' section |
| | b) | Describe how genetic variants were handled in the analyses and, if applicable, how their weights were selected | See entire 'Statistical methods' section with details varying throughout the manuscript. Variant selection is specified at relevant analytical stages. |
| | c) | Describe the MR estimator (e.g. two-stage least squares, Wald ratio) and related statistics. Detail the included covariates and, in case of two-sample MR, whether the same covariate set was used for adjustment in the two samples | Our primary two-sample MR analyses used the inverse variance weighted (IVW) estimator, implemented in the TwoSampleMR R package (31). When genetic variants are used as instrumental variables in MR, the assumptions of instrumental variables must be met, i.e., the genetic variants used must (i) be strongly associated with the exposure of interest and relevant to the population studied, here pregnant women ("relevance"), (ii) not share common causes with the outcome ("independence"), and (iii) not affect the outcome other than through the exposure ("exclusion-restriction") (32). We conducted sensitivity analyses, which relax the assumptions made about horizontal pleiotropy, including MR Egger regression (33), and the weighted median-based estimator (34). We ran IVW multivariable MR (MVMR), an extension of MR that employs multiple genetic variants associated with multiple measured risk factors, to calculate the direct and indirect effects of BMI between menarche and first birth on breast cancer outcomes. This analysis accounted for prepubertal and later-life adult body size separately (Figures 2A, B and C). These two additional time points were not included together in a single model due to the risk of |

| | | | | |
|---|---|---|---|---|
| | | | | weak instrument bias. Instrument strength was evaluated using the two-sample conditional F-statistic described elsewhere (35). |
| | | d) | Explain how missing data were addressed | A complete-case approach was used, excluding individuals with missing values from the analysis. Due to the skewed distribution of BMI in the data, a natural log transformation was applied prior to running the GWAS analyses within each cohort. GWAS analyses were carried out independently by individual studies, each following their own quality control (QC) procedures (Supplementary Material 2 and elsewhere (3, 4, 13, 14). All analyses were restricted post hoc to common genetic variants with a minor allele frequency (MAF) > 0.01. |
| | | e) | If applicable, indicate how multiple testing was addressed | NA |
| 7 | **Assessment of assumptions** | | Describe any methods or prior knowledge used to assess the assumptions or justify their validity | Our primary two-sample MR analyses used the inverse variance weighted (IVW) estimator, implemented in the TwoSampleMR R package (31). When genetic variants are used as instrumental variables in MR, the assumptions of instrumental variables must be met, i.e., the genetic variants used must (i) be strongly associated with the exposure of interest and relevant to the population studied, here pregnant women ("relevance"), (ii) not share common causes with the outcome ("independence"), and (iii) not affect the outcome other than through the exposure ("exclusion-restriction") (32). We conducted sensitivity analyses, which relax the assumptions made about horizontal pleiotropy, including MR Egger regression (33), and the weighted median-based estimator (34). |
| 8 | **Sensitivity analyses and additional analyses** | | Describe any sensitivity analyses or additional analyses performed (e.g. comparison of effect estimates from different approaches, independent replication, bias analytic techniques, validation of instruments, simulations) | To minimise pregnancy-related confounding, we restricted the BMI exposure GWAS to nulliparous women, enabling a clearer assessment of BMI's direct influence across life stages. In contrast, the breast cancer outcome GWAS included both nulliparous and parous women. This discrepancy in selection mechanisms introduces potential selection bias in the genetic effect estimates for BMI but not for breast cancer. Specifically, conditioning on parity in the BMI GWAS may induce bias in the estimated SNP effects on BMI ($\hat{\beta}_{Gx}$), whereas breast cancer estimates ($\hat{\beta}_{Gy}$), remain unaffected as parity was not conditioned on in the outcome GWAS.<br><br>In this two-sample MR setting, the MR estimate is given by:<br><br>$$\hat{\beta}_{MR} = \frac{\hat{\beta}_{Gy}}{\hat{\beta}_{Gx}}$$<br><br>Where:<br><br>$E(\hat{\beta}_{Gx}) = \beta_{Gx} + \text{bias}$<br><br>$E(\hat{\beta}_{Gy}) = \beta_{Gy}$ |

Since $\hat{\beta}_{Gy}$ is not subject to the same selection mechanism and there is no reason to assume that the selection-induced bias in $\beta_{Gx}$ is correlated with $\hat{\beta}_{Gy}$, a major spurious association in the MR analysis is unlikely. However, selection bias in the BMI GWAS could distort the MR estimate, with the direction and magnitude depending on whether and how BMI influences parity within the restricted sample. In an extreme scenario it could induce false positive associations with BMI that arise due to collider bias though this is unlikely at current sample sizes.

To empirically assess this, we performed univariable MR analyses estimating the effect of parity on BMI in nulliparous women across different life stages (menarche to <40 years, <20 years, 20 to <30 years, and 30 to <40 years). While parity itself cannot causally influence BMI in nulliparous women, genetic variants associated with parity may exhibit pleiotropic effects on BMI through shared metabolic and reproductive pathways. If these parity-associated variants also influence BMI within our restricted sample, it would suggest selection bias related to reproductive behaviour, implying that conditioning on parity in the BMI GWAS may have introduced collider bias.

In addition, a later onset of menarche has been linked to a reduced risk of breast cancer (36, 37). Childhood body size has been shown to accelerate the timing of menarche, whilst an earlier menarche increases the likelihood of increased body size in adulthood (38, 39). With these traits sharing a complex and interconnected relationship, we conduct sensitivity analyses estimating the effect of BMI in the lifestages analysed in nulliparous women on overall breast cancer accounting for age at menarche in MVMR analyses. Age at menarche is treated as a confounder, as it may influence both BMI and breast cancer risk through hormonal and metabolic pathways. Including it in the MVMR model ensures that observed BMI effects are not simply a reflection of differences in pubertal timing (pages 13-14)

**9 Software and pre-registration**

a) Name statistical software and package(s), including version and settings used

We meta-analysed summary statistics from the five cohorts for each lifestage-specific GWAS separately, using a fixed effects model employed in METAL version 2020-05-05 (40).

This was performed using PLINK (8) and genotype data from European individuals from phase 3 v5 enrolled in the 1000 genomes project as a reference panel (41).

Our primary two-sample MR analyses used the inverse variance weighted (IVW) estimator, implemented in the TwoSampleMR R package (31).

| | | | | |
|---|---|---|---|---|
| | | | | We conducted statistical analyses in R version 4.3.3 (42). |
| | | b) | State whether the study protocol and details were pre-registered (as well as when and where) | NA |
| | **RESULTS** | | | |
| 10 | **Descriptive data** | | | |
| | | a) | Report the numbers of individuals at each stage of included studies and reasons for exclusion. Consider use of a flow diagram | See Table 1 and Figure 1 and:<br><br>Measurements of body mass index (BMI) between menarche and age <40 years Measurements of body mass index (BMI) between menarche and age <40 years were available for 56,863 nulliparous women across five cohorts: the Avon Longitudinal Study of Parents and Children (ALSPAC), the Trøndelag Health Study (HUNT), the Norwegian Mother, Father and Child Cohort Study (MoBa), Generation R, and Generation Scotland. Corresponding phenotype sample sizes and cohort characteristics for age and BMI are presented in Table 1. Within the primary <40 year group, mean ages at measurement ranged from 23.9 years in ALSPAC to 29.4 years in Generation R, while mean BMI values varied between 22.8 kg/m² in ALSPAC and 24.5 kg/m² in Generation Scotland. To explore patterns across narrower age intervals within this broader period, we also conducted analyses stratified into three groups: menarche to <20 years, 20 to <30 years, and 30 to <40 years. Phenotypic sample sizes and summary measures of age and BMI for the full <40 group and each stratum are shown in Table 1.<br><br>.<br><br>*Lifestage-stratified genome-wide association studies (GWAS) and meta-analysis*<br><br>Genome-wide association analyses (GWAS) for BMI were conducted in up to 56,628 nulliparous women with measures between menarche and age <40 years, and results were meta-analysed across cohorts. Stratified meta-analyses for the three life stages – menarche to <20 years, 20 to <30 years, and 30 to <40 years – included 11,365, 30,272 and 16,565, respectively (Figure 1). For cohorts with repeat measures within one life stage (e.g., menarche to <40 years), we retained the time point with the largest sample size (e.g., for ALSPAC, only BMI data from participants <20 years were included). Considering results across all GWAS time windows (menarche to <20, 20 to <30, 30 to <40, and the combined menarche to <40), we identified a total of 45 independent variants. |
| | | b) | Report summary statistics for phenotypic exposure(s), outcome(s), and other relevant variables (e.g. means, SDs, proportions) | See Table 1 and able S2 in Supplementary |

| | | | | |
|---|---|---|---|---|
| | | c) | If the data sources include meta-analyses of previous studies, provide the assessments of heterogeneity across these studies | Cochran's Q statistic was used to evaluate heterogeneity among the 31 instrumental variables across the five cohort studies included in the primary analysis. To account for multiple comparisons, a Bonferroni correction was applied. After adjustment, none of the SNPs exhibited evidence of heterogeneity ($Q_{bonf}$<0.05), suggesting consistent effect estimates across studies. This reinforces their validity in downstream MR analyses, reducing concerns that study-level differences may confound associations with the outcome. |
| | | d) | For two-sample MR:<br>i. Provide justification of the similarity of the genetic variant-exposure associations between the exposure and outcome samples<br>ii. Provide information on the number of individuals who overlap between the exposure and outcome studies | See 'Data sources and study design' in the 'Materials and methods' section |
| 11 | **Main results** | | | |
| | | a) | Report the associations between genetic variant and exposure, and between genetic variant and outcome, preferably on an interpretable scale | See Table S3 in Supplementary. |
| | | b) | Report MR estimates of the relationship between exposure and outcome, and the measures of uncertainty from the MR analysis, on an interpretable scale, such as odds ratio or relative risk per SD difference | See Table S5 in Supplementary and the 'Lifecourse Mendelian Randomization (MR) analysis' section in Results, specifically:<br>Univariable MR analyses using the 31 identified SNPs indicated strong evidence that higher genetically predicted BMI in nulliparous women between menarche and <40 years reduced the risk of overall breast cancer (IVW odds ratio (OR), 95% CI: 0.76, 0.67 to 0.86, P =1.27x10-5) (Figure 5; Table S5). Higher genetically predicted BMI in nulliparous women between menarche and <40 years additionally reduced the risk of most breast cancer subtypes, with the exception of the HER-2 enriched subtype. In multivariable MR accounting for adult body size, there remained evidence that higher genetically predicted BMI in nulliparous women between menarche and <40 years reduced the risk of overall breast cancer and most breast cancer subtypes, with the exception of the HER-2 enriched subtype. Adjustment for prepubertal body size led to a marked, though not complete, attenuation of these effects, consistent with part of the protection being shared with childhood adiposity.<br>In the narrower life-stage periods obtained by partitioning the broader fertile-window measure, univariable MR analyses indicated evidence that higher genetically predicted BMI in nulliparous women between menarche to <20, 20 to <30, and 30 to <40 years reduced the risk of overall breast cancer and most subtypes, although the strength and precision of effects varied by subtype and life-stage period. After accounting for later life adult body size, protective effects were largely retained, with most estimates still supporting a reduction in risk. In contrast, adjusting for prepubertal body size removed much of the |

| | | | | |
|---|---|---|---|---|
| | | | | apparent protection in these narrower periods (Figure 5; Table S5). Consistent patterns of associations were observed using the weighted median method employed for robustness. In addition, results using the MR-Egger method did not provide evidence that horizontal pleiotropy was responsible for the estimates derived (Table S5).<br>Multivariable MR results should be interpreted with caution since conditional F-statistic was <10 for each of the BMI exposures included in these models, indicating potential for weak instrument bias (Table S5). |
| | | c) | If relevant, consider translating estimates of relative risk into absolute risk for a meaningful time period | NA |
| | | d) | Consider plots to visualize results (e.g. forest plot, scatterplot of associations between genetic variants and outcome versus between genetic variants and exposure) | See entire 'Results' section as well as Supplementary Figures S1-S5. |
| 12 | **Assessment of assumptions** | | | |
| | | a) | Report the assessment of the validity of the assumptions | See Table S5 in Supplementary and the 'Lifecourse Mendelian Randomization (MR) analysis' section in Results, specifically:<br>Consistent patterns of associations were observed using the weighted median method employed for robustness. In addition, results using the MR-Egger method did not provide evidence that horizontal pleiotropy was responsible for the estimates derived (Table S5). |
| | | b) | Report any additional statistics (e.g., assessments of heterogeneity across genetic variants, such as $I^2$, Q statistic or E-value) | See entire 'Lifestage-stratified genome-wide association studies (GWAS) and meta-analysis' section in Results |
| 13 | **Sensitivity analyses and additional analyses** | | | |
| | | a) | Report any sensitivity analyses to assess the robustness of the main results to violations of the assumptions | See Table S5 in Supplementary and the 'Lifecourse Mendelian Randomization (MR) analysis' section in Results, specifically:<br>Consistent patterns of associations were observed using the weighted median method employed for robustness. In addition, results using the MR-Egger method did not provide evidence that horizontal pleiotropy was responsible for the estimates derived (Table S5). |
| | | b) | Report results from other sensitivity analyses or additional analyses | Our analyses to investigate potential collider bias indicated very little evidence that genetically proxied parity associated with BMI in nulliparous women between menarche and <40 years, menarche and <20 years, and 20 and <30 years. There was some evidence that genetically proxied parity associated with lower BMI in nulliparous women between 30 and <40 years (Table S6). These results suggest |

| | | | |
|---|---|---|---|
| | | | collider bias due to selecting nulliparous women is likely to be of little concern within this study.<br><br>After adjusting for age at menarche, the estimated effect of BMI in nulliparous women between menarche and <40 years on overall breast cancer remains largely consistent with the UVMR analyses (Table S7). Age at menarche itself showed little direct effect on overall breast cancer or most subtypes once BMI was accounted for. |
| | | c) Report any assessment of direction of causal relationship (e.g., bidirectional MR) | |
| | | d) When relevant, report and compare with estimates from non-MR analyses | See Discussion section. |
| | | e) Consider additional plots to visualize results (e.g., leave-one-out analyses) | See entire 'Results' section. |
| | **DISCUSSION** | | |
| 14 | **Key results** | Summarize key results with reference to study objectives | In this study, we undertook GWAS and MR analyses within a lifecourse framework to (i) assess the consistency of the genetic effects of BMI across different life stages, and (ii) investigate the effects of genetically proxied BMI in nulliparous women between menarche and <40 years on the risk of overall breast cancer and seven subtypes. This work builds on previous research to further the investigation into the influence of BMI during critical periods on breast cancer risk (24, 25, 43). Motivated by recent findings suggesting that a larger prepubertal body size, used as a proxy for BMI, may offer protection against breast cancer risk, while later life body size shows very little effect, we aimed to better understand the interval between puberty and later life. Specifically, we focused on the period between menarche and first full-term pregnancy, a crucial window of vulnerability for later life breast cancer development.<br><br>We observed considerable variation in genetic effects on BMI across different life stages for nine of the 45 discovery variants identified in this study. In addition, genome-wide cross-lifestage genetic correlations demonstrated variation, e.g., when comparing prepubertal body size and later-life body size with BMI from menarche to 40 years. In the univariable MR analyses, higher BMI in nulliparous women from menarche to <40 years was found to consistently reduce risk of breast cancer, including BMI measured in the separate age periods within this life stage (menarche to <20 years, 20 to <30, and 30 to <40 years), and risk of overall breast cancer and its subtypes. These protective effects largely remained upon accounting for later life adult body size. In contrast, adjusting for prepubertal body size in multivariable MR led to greater attenuation, indicating that part of the protective effect of BMI |

| | | | |
|---|---|---|---|
| | | | in early reproductive life may be explained by body size before puberty. The attenuation was more pronounced for the narrower life-stage periods than for the full fertile-window measure, reflecting both biological and methodological influences. Biologically, part of the apparent effect across all reproductive-age windows overlaps with childhood adiposity, so adjustment for prepubertal body size reduces the estimates. Methodologically, the narrower age-specific GWASs have substantially smaller sample sizes than the childhood GWAS. In multivariable MR, this imbalance can magnify the degree of attenuation by limiting power to detect effects independent of childhood size. By contrast, the broader reproductive-age measure is based on a larger GWAS sample and therefore shows less attenuation, despite adjustment for the same childhood body size data. Importantly, no such attenuation was observed when adjusting for later-life body size, even though similar sample size imbalances exist. This indicates that differential power alone may not fully explain the observed differences. Future methodological work is needed to examine how sample size disparities across life-stage GWASs influence estimates in multivariable MR and to develop strategies for accounting for this in lifecourse applications. |
| 15 | **Limitations** | Discuss limitations of the study, taking into account the validity of the IV assumptions, other sources of potential bias, and imprecision. Discuss both direction and magnitude of any potential bias and any efforts to address them | This study has several limitations that should be considered when interpreting the findings. First, low conditional F-statistics were observed in the MVMR analysis, indicating potential for weak instrument bias. As a result, our MVMR findings should be interpreted with some caution. Second, while we combined data from five large European longitudinal cohorts, the sample size remains smaller (N=90,115) than that available for comparable measures of prepubertal and later-life body size derived using in UK Biobank data (N=246,511). This reduced sample size may limit statistical power for detecting smaller effect sizes. Third, used a prepubertal GWAS based on reported body size rather than measured BMI, as it offered a much larger sample size than any other GWAS of BMI at this life stage. However, this choice may distort MVMR analysis due to measurement error or differences in variance of effect sizes, and future work to precisely estimate childhood body size genetic effects are warranted. Fourth, there is the possibility of participant overlap between the HUNT and MoBa cohorts. While HUNT recruited individuals from Trøndelag County, Norway, MoBa had a national recruitment strategy. Some participants in HUNT may have also participated in MoBa, which could introduce a degree of sample duplication. Lastly, as our analysis was restricted to individuals of |

| | | | |
|---|---|---|---|
| | | | European ancestry, the generalisability of our findings to other populations is limited. Further research is needed to confirm these findings in more diverse populations. |
| 16 | **Interpretation** | | |
| | | a) Meaning: Give a cautious overall interpretation of results in the context of their limitations and in comparison with other studies | See entire Discussion section, including Conclusion:<br>This study offers important insights into the genetic influences on BMI across different life stages and its causal relationship with breast cancer risk, focusing on the period between menarche and under 40 years in nulliparous women as a key window of susceptibility. While higher BMI in this interval appeared strongly protective against breast cancer in univariable analyses, the effect substantially, though not entirely, attenuated after accounting for childhood body size, used as a proxy for BMI. This pattern suggests that the protection may arise from the combined influence of greater adiposity in both childhood and early adulthood. These results have important implications for breast cancer prevention, underscoring earlier life stages as critical periods for potential interventions. |
| | | b) Mechanism: Discuss underlying biological mechanisms that could drive a potential causal relationship between the investigated exposure and the outcome, and whether the gene-environment equivalence assumption is reasonable. Use causal language carefully, clarifying that IV estimates may provide causal effects only under certain assumptions | The relationship between body size and breast cancer risk is complex. In conventional epidemiological settings, body mass index (BMI) has been linked to an increased risk of breast cancer in postmenopausal women and a decreased risk of breast cancer in premenopausal women (44-47). A plausible explanation includes the differing levels of oestrogen exposure between women experiencing overweight and normal-weight (48). Premenopausal women experiencing overweight tend to have longer anovulatory cycles, which reduces their exposure to ovarian hormones, potentially lowering their breast cancer risk. After menopause, fat tissue serves as another source of oestrogen production, which increases breast cancer risk among overweight women (49-52). Whilst our study did not specifically examine the relationship between overweight status and breast cancer pre and post menopause, effect estimates showed minimal variation between increased BMI and any of the breast cancer subtypes investigated in our study, with the exception of HER-2 enriched. Specifically, increased BMI in the 20 to <30-year category shifted from having a protective effect in univariable MR analyses to an increased risk of the HER-2 enriched subtype after accounting for body size later in life, though evidence supporting this effect is limited. There are likely complex interactions between body fat distribution, metabolic health, and HER-2 driven cell growth, potentially explaining why the HER-2 enriched subtype exhibited distinct patterns in response to higher BMI at different ages. |

| | | | |
|---|---|---|---|
| | | | This pattern is not observed in Luminal B1 (HER2+/ER+) tumours, suggesting that there may be a unique effect of HER-2 expression specifically in non-hormonally regulated tumours. |
| | | c) Clinical relevance: Discuss whether the results have clinical or public policy relevance, and to what extent they inform effect sizes of possible interventions | This study is an important and novel analysis with multiple strengths. It focuses on a specific age range where the protective effect of BMI on breast cancer risk begins to wane —an insight not previously achieved using causal inference methods. By examining this critical period between menarche and first birth, we enhance understanding on the potential interaction of nulliparity and BMI on breast cancer development. Focusing on this timeframe also allowed us to obtain an estimate that minimised the confounding effects introduced by the physiological changes associated with a first pregnancy. To strengthen the analysis, we integrated data from five large European longitudinal cohort studies, generating robust GWAS results on BMI. This approach allowed us to gain key insights into the consistency of BMI-related genetic effects across different stages of life. In addition, these data are not only useful for this particular study but also offer a valuable resource for future research into the effects of BMI in women at these life stages on other health outcomes. |
| 17 | **Generalizability** | Discuss the generalizability of the study results (a) to other populations, (b) across other exposure periods/timings, and (c) across other levels of exposure | Lastly, as our analysis was restricted to individuals of European ancestry, the generalisability of our findings to other populations is limited. Further research is needed to confirm these findings in more diverse populations. |
| | **OTHER INFORMATION** | | |
| 18 | **Funding** | Describe sources of funding and the role of funders in the present study and, if applicable, sources of funding for the databases and original study or studies on which the present study is based | GMP, AH, GL, ES, RR, GH, and GDS were supported by the Integrative Epidemiology Unit which receives funding from the UK Medical Research Council and the University of Bristol (MC_UU_00032/1). GDS conducts research at the NIHR Biomedical Research Centre at the University Hospitals Bristol NHS Foundation Trust and the University of Bristol. The views expressed in this publication are those of the author(s) and not necessarily those of the NHS, the National Institute for Health Research or the Department of Health. The genotyping in HUNT was supported by the National Institutes of Health (NIH); University of Michigan; The Research Council of Norway (RCN); The Liaison Committee for Education, Research and Innovation in Central Norway; and the Joint Research Committee between St. Olavs hospital and the Faculty of Medicine and Health Sciences, NTNU. LB, BOA, and |

BMB work in a research unit financially supported by the Liaison Committee for education, research and innovation in Central Norway and the Joint Research Committee between St. Olavs Hospital and the Faculty of Medicine and Health Sciences, NTNU. This study used data from Medical Birth Registry of Norway (MBRN). Generation Scotland received core support from the Chief Scientist Office of the Scottish Government Health Directorates [CZD/16/6] and the Scottish Funding Council [HR03006] and is currently supported by the Wellcome Trust [216767/Z/19/Z]. Genotyping of the GS:SFHS samples was carried out by the Genetics Core Laboratory at the Edinburgh Clinical Research Facility, University of Edinburgh, Scotland and was funded by the Medical Research Council UK and the Wellcome Trust (Wellcome Trust Strategic Award "STratifying Resilience and Depression Longitudinally" (STRADL) Reference 104036/Z/14/Z. CH was supported by an MRC University Unit core grant MC_UU_00007/10 (QTL in Health and Disease program). BLL acknowledges support from the University of Bristol (Vice-Chancellor's Research Fellowship), Academy of Medical Sciences/Wellcome Trust/the Government Department of Business, Energy and Industrial Strategy/British Heart Foundation/Diabetes UK Springboard Award (SBF003/1170), Elizabeth Blackwell Institute for Health Research (University of Bristol), and Wellcome Trust Institutional Strategic Support Fund (204813/Z/16/Z) and Wellcome Trust Career Development Award (227849/Z/23/Z). AHavdahl was supported by the Research Council of Norway (#336085) and the South-Eastern Norway Regional Health Authority (#2020022; # 2922083; #2019097; #2018059; #2021045). The general design of the Generation R Study is made possible by financial support from Erasmus MC, University Medical Center Rotterdam, Erasmus University Rotterdam, the Netherlands Organization for Health Research and Development (ZonMw), the Netherlands Organization for Scientific Research (NWO), the Ministry of Health, Welfare and Sport, and the Ministry of Youth and Families. The parental genotyping in Generation R was supported by the ERC under the European Union's Horizon 2020 research and innovation programme (iRISK; grant agreement No 863981).

This project received funding from the European Union's Horizon Europe Research and Innovation Programme under grant agreement nº 101137146 (STAGE project). UK participants in Horizon Europe Project STAGE are supported by UKRI grant numbers 10112787 (Beta Technology), 10099041 (University of Bristol) and 10109957 (Imperial College London).

| | | | |
|---|---|---|---|
| 19 | **Data and data sharing** | Provide the data used to perform all analyses or report where and how the data can be accessed and reference these sources in the article. Provide the statistical code needed to reproduce the results in the article, or report whether the code is publicly accessible and if so, where | All genetic instruments derived in this study are in the supplementary tables. Genome-wide association study summary statistics will be made available on a public repository upon publication. |
| 20 | **Conflicts of Interest** | All authors should declare all potential conflicts of interest | Authors declare no conflict of interest. |

This checklist is copyrighted by the Equator Network under the Creative Commons Attribution 3.0 Unported (CC BY 3.0) license.

*Table S2. Summary of breast cancer GWAS datasets from the BCAC 2017 and BCAC 2020 releases.*

| Phenotype | Subtype | Receptor/grade status | Lead author name | Pubmed ID | Release | Sample size (female) | Cases | Controls | % cases |
|---|---|---|---|---|---|---|---|---|---|
| Breast cancer | ER+ | ER+ | Michailidou K | 29059683 | 2017 | 175,475 | 69,501 | 105,974 | 39.60% |
| Breast cancer | ER- | ER- | | | | 127,442 | 21,468 | 105,974 | 16.90% |
| Breast cancer | Luminal A | ER+ and/or PR+, HER2−, grades 1 and 2 | Zhang H | 32424353 | 2020 | 155,244 | 63,767 | 91,477 | 41.10% |
| Breast cancer | Luminal B1 | ER+ and/or PR+, HER2+ | | | | 107,419 | 15,942 | 91,477 | 14.80% |
| Breast cancer | Luminal B2 | ER+ and/or PR+, HER2−, grade 3 | | | | 107,419 | 15,942 | 91,477 | 14.80% |
| Breast cancer | HER2-enriched | ER− and PR−, HER2+ | | | | 102,105 | 10,628 | 91,477 | 10.40% |
| Breast cancer | TNBC | ER− and PR−, HER2− | | | | 100,079 | 8,602 | 91,477 | 8.60% |
| Prepubertal body size | N/A | N/A | Richardson TG | 32376654 | 2020 | 246,511 | N/A | N/A | N/A |
| Later life adult body size | N/A | N/A | | | | 246,511 | N/A | N/A | N/A |

*Table S3. Meta-analysed genome-wide association study results for BMI in nulliparous women between menarche and <40 years, menarche and <20 years, 20 and <30 years and 30 and <40 years.*

SNP - single nucleotide polymorphism identifier, beta^ - effect estimate coefficient for SNP on log-transformed BMI, beta - effect estimate coefficient for SNP on body size category, SE- standard error of the effect estimate, P - corresponding p-value, * - indicates SNPs retained in the final set identified through LD clumping across all time windows.

- Independent discovery SNPs for BMI in nulliparous women between menarche and <40 years
- Independent discovery SNPs for BMI in nulliparous women between menarche and <20 years
- Independent discovery SNPs for BMI in nulliparous women between 20 and <30 years
- Independent discovery SNPs for BMI in nulliparous women between 30 and <40 years

| | | | | | | | Age group cateogory | | | | | | | | | | | | | | | | | | | | | | | |
|---|---|---|---|---|---|---|---|---|---|---|---|---|---|---|---|---|---|---|---|---|---|---|---|---|---|---|---|---|---|---|
| | | | | | | | BMI in nulliparous women between menarche and <40 years | | | | BMI in nulliparous women between menarche and <20 years | | | | BMI in nulliparous women between 20 and <30 years | | | | BMI in nulliparous women between 30 and <40 years | | | | Prepubertal body size (Richardson et al.) | | | | Later life adult body size (Richardson et al.) | | | |
| SNP | Chromosome | Base position (build hg19) | Base position (build hg38) | Closest gene | Effect allele | Other allele | Effect allele frequency | Beta^ | Standard Error | P | Effect allele frequency | Beta^ | Standard Error | P | Effect allele frequency | Beta^ | Standard Error | P | Effect allele frequency | Beta^ | Standard Error | P | Effect allele frequency | Beta | Standard Error | P | Effect allele frequency | Beta | Standard Error | P |
| rs62033406 | 16 | 53824226 | 53790314 | FTO | G | A | 0.418 | 0.016 | 0.001 | 1.15E-61 | 0.418 | 0.016 | 0.002 | 1.95E-16 | 0.421 | 0.016 | 0.001 | 2.63E-31 | 0.417 | 0.015 | 0.002 | 1.26E-18 | 0.408 | 0.045 | 0.002 | 8.80E-118 | 0.408 | 0.042 | 0.002 | 1.20E-98 |
| rs7707628* | 5 | 153546900 | 154167340 | MFAP3 | C | T | 0.664 | -0.006 | 0.001 | 6.17E-09 | 0.669 | -0.003 | 0.002 | 1.02E-01 | 0.665 | -0.006 | 0.001 | 6.74E-06 | 0.656 | -0.006 | 0.002 | 2.87E-04 | 0.646 | -0.015 | 0.002 | 9.20E-14 | 0.646 | -0.011 | 0.002 | 1.10E-07 |
| rs60186497* | 14 | 30479220 | 30010014 | PRKD1 | T | C | 0.037 | 0.014 | 0.003 | 2.76E-08 | 0.039 | 0.013 | 0.005 | 1.16E-02 | 0.036 | 0.015 | 0.004 | 2.96E-05 | 0.037 | 0.011 | 0.005 | 1.98E-02 | 0.044 | 0.030 | 0.005 | 2.70E-10 | 0.044 | 0.021 | 0.005 | 1.50E-05 |
| rs2601781* | 16 | 4017392 | 3967391 | ADCY9 | A | T | 0.632 | 0.006 | 0.001 | 4.20E-09 | 0.631 | 0.006 | 0.002 | 2.70E-03 | 0.631 | 0.006 | 0.001 | 1.56E-05 | 0.633 | 0.004 | 0.002 | 1.80E-02 | 0.618 | 0.017 | 0.002 | 4.20E-18 | 0.618 | 0.016 | 0.002 | 2.10E-14 |
| rs13130484* | 4 | 45175691 | 45173674 | THAP12P9; PRDX4P1 | T | C | 0.414 | 0.006 | 0.001 | 7.04E-11 | 0.415 | 0.004 | 0.002 | 4.59E-02 | 0.413 | 0.008 | 0.001 | 1.34E-08 | 0.419 | 0.006 | 0.002 | 1.18E-04 | 0.432 | 0.023 | 0.002 | 4.20E-32 | 0.432 | 0.017 | 0.002 | 7.80E-17 |
| rs6098816* | 20 | 54385642 | 55810586 | CBLN4 | A | C | 0.174 | -0.007 | 0.001 | 4.74E-08 | 0.173 | -0.004 | 0.003 | 1.34E-01 | 0.173 | -0.008 | 0.002 | 2.72E-05 | 0.174 | -0.003 | 0.002 | 1.57E-01 | 0.808 | -0.015 | 0.002 | 4.80E-09 | 0.808 | -0.002 | 0.002 | 4.00E-01 |
| rs28693508* | 9 | 120361418 | 117599140 | TLR4 | G | C | 0.354 | -0.005 | 0.001 | 2.19E-08 | 0.351 | -0.005 | 0.002 | 2.09E-02 | 0.354 | -0.005 | 0.001 | 1.82E-04 | 0.352 | -0.007 | 0.002 | 2.79E-04 | 0.339 | -0.011 | 0.002 | 1.10E-07 | 0.339 | -0.009 | 0.002 | 1.10E-05 |
| rs62155011* | 2 | 105565719 | 104949261 | MRPS9 | C | T | 0.404 | 0.006 | 0.001 | 1.19E-08 | 0.411 | 0.006 | 0.002 | 4.21E-03 | 0.405 | 0.006 | 0.001 | 8.57E-06 | 0.395 | 0.005 | 0.002 | 8.91E-03 | 0.375 | 0.004 | 0.002 | 3.30E-02 | 0.375 | 0.007 | 0.002 | 9.10E-04 |
| rs574367* | 1 | 177873210 | 177904075 | LINC01741, SEC16B | T | G | 0.226 | 0.014 | 0.001 | 2.82E-35 | 0.226 | 0.013 | 0.002 | 2.22E-08 | 0.227 | 0.012 | 0.002 | 1.24E-14 | 0.221 | 0.013 | 0.002 | 8.58E-12 | 0.209 | 0.053 | 0.002 | 1.30E-112 | 0.209 | 0.036 | 0.002 | 6.60E-51 |
| rs59086897* | 2 | 25145173 | 24922304 | ADCY3,DNAJC27 | A | T | 0.489 | 0.010 | 0.001 | 1.27E-24 | 0.494 | 0.012 | 0.002 | 1.38E-10 | 0.489 | 0.010 | 0.001 | 4.85E-14 | 0.487 | 0.008 | 0.002 | 3.181E-07 | 0.487 | 0.040 | 0.002 | 1.30E-95 | 0.487 | 0.024 | 0.002 | 2.60E-34 |
| rs10191896* | 2 | 213421764 | 212557040 | ERBB4 | T | C | 0.371 | -0.010 | 0.002 | 3.50E-08 | 0.362 | 0.000 | 0.004 | 9.94E-01 | 0.368 | -0.011 | 0.002 | 5.95E-07 | 0.375 | -0.007 | 0.003 | 1.59E-02 | 0.376 | -0.001 | 0.002 | 5.60E-01 | 0.376 | -0.005 | 0.002 | 1.00E-02 |
| rs11615823* | 12 | 26815139 | 26662206 | BHLHE41 | C | T | 0.031 | -0.015 | 0.003 | 3.17E-08 | 0.033 | -0.010 | 0.005 | 5.29E-02 | 0.031 | -0.019 | 0.004 | 4.40E-07 | 0.031 | -0.082 | 0.143 | 5.67E-01 | 0.030 | -0.003 | 0.006 | 6.80E-01 | 0.030 | -0.010 | 0.006 | 9.40E-02 |
| rs2764261* | 6 | 108927842 | 108606639 | FOXO3 | G | A | 0.632 | 0.005 | 0.001 | 2.52E-08 | 0.628 | 0.006 | 0.002 | 2.74E-03 | 0.637 | 0.006 | 0.001 | 4.35E-05 | 0.627 | 0.004 | 0.002 | 2.18E-02 | 0.625 | 0.008 | 0.002 | 4.00E-05 | 0.625 | 0.012 | 0.002 | 1.40E-08 |
| rs7613360* | 3 | 49916710 | 49879277 | MST1R,ACTL11P | T | C | 0.392 | 0.006 | 0.001 | 3.22E-09 | 0.392 | 0.003 | 0.002 | 8.60E-02 | 0.397 | 0.007 | 0.001 | 1.56E-07 | 0.385 | 0.004 | 0.002 | 2.60E-02 | 0.394 | 0.005 | 0.002 | 1.60E-02 | 0.394 | 0.014 | 0.002 | 3.90E-12 |
| rs13387091* | 2 | 650980 | 650980 | LINC01875, TMEM18 | A | G | 0.166 | -0.013 | 0.001 | 1.83E-26 | 0.167 | -0.010 | 0.003 | 3.90E-05 | 0.165 | -0.014 | 0.002 | 3.40E-16 | 0.169 | -0.014 | 0.002 | 1.95E-10 | 0.172 | -0.044 | 0.003 | 1.20E-67 | 0.172 | -0.036 | 0.003 | 1.40E-43 |
| rs6494678* | 15 | 67946247 | 67653909 | MAP2K5 | T | C | 0.241 | -0.006 | 0.001 | 3.88E-08 | 0.239 | -0.004 | 0.002 | 5.77E-02 | 0.240 | -0.009 | 0.002 | 1.80E-08 | 0.236 | -0.005 | 0.002 | 8.62E-03 | 0.254 | -0.021 | 0.002 | 1.10E-20 | 0.254 | -0.016 | 0.002 | 1.20E-12 |
| rs114670539* | 2 | 207064335 | 206199611 | CMKLR2 | T | C | 0.041 | 0.014 | 0.002 | 2.08E-09 | 0.040 | 0.014 | 0.005 | 6.06E-03 | 0.041 | 0.015 | 0.003 | 4.55E-06 | 0.045 | 0.016 | 0.004 | 6.94E-05 | 0.058 | 0.041 | 0.004 | 9.80E-23 | 0.058 | 0.007 | 0.004 | 8.60E-02 |
| rs7442885* | 5 | 87682877 | 88387060 | TMEM161B-DT | G | C | 0.197 | -0.007 | 0.001 | 4.31E-08 | 0.199 | -0.004 | 0.002 | 8.42E-02 | 0.198 | -0.008 | 0.002 | 4.55E-06 | 0.197 | -0.008 | 0.002 | 1.97E-04 | 0.214 | -0.012 | 0.002 | 2.00E-06 | 0.214 | -0.017 | 0.002 | 1.50E-12 |

| | | | | | | | | | | | | | | | | | | | | | | | | | | | | | | |
|---|---|---|---|---|---|---|---|---|---|---|---|---|---|---|---|---|---|---|---|---|---|---|---|---|---|---|---|---|---|---|
| rs7462788* | 8 | 143335170 | 142253809 | PTK2 | T | G | 0.398 | 0.006 | 0.001 | 3.34E-09 | 0.398 | 0.003 | 0.002 | 1.17E-01 | 0.398 | 0.008 | 0.001 | 2.40E-08 | 0.394 | 0.003 | 0.002 | 8.67E-02 | 0.395 | 0.003 | 0.002 | 1.00E-01 | 0.395 | 0.008 | 0.002 | 7.70E-05 |
| rs10840050* | 11 | 8473707 | 8452160 | STK33 | A | G | 0.365 | -0.005 | 0.001 | 4.39E-08 | 0.362 | -0.005 | 0.002 | 6.17E-03 | 0.363 | -0.004 | 0.001 | 2.12E-03 | 0.367 | -0.008 | 0.002 | 1.14E-05 | 0.373 | -0.006 | 0.002 | 1.20E-03 | 0.373 | -0.011 | 0.002 | 1.00E-07 |
| rs2180454* | 14 | 29690513 | 29221307 | LINC02327 | C | T | 0.734 | 0.007 | 0.001 | 1.28E-11 | 0.733 | 0.005 | 0.002 | 1.36E-02 | 0.737 | 0.007 | 0.002 | 4.66E-06 | 0.731 | 0.007 | 0.002 | 3.92E-04 | 0.772 | 0.006 | 0.002 | 4.60E-03 | 0.772 | 0.012 | 0.002 | 1.10E-07 |
| rs3115665* | 6 | 31589264 | 31621487 | PRRC2A | C | G | 0.805 | -0.009 | 0.001 | 6.47E-11 | 0.802 | -0.005 | 0.003 | 3.47E-02 | 0.805 | -0.010 | 0.002 | 5.00E-07 | 0.806 | -0.010 | 0.002 | 3.56E-05 | 0.804 | -0.016 | 0.002 | 2.00E-11 | 0.804 | -0.014 | 0.002 | 7.00E-09 |
| rs815339* | 1 | 190116575 | 190147445 | BRINP3 | A | T | 0.488 | 0.005 | 0.001 | 7.31E-09 | 0.484 | 0.007 | 0.002 | 3.24E-04 | 0.490 | 0.006 | 0.001 | 1.47E-06 | 0.486 | 0.003 | 0.002 | 5.00E-02 | 0.505 | 0.011 | 0.002 | 2.50E-08 | 0.505 | 0.011 | 0.002 | 1.40E-08 |
| rs2076308* | 6 | 50791640 | 50823927 | TFAP2B | C | G | 0.185 | 0.009 | 0.001 | 6.74E-14 | 0.187 | 0.008 | 0.002 | 1.22E-03 | 0.183 | 0.011 | 0.002 | 1.27E-10 | 0.188 | 0.005 | 0.002 | 1.08E-02 | 0.180 | 0.027 | 0.003 | 8.90E-27 | 0.180 | 0.025 | 0.003 | 3.90E-23 |
| rs1902666* | 10 | 88011779 | 86252022 | GRID1 | T | C | 0.885 | 0.008 | 0.002 | 3.92E-08 | 0.888 | 0.009 | 0.003 | 3.09E-03 | 0.887 | 0.008 | 0.002 | 8.11E-05 | 0.884 | 0.009 | 0.003 | 6.90E-04 | 0.870 | 0.007 | 0.003 | 2.10E-02 | 0.870 | 0.006 | 0.003 | 2.90E-02 |
| rs3810291* | 19 | 47569003 | 47065746 | ZC3H4 | A | G | 0.677 | 0.006 | 0.001 | 4.82E-08 | 0.677 | 0.006 | 0.002 | 9.27E-03 | 0.677 | 0.006 | 0.002 | 1.21E-04 | 0.677 | 0.006 | 0.002 | 6.44E-04 | 0.676 | 0.015 | 0.002 | 2.60E-13 | 0.676 | 0.015 | 0.002 | 4.70E-13 |
| rs663129* | 18 | 57838401 | 60171168 | RNU4-17P,MC4R | A | G | 0.254 | 0.010 | 0.001 | 7.80E-20 | 0.254 | 0.008 | 0.002 | 2.54E-04 | 0.253 | 0.011 | 0.002 | 9.89E-12 | 0.251 | 0.012 | 0.002 | 1.87E-09 | 0.233 | 0.035 | 0.002 | 1.20E-55 | 0.233 | 0.038 | 0.002 | 5.00E-61 |
| rs7132908* | 12 | 50263148 | 49869365 | FAIM2 | A | G | 0.398 | 0.008 | 0.001 | 7.33E-18 | 0.396 | 0.009 | 0.002 | 8.39E-06 | 0.398 | 0.008 | 0.001 | 2.18E-09 | 0.402 | 0.008 | 0.002 | 2.20E-06 | 0.384 | 0.031 | 0.002 | 1.80E-56 | 0.384 | 0.019 | 0.002 | 8.20E-21 |
| rs7525548* | 1 | 75001474 | 74535790 | TNNI3K,FPGT-TNNI3K | T | A | 0.567 | -0.008 | 0.001 | 4.00E-19 | 0.564 | -0.009 | 0.002 | 2.86E-06 | 0.567 | -0.012 | 0.001 | 1.96E-18 | 0.568 | -0.003 | 0.002 | 7.54E-02 | 0.563 | -0.032 | 0.002 | 2.20E-58 | 0.563 | -0.013 | 0.002 | 4.00E-11 |
| rs141224959* | 2 | 380055 | 380055 | LINC01865 | A | G | 0.028 | -0.024 | 0.003 | 1.67E-16 | 0.028 | -0.018 | 0.006 | 1.93E-03 | 0.027 | -0.025 | 0.004 | 8.23E-10 | 0.029 | -0.026 | 0.005 | 1.77E-07 | 0.038 | -0.067 | 0.005 | 1.30E-40 | 0.038 | -0.047 | 0.005 | 6.90E-20 |
| rs11030377* | 11 | 28576651 | 28555104 | METTL15 | G | A | 0.498 | -0.006 | 0.001 | 6.57E-11 | 0.503 | 0.005 | 0.002 | 1.02E-02 | 0.503 | 0.006 | 0.001 | 2.60E-06 | 0.500 | 0.007 | 0.002 | 4.64E-05 | 0.502 | 0.009 | 0.002 | 9.50E-07 | 0.502 | 0.005 | 0.002 | 6.60E-03 |
| rs10182181* | 2 | 25150296 | 24927427 | ADCY3,DNAJC27 | G | A | 0.489 | 0.010 | 0.001 | 1.15E-61 | 0.494 | 0.012 | 0.002 | 1.24E-10 | 0.949 | 0.010 | 0.001 | 1.06E-13 | 0.487 | 0.009 | 0.002 | 2.22E-07 | 0.486 | 0.040 | 0.002 | 1.10E-96 | 0.486 | 0.024 | 0.002 | 2.10E-34 |
| rs2094510 | 1 | 177843479 | 177874344 | SEC16B | T | C | 0.227 | -0.013 | 0.001 | 2.78E-30 | 0.227 | -0.014 | 0.002 | 7.01E-10 | 0.229 | -0.011 | 0.002 | 1.31E-11 | 0.222 | -0.012 | 0.002 | 2.79E-09 | 0.790 | -0.048 | 0.002 | 3.40E-92 | 0.790 | -0.032 | 0.002 | 3.60E-40 |
| rs539515 | 1 | 177889025 | 177919890 | LINC01741, SEC16B | C | A | 0.223 | 0.014 | 0.001 | 1.87E-34 | 0.224 | 0.012 | 0.002 | 2.90E-08 | 0.225 | 0.012 | 0.002 | 1.04E-14 | 0.217 | 0.013 | 0.002 | 4.20E-12 | 0.205 | 0.055 | 0.002 | 3.60E-116 | 0.205 | 0.038 | 0.002 | 9.60E-54 |
| rs77165542 | 2 | 430975 | 430975 | LINC01865, LINC01874 | T | C | 0.025 | 0.032 | 0.003 | 4.09E-25 | 0.025 | 0.021 | 0.006 | 5.95E-04 | 0.025 | -0.033 | 0.004 | 1.64E-14 | 0.027 | 0.035 | 0.005 | 9.74E-12 | 0.036 | -0.090 | 0.005 | 1.50E-66 | 0.036 | -0.068 | 0.005 | 3.00E-37 |
| rs688671 | 18 | 57867526 | 60200293 | RNU4-17P,MC4R | G | A | 0.287 | 0.009 | 0.001 | 2.70E-19 | 0.287 | 0.007 | 0.002 | 1.28E-03 | 0.287 | 0.011 | 0.002 | 1.18E-14 | 0.285 | 0.010 | 0.002 | 8.51E-08 | 0.269 | 0.031 | 0.002 | 1.40E-46 | 0.269 | 0.035 | 0.002 | 1.20E-56 |
| rs5017302 | 2 | 631069 | 631069 | LINC01875, TMEM18 | A | G | 0.832 | -0.013 | 0.001 | 2.95E-26 | 0.832 | -0.010 | 0.003 | 5.36E-05 | 0.834 | 0.015 | 0.002 | 1.26E-16 | 0.830 | -0.014 | 0.002 | 4.08E-10 | 0.824 | 0.044 | 0.003 | 2.10E-66 | 0.824 | 0.036 | 0.003 | 4.30E-44 |
| rs55872725 | 16 | 53809123 | 53775211 | FTO | T | C | 0.413 | -0.016 | 0.001 | 4.11E-61 | 0.413 | -0.014 | 0.002 | 2.61E-14 | 0.415 | 0.016 | 0.001 | 6.84E-32 | 0.410 | -0.015 | 0.002 | 5.97E-19 | 0.402 | 0.048 | 0.002 | 6.40E-133 | 0.402 | 0.045 | 0.002 | 4.50E-113 |
| rs7553158 | 1 | 75005238 | 74539554 | TNNI3K,FPGT-TNNI3K | A | G | 0.567 | 0.008 | 0.001 | 4.19E-19 | 0.564 | 0.009 | 0.002 | 2.76E-06 | 0.567 | -0.012 | 0.001 | 1.73E-18 | 0.568 | 0.003 | 0.002 | 7.63E-02 | 0.563 | -0.032 | 0.002 | 3.40E-58 | 0.563 | -0.013 | 0.002 | 3.40E-11 |
| rs12507026 | 4 | 45181334 | 45179317 | THAP12P9, PRDX4P1 | T | A | 0.415 | 0.006 | 0.001 | 8.84E-11 | 0.416 | 0.004 | 0.002 | 6.21E-02 | 0.414 | 0.008 | 0.001 | 9.26E-09 | 0.419 | 0.006 | 0.002 | 1.94E-04 | 0.433 | 0.023 | 0.002 | 3.00E-32 | 0.433 | 0.017 | 0.002 | 2.90E-17 |
| rs4633164 | 8 | 143334191 | 142252830 | TSNARE1 | T | C | 0.397 | -0.006 | 0.001 | 4.93E-09 | 0.398 | -0.003 | 0.002 | 1.50E-01 | 0.398 | 0.008 | 0.001 | 2.17E-08 | 0.394 | -0.003 | 0.002 | 7.79E-02 | 0.395 | 0.003 | 0.002 | 1.00E-01 | 0.395 | 0.008 | 0.002 | 5.10E-05 |
| rs62048402 | 16 | 53803223 | 53769311 | FTO | A | G | 0.412 | -0.016 | 0.001 | 2.26E-61 | 0.413 | -0.014 | 0.002 | 2.07E-14 | 0.415 | -0.016 | 0.001 | 1.23E-31 | 0.409 | 0.015 | 0.002 | 2.89E-19 | 0.402 | 0.048 | 0.002 | 2.00E-133 | 0.402 | 0.045 | 0.002 | 1.70E-113 |
| rs10188334 | 2 | 653874 | 653874 | LINC01875, TMEM18 | T | C | 0.166 | 0.013 | 0.001 | 2.85E-25 | 0.167 | 0.010 | 0.003 | 6.23E-05 | 0.165 | 0.014 | 0.002 | 5.73E-15 | 0.169 | -0.014 | 0.002 | 7.53E-11 | 0.172 | -0.044 | 0.003 | 1.10E-65 | 0.172 | -0.036 | 0.003 | 3.80E-43 |
| rs509325 | 1 | 177894591 | 177925456 | SEC16B | G | T | 0.225 | 0.014 | 0.001 | 8.21E-35 | 0.226 | 0.013 | 0.002 | 1.78E-08 | 0.227 | 0.012 | 0.002 | 1.73E-14 | 0.219 | 0.014 | 0.002 | 4.18E-12 | 0.205 | 0.055 | 0.002 | 7.40E-116 | 0.205 | 0.037 | 0.002 | 6.60E-53 |
| rs571312 | 18 | 57839769 | 60172536 | RNU4-17P,MC4R | A | C | 0.254 | -0.010 | 0.001 | 8.74E-20 | 0.254 | -0.008 | 0.002 | 2.54E-04 | 0.253 | -0.011 | 0.002 | 1.04E-11 | 0.251 | 0.012 | 0.002 | 1.74E-09 | 0.233 | 0.035 | 0.002 | 3.00E-55 | 0.233 | 0.038 | 0.002 | 4.00E-61 |

*Table S4. Genetic correlation coefficients between childhood body size and the four GWAS of BMI in nulliparous women between menarche and <40 years, menarche and <20 years, 20 and <30 years and 30 and <40 years.*

rG – genetic correlation, SE – standard error of rG, P - corresponding p-value

| Phenotype A (age group or life stage and sample) | Phenotype B (age group or life stage and sample) | Sample size in Phenotype A | Sample size in Phenotype B | Total sample size | rG | SE | P |
|---|---|---|---|---|---|---|---|
| Prepubertal body size (UKB) | BMI in nulliparous women between menarche and <40 years (meta-analysis) | 246511 | 56628 | 303139 | 0.76 | 0.02 | 2.13E-223 |
| Prepubertal body size (UKB) | BMI in nulliparous women between menarche and <20 years (meta-analysis) | 246511 | 11396 | 257907 | 0.88 | 0.07 | 3.10E-41 |
| Prepubertal body size (UKB) | BMI in nulliparous women between 20 and <30 years (meta-analysis) | 246511 | 30272 | 276783 | 0.74 | 0.03 | 2.39E-121 |
| Prepubertal body size (UKB) | BMI in nulliparous women between 30 and <40 years (meta-analysis) | 246511 | 16565 | 263076 | 0.71 | 0.05 | 1.67E-47 |
| Prepubertal body size (UKB) | Later life adult body size (UKB) | 246511 | 246511 | 493022 | 0.50 | 0.02 | 1.92E-101 |
| Later life adult body size (UKB) | BMI in nulliparous women between menarche and 40 years (meta-analysis) | 246511 | 56628 | 303139 | 0.85 | 0.02 | 0.00E+00 |
| Later life adult body size (UKB) | BMI in nulliparous women between menarche and <20 years (meta-analysis) | 246511 | 11396 | 257907 | 0.68 | 0.06 | 4.99E-33 |
| Later life adult body size (UKB) | BMI in nulliparous women between 20 and <30 years (meta-analysis) | 246511 | 30272 | 276783 | 0.83 | 0.03 | 5.35E-190 |
| Later life adult body size (UKB) | BMI in nulliparous women between 30 and <40 years (meta-analysis) | 246511 | 16565 | 263076 | 0.95 | 0.05 | 6.41E-75 |
| BMI in nulliparous women between menarche and <20 years (meta-analysis) | BMI in nulliparous women between 20 and <30 years (meta-analysis) | 11396 | 30272 | 41668 | 0.81 | 0.07 | 2.10E-30 |
| BMI in nulliparous women between menarche and <20 years (meta-analysis) | BMI in nulliparous women between 30 and <40 years (meta-analysis) | 11396 | 1656500 | 1667896 | 0.88 | 0.10 | 1.13E-17 |
| BMI in nulliparous women between menarche and <20 years (meta-analysis) | BMI in nulliparous women between menarche and <40 years (meta-analysis) | 11396 | 56628 | 68024 | 0.99 | 0.05 | 2.59E-76 |
| BMI in nulliparous women between 20 and <30 years (meta-analysis) | BMI in nulliparous women between 30 and <40 years (meta-analysis) | 30272 | 16565 | 46837 | 0.99 | 0.06 | 2.24E-53 |
| BMI in nulliparous women between 20 and <30 years (meta-analysis) | BMI in nulliparous women between menarche and <40 years (meta-analysis) | 30272 | 56628 | 86900 | 0.97 | 0.01 | 0.00E+00 |
| BMI in nulliparous women between 30 and <40 years (meta-analysis) | BMI in nulliparous women between menarche and <40 years (meta-analysis) | 16565 | 56628 | 73193 | 1.30 | 0.05 | 8.28E-159 |

*Table S5. Univariable and multivariable Mendelian randomization analyses for BMI in nulliparous women between menarche and <40 years onto overall breast cancer and subtypes.*

nSNP - number of single nucleotide polymorphism identifiers, Beta - effect estimate coefficient for SNP on BMI, SE- standard error of the effect estimate, LCI - lower confidence interval, UCI - upper confidence interval, P - corresponding p-value, IVW - inverse variance weighted, MR - Mendelian ranodomization

| Exposure | Accounting for | Outcome | F-statistic | nSNP | beta | SE | P | Odds ratio | LCI | UCI | Method | MR |
|---|---|---|---|---|---|---|---|---|---|---|---|---|
| BMI in nulliparous women between menarche and <40 years | -- | Overall breast cancer | 63.7 | 22 | -0.27 | 0.06 | 1.27E-05 | 0.76 | 0.67 | 0.86 | IVW | Univariable |
| BMI in nulliparous women between menarche and <40 years | -- | Overall breast cancer | 63.7 | 22 | -0.33 | 0.05 | 9.95E-11 | 0.72 | 0.65 | 0.79 | Weighted median | Univariable |
| BMI in nulliparous women between menarche and <40 years | -- | Overall breast cancer | 63.7 | 22 | -0.66 | 0.14 | 1.31E-04 | 0.52 | 0.39 | 0.68 | MR Egger | Univariable |
| BMI in nulliparous women between menarche and <20 years | -- | Overall breast cancer | 48.4 | 3 | -0.41 | 0.06 | 7.40E-13 | 0.66 | 0.59 | 0.74 | IVW | Univariable |
| BMI in nulliparous women between menarche and <20 years | -- | Overall breast cancer | 48.4 | 3 | -0.42 | 0.06 | 3.84E-11 | 0.66 | 0.58 | 0.75 | Weighted median | Univariable |
| BMI in nulliparous women between menarche and <20 years | -- | Overall breast cancer | 48.4 | 3 | -0.72 | 0.69 | 4.85E-01 | 0.48 | 0.12 | 1.88 | MR Egger | Univariable |
| BMI in nulliparous women between 20 and <30 years | -- | Overall breast cancer | 59.2 | 11 | -0.32 | 0.08 | 4.31E-05 | 0.73 | 0.62 | 0.85 | IVW | Univariable |
| BMI in nulliparous women between 20 and <30 years | -- | Overall breast cancer | 59.2 | 11 | -0.33 | 0.06 | 1.82E-07 | 0.72 | 0.63 | 0.81 | Weighted median | Univariable |
| BMI in nulliparous women between 20 and <30 years | -- | Overall breast cancer | 59.2 | 11 | -0.47 | 0.27 | 1.18E-01 | 0.63 | 0.37 | 1.06 | MR Egger | Univariable |
| BMI in nulliparous women between 30 and <40 years | -- | Overall breast cancer | 49.2 | 5 | -0.37 | 0.10 | 1.70E-04 | 0.69 | 0.57 | 0.84 | IVW | Univariable |
| BMI in nulliparous women between 30 and <40 years | -- | Overall breast cancer | 49.2 | 5 | -0.34 | 0.07 | 1.10E-06 | 0.71 | 0.62 | 0.82 | Weighted median | Univariable |
| BMI in nulliparous women between 30 and <40 years | -- | Overall breast cancer | 49.2 | 5 | 0.02 | 0.38 | 9.62E-01 | 1.02 | 0.48 | 2.15 | MR Egger | Univariable |
| BMI in nulliparous women between menarche and <40 years | Prepubertal body size | Overall breast cancer | 2.9 | 16 | -0.13 | 0.06 | 3.13E-02 | 0.88 | 0.79 | 0.99 | IVW | Multivariable |
| Prepubertal body size | BMI in nulliparous women between menarche and <40 years | Overall breast cancer | 3.6 | 79 | -0.15 | 0.08 | 4.92E-02 | 0.86 | 0.74 | 1.00 | IVW | Multivariable |
| BMI in nulliparous women between menarche and <40 years | Adult body size | Overall breast cancer | 3.6 | 18 | -0.27 | 0.06 | 1.77E-06 | 0.76 | 0.68 | 0.85 | IVW | Multivariable |
| Adult body size | BMI in nulliparous women between menarche and <40 years | Overall breast cancer | 4.9 | 115 | 0.06 | 0.07 | 3.93E-01 | 1.07 | 0.92 | 1.23 | IVW | Multivariable |
| BMI in nulliparous women between menarche and <20 years | Prepubertal body size | Overall breast cancer | 1.2 | 3 | -0.05 | 0.05 | 3.31E-01 | 0.95 | 0.86 | 1.05 | IVW | Multivariable |
| Prepubertal body size | BMI in nulliparous women between menarche and <20 years | Overall breast cancer | 1.9 | 73 | -0.24 | 0.08 | 1.43E-03 | 0.79 | 0.68 | 0.91 | IVW | Multivariable |
| BMI in nulliparous women between menarche and <20 years | Adult body size | Overall breast cancer | 1.8 | 3 | -0.17 | 0.05 | 3.09E-04 | 0.84 | 0.77 | 0.92 | IVW | Multivariable |

| | | | | | | | | | | | | |
|---|---|---|---|---|---|---|---|---|---|---|---|---|
| Adult body size | BMI in nulliparous women between menarche and <20 years | Overall breast cancer | 3.9 | 114 | -0.08 | 0.06 | 2.36E-01 | 0.93 | 0.82 | 1.05 | IVW | Multivariable |
| BMI in nulliparous women between 20 and <30 years | Prepubertal body size | Overall breast cancer | 2.1 | 10 | -0.06 | 0.06 | 3.64E-01 | 0.95 | 0.84 | 1.07 | IVW | Multivariable |
| Prepubertal body size | BMI in nulliparous women between 20 and <30 years | Overall breast cancer | 2.8 | 77 | -0.24 | 0.08 | 2.31E-03 | 0.79 | 0.67 | 0.92 | IVW | Multivariable |
| BMI in nulliparous women between 20 and <30 years | Adult body size | Overall breast cancer | 2.1 | 10 | -0.20 | 0.06 | 1.65E-03 | 0.82 | 0.73 | 0.93 | IVW | Multivariable |
| Adult body size | BMI in nulliparous women between 20 and <30 years | Overall breast cancer | 3.2 | 116 | -0.03 | 0.08 | 7.40E-01 | 0.97 | 0.83 | 1.14 | IVW | Multivariable |
| BMI in nulliparous women between 30 and <40 years | Prepubertal body size | Overall breast cancer | 1.7 | 3 | -0.02 | 0.05 | 6.32E-01 | 0.98 | 0.90 | 1.07 | IVW | Multivariable |
| Prepubertal body size | BMI in nulliparous women between 30 and <40 years | Overall breast cancer | 3.3 | 75 | -0.27 | 0.07 | 3.45E-05 | 0.76 | 0.67 | 0.87 | IVW | Multivariable |
| BMI in nulliparous women between 30 and <40 years | Adult body size | Overall breast cancer | 1.8 | 3 | -0.18 | 0.05 | 2.62E-04 | 0.84 | 0.76 | 0.92 | IVW | Multivariable |
| Adult body size | BMI in nulliparous women between 30 and <40 years | Overall breast cancer | 4.5 | 114 | -0.07 | 0.07 | 3.17E-01 | 0.94 | 0.82 | 1.06 | IVW | Multivariable |
| BMI in nulliparous women between menarche and <40 years | -- | ER+ breast cancer | 63.7 | 22 | -0.27 | 0.06 | 5.18E-06 | 0.76 | 0.68 | 0.86 | IVW | Univariable |
| BMI in nulliparous women between menarche and <40 years | -- | ER+ breast cancer | 63.7 | 22 | -0.24 | 0.06 | 7.41E-05 | 0.79 | 0.70 | 0.89 | Weighted median | Univariable |
| BMI in nulliparous women between menarche and <40 years | -- | ER+ breast cancer | 63.7 | 22 | -0.56 | 0.14 | 8.42E-04 | 0.57 | 0.43 | 0.75 | MR Egger | Univariable |
| BMI in nulliparous women between menarche and <20 years | -- | ER+ breast cancer | 48.4 | 3 | -0.40 | 0.06 | 1.66E-11 | 0.67 | 0.60 | 0.75 | IVW | Univariable |
| BMI in nulliparous women between menarche and <20 years | -- | ER+ breast cancer | 48.4 | 3 | -0.41 | 0.07 | 8.71E-09 | 0.66 | 0.57 | 0.76 | Weighted median | Univariable |
| BMI in nulliparous women between menarche and <20 years | -- | ER+ breast cancer | 48.4 | 3 | -0.69 | 0.73 | 5.18E-01 | 0.50 | 0.12 | 2.09 | MR Egger | Univariable |
| BMI in nulliparous women between 20 and <30 years | -- | ER+ breast cancer | 58.3 | 11 | -0.33 | 0.07 | 5.61E-06 | 0.72 | 0.62 | 0.83 | IVW | Univariable |
| BMI in nulliparous women between 20 and <30 years | -- | ER+ breast cancer | 58.3 | 11 | -0.31 | 0.06 | 3.70E-07 | 0.73 | 0.65 | 0.83 | Weighted median | Univariable |
| BMI in nulliparous women between 20 and <30 years | -- | ER+ breast cancer | 58.3 | 11 | -0.50 | 0.25 | 7.73E-02 | 0.61 | 0.37 | 0.99 | MR Egger | Univariable |
| BMI in nulliparous women between 30 and <40 years | -- | ER+ breast cancer | 49.2 | 5 | -0.36 | 0.09 | 3.64E-05 | 0.70 | 0.59 | 0.83 | IVW | Univariable |
| BMI in nulliparous women between 30 and <40 years | -- | ER+ breast cancer | 49.2 | 5 | -0.28 | 0.07 | 1.26E-04 | 0.75 | 0.65 | 0.87 | Weighted median | Univariable |
| BMI in nulliparous women between 30 and <40 years | -- | ER+ breast cancer | 49.2 | 5 | -0.29 | 0.40 | 5.16E-01 | 0.75 | 0.34 | 1.63 | MR Egger | Univariable |
| BMI in nulliparous women between menarche and <40 years | Prepubertal body size | ER+ breast cancer | 2.9 | 16 | -0.14 | 0.06 | 2.54E-02 | 0.87 | 0.76 | 0.98 | IVW | Multivariable |
| Prepubertal body size | BMI in nulliparous women between menarche and <40 years | ER+ breast cancer | 3.6 | 79 | -0.09 | 0.08 | 2.72E-01 | 0.91 | 0.77 | 1.08 | IVW | Multivariable |

| | | | | | | | | | | | | |
|---|---|---|---|---|---|---|---|---|---|---|---|---|
| BMI in nulliparous women between menarche and <40 years | Adult body size | ER+ breast cancer | 3.7 | 18 | -0.19 | 0.04 | 5.09E-06 | 0.82 | 0.76 | 0.90 | IVW | Multivariable |
| Adult body size | BMI in nulliparous women between menarche and <40 years | ER+ breast cancer | 5.6 | 114 | 0.08 | 0.08 | 3.27E-01 | 1.09 | 0.92 | 1.28 | IVW | Multivariable |
| BMI in nulliparous women between menarche and <20 years | Prepubertal body size | ER+ breast cancer | 1.2 | 3 | -0.06 | 0.06 | 2.82E-01 | 0.94 | 0.84 | 1.05 | IVW | Multivariable |
| Prepubertal body size | BMI in nulliparous women between menarche and <20 years | ER+ breast cancer | 1.9 | 73 | -0.18 | 0.08 | 3.19E-02 | 0.84 | 0.71 | 0.98 | IVW | Multivariable |
| BMI in nulliparous women between menarche and <20 years | Adult body size | ER+ breast cancer | 1.8 | 3 | -0.19 | 0.05 | 3.77E-04 | 0.83 | 0.74 | 0.92 | IVW | Multivariable |
| Adult body size | BMI in nulliparous women between menarche and <20 years | ER+ breast cancer | 3.9 | 113 | -0.05 | 0.07 | 4.61E-01 | 0.95 | 0.82 | 1.09 | IVW | Multivariable |
| BMI in nulliparous women between 20 and <30 years | Prepubertal body size | ER+ breast cancer | 2.2 | 10 | -0.04 | 0.07 | 5.49E-01 | 0.96 | 0.84 | 1.09 | IVW | Multivariable |
| Prepubertal body size | BMI in nulliparous women between 20 and <30 years | ER+ breast cancer | 4.2 | 77 | -0.21 | 0.09 | 1.32E-02 | 0.81 | 0.68 | 0.96 | IVW | Multivariable |
| BMI in nulliparous women between 20 and <30 years | Adult body size | ER+ breast cancer | 2.2 | 10 | -0.21 | 0.07 | 2.66E-03 | 0.81 | 0.70 | 0.93 | IVW | Multivariable |
| Adult body size | BMI in nulliparous women between 20 and <30 years | ER+ breast cancer | 4.5 | 115 | -0.01 | 0.09 | 9.10E-01 | 0.99 | 0.83 | 1.19 | IVW | Multivariable |
| BMI in nulliparous women between 30 and <40 years | Prepubertal body size | ER+ breast cancer | 1.7 | 3 | -0.02 | 0.05 | 6.51E-01 | 0.98 | 0.89 | 1.08 | IVW | Multivariable |
| Prepubertal body size | BMI in nulliparous women between 30 and <40 years | ER+ breast cancer | 3.3 | 75 | -0.22 | 0.07 | 2.33E-03 | 0.80 | 0.69 | 0.92 | IVW | Multivariable |
| BMI in nulliparous women between 30 and <40 years | Adult body size | ER+ breast cancer | 1.8 | 3 | -0.21 | 0.05 | 7.92E-05 | 0.81 | 0.73 | 0.90 | IVW | Multivariable |
| Adult body size | BMI in nulliparous women between 30 and <40 years | ER+ breast cancer | 4.4 | 113 | -0.03 | 0.07 | 7.21E-01 | 0.97 | 0.85 | 1.12 | IVW | Multivariable |
| BMI in nulliparous women between menarche and <40 years | -- | ER- breast cancer | 63.7 | 22 | -0.42 | 0.10 | 1.59E-05 | 0.66 | 0.55 | 0.80 | IVW | Univariable |
| BMI in nulliparous women between menarche and <40 years | -- | ER- breast cancer | 63.7 | 22 | -0.37 | 0.10 | 3.55E-04 | 0.69 | 0.56 | 0.85 | Weighted median | Univariable |
| BMI in nulliparous women between menarche and <40 years | -- | ER- breast cancer | 63.7 | 22 | -0.73 | 0.25 | 7.74E-03 | 0.48 | 0.30 | 0.78 | MR Egger | Univariable |
| BMI in nulliparous women between menarche and <20 years | -- | ER- breast cancer | 48.4 | 3 | -0.54 | 0.14 | 8.30E-05 | 0.58 | 0.45 | 0.76 | IVW | Univariable |
| BMI in nulliparous women between menarche and <20 years | -- | ER- breast cancer | 48.4 | 3 | -0.64 | 0.11 | 2.17E-09 | 0.53 | 0.43 | 0.65 | Weighted median | Univariable |
| BMI in nulliparous women between menarche and <20 years | -- | ER- breast cancer | 48.4 | 3 | -0.74 | 1.80 | 7.54E-01 | 0.48 | 0.01 | 16.46 | MR Egger | Univariable |
| BMI in nulliparous women between 20 and <30 years | -- | ER- breast cancer | 58.3 | 11 | -0.50 | 0.10 | 1.33E-07 | 0.60 | 0.50 | 0.73 | IVW | Univariable |
| BMI in nulliparous women between 20 and <30 years | -- | ER- breast cancer | 58.3 | 11 | -0.43 | 0.10 | 3.43E-05 | 0.65 | 0.53 | 0.80 | Weighted median | Univariable |
| BMI in nulliparous women between 20 and <30 years | -- | ER- breast cancer | 58.3 | 11 | -0.57 | 0.34 | 1.24E-01 | 0.56 | 0.29 | 1.09 | MR Egger | Univariable |

| | | | | | | | | | | | | |
|---|---|---|---|---|---|---|---|---|---|---|---|---|
| BMI in nulliparous women between 30 and <40 years | -- | ER- breast cancer | 49.2 | 5 | -0.48 | 0.15 | 1.79E-03 | 0.62 | 0.46 | 0.84 | IVW | Univariable |
| BMI in nulliparous women between 30 and <40 years | -- | ER- breast cancer | 49.2 | 5 | -0.33 | 0.14 | 1.65E-02 | 0.72 | 0.55 | 0.94 | Weighted median | Univariable |
| BMI in nulliparous women between 30 and <40 years | -- | ER- breast cancer | 49.2 | 5 | -0.08 | 0.67 | 9.11E-01 | 0.92 | 0.25 | 3.40 | MR Egger | Univariable |
| BMI in nulliparous women between menarche and <40 years | Prepubertal body size | ER- breast cancer | 3.0 | 16 | -0.15 | 0.09 | 8.05E-02 | 0.86 | 0.72 | 1.02 | IVW | Multivariable |
| Prepubertal body size | BMI in nulliparous women between menarche and <40 years | ER- breast cancer | 4.4 | 79 | -0.21 | 0.11 | 6.97E-02 | 0.81 | 0.65 | 1.02 | IVW | Multivariable |
| BMI in nulliparous women between menarche and <40 years | Adult body size | ER- breast cancer | 3.7 | 18 | -0.24 | 0.06 | 4.24E-05 | 0.79 | 0.70 | 0.88 | IVW | Multivariable |
| Adult body size | BMI in nulliparous women between menarche and <40 years | ER- breast cancer | 5.6 | 114 | 0.15 | 0.11 | 1.87E-01 | 1.16 | 0.93 | 1.45 | IVW | Multivariable |
| BMI in nulliparous women between menarche and <20 years | Prepubertal body size | ER- breast cancer | 1.2 | 3 | -0.04 | 0.08 | 5.77E-01 | 0.96 | 0.82 | 1.12 | IVW | Multivariable |
| Prepubertal body size | BMI in nulliparous women between menarche and <20 years | ER- breast cancer | 1.9 | 73 | -0.32 | 0.11 | 4.82E-03 | 0.72 | 0.58 | 0.91 | IVW | Multivariable |
| BMI in nulliparous women between menarche and <20 years | Adult body size | ER- breast cancer | 1.8 | 3 | -0.27 | 0.07 | 2.92E-04 | 0.77 | 0.66 | 0.89 | IVW | Multivariable |
| Adult body size | BMI in nulliparous women between menarche and <20 years | ER- breast cancer | 3.9 | 113 | 0.03 | 0.10 | 7.53E-01 | 1.03 | 0.85 | 1.25 | IVW | Multivariable |
| BMI in nulliparous women between 20 and <30 years | Prepubertal body size | ER- breast cancer | 2.2 | 10 | -0.17 | 0.09 | 5.79E-02 | 0.84 | 0.70 | 1.01 | IVW | Multivariable |
| Prepubertal body size | BMI in nulliparous women between 20 and <30 years | ER- breast cancer | 4.2 | 77 | -0.21 | 0.12 | 8.44E-02 | 0.81 | 0.65 | 1.03 | IVW | Multivariable |
| BMI in nulliparous women between 20 and <30 years | Adult body size | ER- breast cancer | 2.2 | 10 | -0.38 | 0.09 | 5.93E-05 | 0.68 | 0.57 | 0.82 | IVW | Multivariable |
| Adult body size | BMI in nulliparous women between 20 and <30 years | ER- breast cancer | 4.5 | 115 | 0.18 | 0.12 | 1.58E-01 | 1.19 | 0.93 | 1.52 | IVW | Multivariable |
| BMI in nulliparous women between 30 and <40 years | Prepubertal body size | ER- breast cancer | 1.7 | 3 | 0.04 | 0.07 | 5.85E-01 | 1.04 | 0.91 | 1.19 | IVW | Multivariable |
| Prepubertal body size | BMI in nulliparous women between 30 and <40 years | ER- breast cancer | 3.3 | 75 | -0.42 | 0.10 | 4.26E-05 | 0.66 | 0.54 | 0.81 | IVW | Multivariable |
| BMI in nulliparous women between 30 and <40 years | Adult body size | ER- breast cancer | 1.8 | 3 | -0.15 | 0.08 | 4.87E-02 | 0.86 | 0.74 | 1.00 | IVW | Multivariable |
| Adult body size | BMI in nulliparous women between 30 and <40 years | ER- breast cancer | 4.4 | 113 | -0.07 | 0.11 | 5.14E-01 | 0.93 | 0.76 | 1.15 | IVW | Multivariable |
| BMI in nulliparous women between menarche and <40 years | -- | Luminal A subtype | 63.7 | 22 | -0.24 | 0.07 | 2.92E-04 | 0.79 | 0.69 | 0.90 | IVW | Univariable |
| BMI in nulliparous women between menarche and <40 years | -- | Luminal A subtype | 63.7 | 22 | -0.22 | 0.07 | 5.80E-04 | 0.80 | 0.70 | 0.91 | Weighted median | Univariable |
| BMI in nulliparous women between menarche and <40 years | -- | Luminal A subtype | 63.7 | 22 | -0.65 | 0.14 | 2.12E-04 | 0.52 | 0.40 | 0.69 | MR Egger | Univariable |
| BMI in nulliparous women between menarche and <20 years | -- | Luminal A subtype | 48.4 | 3 | -0.39 | 0.07 | 6.08E-09 | 0.67 | 0.59 | 0.77 | IVW | Univariable |

| | | | | | | | | | | | | |
|---|---|---|---|---|---|---|---|---|---|---|---|---|
| BMI in nulliparous women between menarche and <20 years | -- | Luminal A subtype | 48.4 | 3 | -0.39 | 0.08 | 4.75E-07 | 0.68 | 0.58 | 0.79 | Weighted median | Univariable |
| BMI in nulliparous women between menarche and <20 years | -- | Luminal A subtype | 48.4 | 3 | -0.90 | 0.75 | 4.41E-01 | 0.41 | 0.09 | 1.76 | MR Egger | Univariable |
| BMI in nulliparous women between 20 and <30 years | -- | Luminal A subtype | 59.2 | 11 | -0.27 | 0.09 | 1.41E-03 | 0.76 | 0.64 | 0.90 | IVW | Univariable |
| BMI in nulliparous women between 20 and <30 years | -- | Luminal A subtype | 59.2 | 11 | -0.15 | 0.07 | 2.43E-02 | 0.86 | 0.75 | 0.98 | Weighted median | Univariable |
| BMI in nulliparous women between 20 and <30 years | -- | Luminal A subtype | 59.2 | 11 | -0.52 | 0.29 | 1.06E-01 | 0.60 | 0.34 | 1.05 | MR Egger | Univariable |
| BMI in nulliparous women between 30 and <40 years | -- | Luminal A subtype | 49.2 | 5 | -0.34 | 0.12 | 4.10E-03 | 0.71 | 0.56 | 0.90 | IVW | Univariable |
| BMI in nulliparous women between 30 and <40 years | -- | Luminal A subtype | 49.2 | 5 | -0.22 | 0.08 | 7.92E-03 | 0.80 | 0.68 | 0.94 | Weighted median | Univariable |
| BMI in nulliparous women between 30 and <40 years | -- | Luminal A subtype | 49.2 | 5 | -0.25 | 0.53 | 6.71E-01 | 0.78 | 0.27 | 2.21 | MR Egger | Univariable |
| BMI in nulliparous women between menarche and <40 years | Prepubertal body size | Luminal A subtype | 2.9 | 16 | -0.07 | 0.07 | 3.58E-01 | 0.94 | 0.81 | 1.08 | IVW | Multivariable |
| Prepubertal body size | BMI in nulliparous women between menarche and <40 years | Luminal A subtype | 3.6 | 79 | -0.16 | 0.09 | 9.74E-02 | 0.86 | 0.71 | 1.03 | IVW | Multivariable |
| BMI in nulliparous women between menarche and <40 years | Adult body size | Luminal A subtype | 3.4 | 18 | -0.28 | 0.07 | 8.04E-05 | 0.76 | 0.66 | 0.87 | IVW | Multivariable |
| Adult body size | BMI in nulliparous women between menarche and <40 years | Luminal A subtype | 4.8 | 115 | 0.12 | 0.09 | 1.81E-01 | 1.13 | 0.94 | 1.36 | IVW | Multivariable |
| BMI in nulliparous women between menarche and <20 years | Prepubertal body size | Luminal A subtype | 1.2 | 3 | -0.02 | 0.06 | 7.75E-01 | 0.98 | 0.87 | 1.11 | IVW | Multivariable |
| Prepubertal body size | BMI in nulliparous women between menarche and <20 years | Luminal A subtype | 1.7 | 73 | -0.23 | 0.09 | 1.18E-02 | 0.79 | 0.66 | 0.95 | IVW | Multivariable |
| BMI in nulliparous women between menarche and <20 years | Adult body size | Luminal A subtype | 1.7 | 3 | -0.21 | 0.06 | 2.66E-04 | 0.81 | 0.72 | 0.91 | IVW | Multivariable |
| Adult body size | BMI in nulliparous women between menarche and <20 years | Luminal A subtype | 3.2 | 114 | 0.01 | 0.08 | 9.15E-01 | 1.01 | 0.86 | 1.18 | IVW | Multivariable |
| BMI in nulliparous women between 20 and <30 years | Prepubertal body size | Luminal A subtype | 2.1 | 10 | 0.04 | 0.07 | 6.11E-01 | 1.04 | 0.90 | 1.20 | IVW | Multivariable |
| Prepubertal body size | BMI in nulliparous women between 20 and <30 years | Luminal A subtype | 2.8 | 77 | -0.28 | 0.10 | 3.11E-03 | 0.75 | 0.62 | 0.91 | IVW | Multivariable |
| BMI in nulliparous women between 20 and <30 years | Adult body size | Luminal A subtype | 2.1 | 10 | -0.18 | 0.08 | 1.94E-02 | 0.83 | 0.72 | 0.97 | IVW | Multivariable |
| Adult body size | BMI in nulliparous women between 20 and <30 years | Luminal A subtype | 3.2 | 116 | 0.01 | 0.10 | 9.38E-01 | 1.01 | 0.83 | 1.23 | IVW | Multivariable |
| BMI in nulliparous women between 30 and <40 years | Prepubertal body size | Luminal A subtype | 1.7 | 3 | 0.04 | 0.05 | 4.13E-01 | 1.05 | 0.94 | 1.16 | IVW | Multivariable |
| Prepubertal body size | BMI in nulliparous women between 30 and <40 years | Luminal A subtype | 3.2 | 75 | -0.29 | 0.08 | 2.98E-04 | 0.75 | 0.64 | 0.88 | IVW | Multivariable |
| BMI in nulliparous women between 30 and <40 years | Adult body size | Luminal A subtype | 1.8 | 3 | -0.18 | 0.06 | 3.15E-03 | 0.84 | 0.75 | 0.94 | IVW | Multivariable |

| | | | | | | | | | | | | |
|---|---|---|---|---|---|---|---|---|---|---|---|---|
| Adult body size | BMI in nulliparous women between 30 and <40 years | Luminal A subtype | 4.3 | 114 | -0.01 | 0.05 | 8.59E-01 | 0.99 | 0.89 | 1.09 | IVW | Multivariable |
| BMI in nulliparous women between menarche and <40 years | -- | Luminal B1 subtype | 63.7 | 22 | -0.22 | 0.07 | 1.54E-03 | 0.80 | 0.70 | 0.92 | IVW | Univariable |
| BMI in nulliparous women between menarche and <40 years | -- | Luminal B1 subtype | 63.7 | 22 | -0.26 | 0.10 | 8.88E-03 | 0.77 | 0.63 | 0.94 | Weighted median | Univariable |
| BMI in nulliparous women between menarche and <40 years | -- | Luminal B1 subtype | 63.7 | 22 | -0.64 | 0.18 | 1.80E-03 | 0.53 | 0.37 | 0.75 | MR Egger | Univariable |
| BMI in nulliparous women between menarche and <20 years | -- | Luminal B1 subtype | 48.4 | 3 | -0.34 | 0.13 | 6.22E-03 | 0.71 | 0.55 | 0.91 | IVW | Univariable |
| BMI in nulliparous women between menarche and <20 years | -- | Luminal B1 subtype | 48.4 | 3 | -0.27 | 0.12 | 2.85E-02 | 0.77 | 0.60 | 0.97 | Weighted median | Univariable |
| BMI in nulliparous women between menarche and <20 years | -- | Luminal B1 subtype | 48.4 | 3 | -1.88 | 0.91 | 2.88E-01 | 0.15 | 0.03 | 0.91 | MR Egger | Univariable |
| BMI in nulliparous women between <20 years and 30 | -- | Luminal B1 subtype | 59.2 | 11 | -0.29 | 0.08 | 1.25E-04 | 0.75 | 0.64 | 0.87 | IVW | Univariable |
| BMI in nulliparous women between <20 years and 30 | -- | Luminal B1 subtype | 59.2 | 11 | -0.29 | 0.11 | 9.21E-03 | 0.75 | 0.60 | 0.93 | Weighted median | Univariable |
| BMI in nulliparous women between <20 years and 30 | -- | Luminal B1 subtype | 59.2 | 11 | -0.61 | 0.24 | 3.15E-02 | 0.54 | 0.34 | 0.87 | MR Egger | Univariable |
| BMI in nulliparous women between 30 and <40 years | -- | Luminal B1 subtype | 49.2 | 5 | -0.37 | 0.09 | 2.87E-05 | 0.69 | 0.58 | 0.82 | IVW | Univariable |
| BMI in nulliparous women between 30 and <40 years | -- | Luminal B1 subtype | 49.2 | 5 | -0.31 | 0.11 | 6.73E-03 | 0.73 | 0.59 | 0.92 | Weighted median | Univariable |
| BMI in nulliparous women between 30 and <40 years | -- | Luminal B1 subtype | 49.2 | 5 | -0.38 | 0.34 | 3.41E-01 | 0.68 | 0.35 | 1.32 | MR Egger | Univariable |
| BMI in nulliparous women between menarche and <40 years | Prepubertal body size | Luminal B1 subtype | 2.9 | 16 | -0.11 | 0.09 | 2.48E-01 | 0.90 | 0.74 | 1.08 | IVW | Multivariable |
| Prepubertal body size | BMI in nulliparous women between menarche and <40 years | Luminal B1 subtype | 3.6 | 79 | -0.21 | 0.12 | 9.27E-02 | 0.81 | 0.64 | 1.04 | IVW | Multivariable |
| BMI in nulliparous women between menarche and <40 years | Adult body size | Luminal B1 subtype | 3.4 | 18 | -0.25 | 0.10 | 1.21E-02 | 0.78 | 0.64 | 0.95 | IVW | Multivariable |
| Adult body size | BMI in nulliparous women between menarche and <40 years | Luminal B1 subtype | 4.8 | 115 | 0.00 | 0.13 | 9.88E-01 | 1.00 | 0.78 | 1.30 | IVW | Multivariable |
| BMI in nulliparous women between menarche and <20 years | Prepubertal body size | Luminal B1 subtype | 1.2 | 3 | -0.12 | 0.09 | 1.61E-01 | 0.88 | 0.75 | 1.05 | IVW | Multivariable |
| Prepubertal body size | BMI in nulliparous women between menarche and <20 years | Luminal B1 subtype | 1.7 | 73 | -0.20 | 0.13 | 1.14E-01 | 0.82 | 0.64 | 1.05 | IVW | Multivariable |
| BMI in nulliparous women between menarche and <20 years | Adult body size | Luminal B1 subtype | 1.7 | 3 | -0.11 | 0.08 | 1.83E-01 | 0.89 | 0.76 | 1.05 | IVW | Multivariable |
| Adult body size | BMI in nulliparous women between menarche and <20 years | Luminal B1 subtype | 3.2 | 114 | -0.18 | 0.11 | 1.13E-01 | 0.84 | 0.67 | 1.04 | IVW | Multivariable |
| BMI in nulliparous women between <20 years and 30 | Prepubertal body size | Luminal B1 subtype | 2.1 | 10 | -0.09 | 0.10 | 3.62E-01 | 0.91 | 0.75 | 1.11 | IVW | Multivariable |

| | | | | | | | | | | | | |
|---|---|---|---|---|---|---|---|---|---|---|---|---|
| Prepubertal body size | BMI in nulliparous women between 20 and <30 years | Luminal B1 subtype | 2.8 | 77 | -0.24 | 0.13 | 6.22E-02 | 0.79 | 0.61 | 1.01 | IVW | Multivariable |
| BMI in nulliparous women between <20 years and 30 | Adult body size | Luminal B1 subtype | 2.1 | 10 | -0.20 | 0.11 | 6.54E-02 | 0.82 | 0.67 | 1.01 | IVW | Multivariable |
| Adult body size | BMI in nulliparous women between 20 and <30 years | Luminal B1 subtype | 3.2 | 116 | -0.08 | 0.14 | 5.84E-01 | 0.93 | 0.70 | 1.22 | IVW | Multivariable |
| BMI in nulliparous women between 30 and <40 years | Prepubertal body size | Luminal B1 subtype | 1.7 | 3 | -0.10 | 0.07 | 1.59E-01 | 0.90 | 0.78 | 1.04 | IVW | Multivariable |
| Prepubertal body size | BMI in nulliparous women between 30 and <40 years | Luminal B1 subtype | 3.2 | 75 | -0.23 | 0.11 | 3.18E-02 | 0.79 | 0.64 | 0.98 | IVW | Multivariable |
| BMI in nulliparous women between 30 and <40 years | Adult body size | Luminal B1 subtype | 1.8 | 3 | -0.26 | 0.08 | 1.46E-03 | 0.77 | 0.65 | 0.90 | IVW | Multivariable |
| Adult body size | BMI in nulliparous women between 30 and <40 years | Luminal B1 subtype | 4.3 | 114 | -0.03 | 0.11 | 7.75E-01 | 0.97 | 0.78 | 1.21 | IVW | Multivariable |
| BMI in nulliparous women between menarche and <40 years | -- | Luminal B2 subtype | 63.7 | 22 | -0.29 | 0.12 | 1.91E-02 | 0.75 | 0.59 | 0.95 | IVW | Univariable |
| BMI in nulliparous women between menarche and <40 years | -- | Luminal B2 subtype | 63.7 | 22 | -0.45 | 0.13 | 8.94E-04 | 0.64 | 0.49 | 0.83 | Weighted median | Univariable |
| BMI in nulliparous women between menarche and <40 years | -- | Luminal B2 subtype | 63.7 | 22 | -0.74 | 0.31 | 2.63E-02 | 0.48 | 0.26 | 0.87 | MR Egger | Univariable |
| BMI in nulliparous women between menarche and <20 years | -- | Luminal B2 subtype | 48.4 | 3 | -0.60 | 0.11 | 1.49E-07 | 0.55 | 0.44 | 0.69 | IVW | Univariable |
| BMI in nulliparous women between menarche and <20 years | -- | Luminal B2 subtype | 48.4 | 3 | -0.55 | 0.15 | 1.74E-04 | 0.58 | 0.43 | 0.77 | Weighted median | Univariable |
| BMI in nulliparous women between menarche and <20 years | -- | Luminal B2 subtype | 48.4 | 3 | -0.52 | 1.07 | 7.11E-01 | 0.59 | 0.07 | 4.83 | MR Egger | Univariable |
| BMI in nulliparous women between 20 and <30 years | -- | Luminal B2 subtype | 59.2 | 11 | -0.35 | 0.15 | 1.67E-02 | 0.71 | 0.53 | 0.94 | IVW | Univariable |
| BMI in nulliparous women between 20 and <30 years | -- | Luminal B2 subtype | 59.2 | 11 | -0.42 | 0.14 | 3.38E-03 | 0.66 | 0.50 | 0.87 | Weighted median | Univariable |
| BMI in nulliparous women between 20 and <30 years | -- | Luminal B2 subtype | 59.2 | 11 | -0.67 | 0.50 | 2.08E-01 | 0.51 | 0.19 | 1.35 | MR Egger | Univariable |
| BMI in nulliparous women between 30 and <40 years | -- | Luminal B2 subtype | 49.2 | 5 | -0.42 | 0.16 | 9.40E-03 | 0.66 | 0.48 | 0.90 | IVW | Univariable |
| BMI in nulliparous women between 30 and <40 years | -- | Luminal B2 subtype | 49.2 | 5 | -0.46 | 0.15 | 2.29E-03 | 0.63 | 0.47 | 0.85 | Weighted median | Univariable |
| BMI in nulliparous women between 30 and <40 years | -- | Luminal B2 subtype | 49.2 | 5 | 0.28 | 0.58 | 6.65E-01 | 1.32 | 0.42 | 4.13 | MR Egger | Univariable |
| BMI in nulliparous women between menarche and <40 years | Prepubertal body size | Luminal B2 subtype | 2.9 | 16 | -0.29 | 0.11 | 7.99E-03 | 0.75 | 0.60 | 0.93 | IVW | Multivariable |
| Prepubertal body size | BMI in nulliparous women between menarche and <40 years | Luminal B2 subtype | 3.6 | 79 | 0.07 | 0.15 | 6.13E-01 | 1.08 | 0.81 | 1.43 | IVW | Multivariable |
| BMI in nulliparous women between menarche and <40 years | Adult body size | Luminal B2 subtype | 3.4 | 18 | -0.36 | 0.11 | 9.49E-04 | 0.70 | 0.57 | 0.87 | IVW | Multivariable |
| Adult body size | BMI in nulliparous women between menarche and <40 years | Luminal B2 subtype | 4.8 | 115 | 0.17 | 0.14 | 2.37E-01 | 1.18 | 0.90 | 1.55 | IVW | Multivariable |

| | | | | | | | | | | | | |
|---|---|---|---|---|---|---|---|---|---|---|---|---|
| BMI in nulliparous women between menarche and <20 years | Prepubertal body size | Luminal B2 subtype | 1.2 | 3 | -0.15 | 0.10 | 1.38E-01 | 0.86 | 0.71 | 1.05 | IVW | Multivariable |
| Prepubertal body size | BMI in nulliparous women between menarche and <20 years | Luminal B2 subtype | 1.7 | 73 | -0.07 | 0.15 | 6.25E-01 | 0.93 | 0.70 | 1.24 | IVW | Multivariable |
| BMI in nulliparous women between menarche and <20 years | Adult body size | Luminal B2 subtype | 1.7 | 3 | -0.19 | 0.09 | 3.38E-02 | 0.83 | 0.70 | 0.99 | IVW | Multivariable |
| Adult body size | BMI in nulliparous women between menarche and <20 years | Luminal B2 subtype | 3.2 | 114 | -0.07 | 0.12 | 5.76E-01 | 0.94 | 0.74 | 1.18 | IVW | Multivariable |
| BMI in nulliparous women between 20 and <30 years | Prepubertal body size | Luminal B2 subtype | 2.1 | 10 | -0.15 | 0.12 | 2.04E-01 | 0.86 | 0.69 | 1.08 | IVW | Multivariable |
| Prepubertal body size | BMI in nulliparous women between 20 and <30 years | Luminal B2 subtype | 2.8 | 77 | -0.06 | 0.15 | 6.68E-01 | 0.94 | 0.70 | 1.26 | IVW | Multivariable |
| BMI in nulliparous women between 20 and <30 years | Adult body size | Luminal B2 subtype | 2.1 | 10 | -0.18 | 0.12 | 1.29E-01 | 0.84 | 0.67 | 1.05 | IVW | Multivariable |
| Adult body size | BMI in nulliparous women between 20 and <30 years | Luminal B2 subtype | 3.2 | 116 | -0.03 | 0.15 | 8.27E-01 | 0.97 | 0.72 | 1.30 | IVW | Multivariable |
| BMI in nulliparous women between 30 and <40 years | Prepubertal body size | Luminal B2 subtype | 1.7 | 3 | -0.10 | 0.09 | 2.36E-01 | 0.90 | 0.76 | 1.07 | IVW | Multivariable |
| Prepubertal body size | BMI in nulliparous women between 30 and <40 years | Luminal B2 subtype | 3.2 | 75 | -0.12 | 0.13 | 3.54E-01 | 0.89 | 0.69 | 1.14 | IVW | Multivariable |
| BMI in nulliparous women between 30 and <40 years | Adult body size | Luminal B2 subtype | 1.8 | 3 | -0.22 | 0.09 | 1.47E-02 | 0.81 | 0.68 | 0.96 | IVW | Multivariable |
| Adult body size | BMI in nulliparous women between 30 and <40 years | Luminal B2 subtype | 4.3 | 114 | -0.02 | 0.12 | 8.37E-01 | 0.98 | 0.77 | 1.23 | IVW | Multivariable |
| BMI in nulliparous women between menarche and <40 years | -- | HER2-enriched subtype | 63.7 | 22 | -0.27 | 0.16 | 8.07E-02 | 0.76 | 0.56 | 1.03 | IVW | Univariable |
| BMI in nulliparous women between menarche and <40 years | -- | HER2-enriched subtype | 63.7 | 22 | -0.28 | 0.19 | 1.33E-01 | 0.75 | 0.52 | 1.09 | Weighted median | Univariable |
| BMI in nulliparous women between menarche and <40 years | -- | HER2-enriched subtype | 63.7 | 22 | -0.99 | 0.38 | 1.56E-02 | 0.37 | 0.18 | 0.77 | MR Egger | Univariable |
| BMI in nulliparous women between menarche and <20 years | -- | HER2-enriched subtype | 48.4 | 3 | -0.58 | 0.17 | 4.69E-04 | 0.56 | 0.40 | 0.78 | IVW | Univariable |
| BMI in nulliparous women between menarche and <20 years | -- | HER2-enriched subtype | 48.4 | 3 | -0.58 | 0.21 | 5.82E-03 | 0.56 | 0.37 | 0.84 | Weighted median | Univariable |
| BMI in nulliparous women between menarche and <20 years | -- | HER2-enriched subtype | 48.4 | 3 | 0.27 | 1.60 | 8.95E-01 | 1.30 | 0.06 | 30.08 | MR Egger | Univariable |
| BMI in nulliparous women between <20 years and 30 | -- | HER2-enriched subtype | 59.2 | 11 | -0.44 | 0.17 | 9.08E-03 | 0.01 | 0.46 | 0.90 | IVW | Univariable |
| BMI in nulliparous women between <20 years and 30 | -- | HER2-enriched subtype | 59.2 | 11 | -0.39 | 0.20 | 4.42E-02 | 0.67 | 0.46 | 0.99 | Weighted median | Univariable |
| BMI in nulliparous women between <20 years and 30 | -- | HER2-enriched subtype | 59.2 | 11 | -0.70 | 0.59 | 2.69E-01 | 0.50 | 0.16 | 1.59 | MR Egger | Univariable |
| BMI in nulliparous women between 30 and <40 years | -- | HER2-enriched subtype | 49.2 | 5 | -0.46 | 0.25 | 6.32E-02 | 0.63 | 0.39 | 1.03 | IVW | Univariable |
| BMI in nulliparous women between 30 and <40 years | -- | HER2-enriched subtype | 49.2 | 5 | -0.44 | 0.22 | 4.26E-02 | 0.65 | 0.42 | 0.99 | Weighted median | Univariable |

| | | | | | | | | | | | | |
|---|---|---|---|---|---|---|---|---|---|---|---|---|
| BMI in nulliparous women between 30 and <40 years | -- | HER2-enriched subtype | 49.2 | 5 | 0.28 | 1.04 | 8.07E-01 | 1.32 | 0.17 | 10.08 | MR Egger | Univariable |
| BMI in nulliparous women between menarche and <40 years | Prepubertal body size | HER2-enriched subtype | 2.9 | 16 | -0.04 | 0.16 | 8.14E-01 | 0.96 | 0.71 | 1.31 | IVW | Multivariable |
| Prepubertal body size | BMI in nulliparous women between menarche and <40 years | HER2-enriched subtype | 3.6 | 79 | -0.24 | 0.20 | 2.37E-01 | 0.79 | 0.53 | 1.17 | IVW | Multivariable |
| BMI in nulliparous women between menarche and <40 years | Adult body size | HER2-enriched subtype | 3.4 | 18 | -0.26 | 0.15 | 7.43E-02 | 0.77 | 0.57 | 1.03 | IVW | Multivariable |
| Adult body size | BMI in nulliparous women between menarche and <40 years | HER2-enriched subtype | 4.8 | 115 | 0.19 | 0.19 | 3.22E-01 | 1.21 | 0.83 | 1.77 | IVW | Multivariable |
| BMI in nulliparous women between menarche and <20 years | Prepubertal body size | HER2-enriched subtype | 1.2 | 3 | 0.01 | 0.14 | 9.41E-01 | 1.01 | 0.77 | 1.32 | IVW | Multivariable |
| Prepubertal body size | BMI in nulliparous women between menarche and <20 years | HER2-enriched subtype | 1.7 | 73 | -0.36 | 0.20 | 7.16E-02 | 0.70 | 0.47 | 1.03 | IVW | Multivariable |
| BMI in nulliparous women between menarche and <20 years | Adult body size | HER2-enriched subtype | 1.7 | 3 | -0.25 | 0.12 | 3.94E-02 | 0.78 | 0.61 | 0.99 | IVW | Multivariable |
| Adult body size | BMI in nulliparous women between menarche and <20 years | HER2-enriched subtype | 3.2 | 114 | 0.10 | 0.16 | 5.30E-01 | 1.11 | 0.80 | 1.53 | IVW | Multivariable |
| BMI in nulliparous women between <20 years and 30 | Prepubertal body size | HER2-enriched subtype | 2.1 | 10 | 0.03 | 0.16 | 8.57E-01 | 1.03 | 0.75 | 1.42 | IVW | Multivariable |
| Prepubertal body size | BMI in nulliparous women between 20 and <30 years | HER2-enriched subtype | 2.8 | 77 | -0.33 | 0.21 | 1.27E-01 | 0.72 | 0.48 | 1.10 | IVW | Multivariable |
| BMI in nulliparous women between <20 years and 30 | Adult body size | HER2-enriched subtype | 2.1 | 10 | -0.18 | 0.16 | 2.72E-01 | 0.84 | 0.61 | 1.15 | IVW | Multivariable |
| Adult body size | BMI in nulliparous women between 20 and <30 years | HER2-enriched subtype | 3.2 | 116 | 0.07 | 0.21 | 7.24E-01 | 1.08 | 0.72 | 1.62 | IVW | Multivariable |
| BMI in nulliparous women between 30 and <40 years | Prepubertal body size | HER2-enriched subtype | 1.7 | 3 | -0.04 | 0.12 | 7.64E-01 | 0.96 | 0.76 | 1.22 | IVW | Multivariable |
| Prepubertal body size | BMI in nulliparous women between 30 and <40 years | HER2-enriched subtype | 3.2 | 75 | -0.28 | 0.18 | 1.24E-01 | 0.76 | 0.53 | 1.08 | IVW | Multivariable |
| BMI in nulliparous women between 30 and <40 years | Adult body size | HER2-enriched subtype | 1.8 | 3 | -0.23 | 0.12 | 6.54E-02 | 0.80 | 0.63 | 1.01 | IVW | Multivariable |
| Adult body size | BMI in nulliparous women between 30 and <40 years | HER2-enriched subtype | 4.3 | 114 | 0.10 | 0.17 | 5.33E-01 | 1.11 | 0.80 | 1.54 | IVW | Multivariable |
| BMI in nulliparous women between menarche and <40 years | -- | Triple-negative subtype | 63.7 | 22 | -0.34 | 0.10 | 8.31E-04 | 0.71 | 0.59 | 0.87 | IVW | Univariable |
| BMI in nulliparous women between menarche and <40 years | -- | Triple-negative subtype | 63.7 | 22 | -0.36 | 0.11 | 1.44E-03 | 0.70 | 0.56 | 0.87 | Weighted median | Univariable |
| BMI in nulliparous women between menarche and <40 years | -- | Triple-negative subtype | 63.7 | 22 | -0.83 | 0.24 | 2.55E-03 | 0.44 | 0.27 | 0.70 | MR Egger | Univariable |
| BMI in nulliparous women between menarche and <20 years | -- | Triple-negative subtype | 48.4 | 3 | -0.56 | 0.21 | 8.90E-03 | 0.57 | 0.38 | 0.87 | IVW | Univariable |
| BMI in nulliparous women between menarche and <20 years | -- | Triple-negative subtype | 48.4 | 3 | -0.62 | 0.15 | 1.90E-05 | 0.54 | 0.40 | 0.71 | Weighted median | Univariable |

| | | | | | | | | | | | | |
|---|---|---|---|---|---|---|---|---|---|---|---|---|
| BMI in nulliparous women between menarche and <20 years | -- | Triple-negative subtype | 48.4 | 3 | -1.57 | 2.64 | 6.59E-01 | 0.21 | 0.00 | 36.90 | MR Egger | Univariable |
| BMI in nulliparous women between 20 and <30 years | -- | Triple-negative subtype | 59.2 | 11 | -0.41 | 0.11 | 1.51E-04 | 0.66 | 0.54 | 0.82 | IVW | Univariable |
| BMI in nulliparous women between 20 and <30 years | -- | Triple-negative subtype | 59.2 | 11 | -0.34 | 0.11 | 2.93E-03 | 0.71 | 0.57 | 0.89 | Weighted median | Univariable |
| BMI in nulliparous women between 20 and <30 years | -- | Triple-negative subtype | 59.2 | 11 | -0.46 | 0.38 | 2.55E-01 | 0.63 | 0.30 | 1.32 | MR Egger | Univariable |
| BMI in nulliparous women between 30 and <40 years | -- | Triple-negative subtype | 49.2 | 5 | -0.40 | 0.18 | 2.55E-02 | 0.67 | 0.47 | 0.95 | IVW | Univariable |
| BMI in nulliparous women between 30 and <40 years | -- | Triple-negative subtype | 49.2 | 5 | -0.21 | 0.14 | 1.32E-01 | 0.81 | 0.62 | 1.07 | Weighted median | Univariable |
| BMI in nulliparous women between 30 and <40 years | -- | Triple-negative subtype | 49.2 | 5 | -0.08 | 0.79 | 9.21E-01 | 0.92 | 0.20 | 4.31 | MR Egger | Univariable |
| BMI in nulliparous women between menarche and <40 years | Prepubertal body size | Triple-negative subtype | 2.9 | 16 | -0.14 | 0.10 | 1.66E-01 | 0.87 | 0.71 | 1.06 | IVW | Multivariable |
| Prepubertal body size | BMI in nulliparous women between menarche and <40 years | Triple-negative subtype | 3.6 | 79 | -0.20 | 0.14 | 1.48E-01 | 0.82 | 0.63 | 1.07 | IVW | Multivariable |
| BMI in nulliparous women between menarche and <40 years | Adult body size | Triple-negative subtype | 3.4 | 18 | -0.31 | 0.10 | 2.45E-03 | 0.73 | 0.60 | 0.90 | IVW | Multivariable |
| Adult body size | BMI in nulliparous women between menarche and <40 years | Triple-negative subtype | 4.8 | 115 | 0.06 | 0.14 | 6.38E-01 | 1.07 | 0.82 | 1.39 | IVW | Multivariable |
| BMI in nulliparous women between menarche and <20 years | Prepubertal body size | Triple-negative subtype | 1.2 | 3 | 0.02 | 0.10 | 8.34E-01 | 1.02 | 0.84 | 1.23 | IVW | Multivariable |
| Prepubertal body size | BMI in nulliparous women between menarche and <20 years | Triple-negative subtype | 1.7 | 73 | -0.35 | 0.14 | 1.17E-02 | 0.70 | 0.53 | 0.92 | IVW | Multivariable |
| BMI in nulliparous women between menarche and <20 years | Adult body size | Triple-negative subtype | 1.7 | 3 | -0.17 | 0.09 | 4.86E-02 | 0.84 | 0.71 | 1.00 | IVW | Multivariable |
| Adult body size | BMI in nulliparous women between menarche and <20 years | Triple-negative subtype | 3.2 | 114 | -0.09 | 0.12 | 4.23E-01 | 0.91 | 0.72 | 1.15 | IVW | Multivariable |
| BMI in nulliparous women between 20 and <30 years | Prepubertal body size | Triple-negative subtype | 2.1 | 10 | -0.27 | 0.11 | 1.45E-02 | 0.76 | 0.61 | 0.95 | IVW | Multivariable |
| Prepubertal body size | BMI in nulliparous women between 20 and <30 years | Triple-negative subtype | 2.8 | 77 | -0.07 | 0.14 | 6.41E-01 | 0.94 | 0.71 | 1.24 | IVW | Multivariable |
| BMI in nulliparous women between 20 and <30 years | Adult body size | Triple-negative subtype | 2.1 | 10 | -0.36 | 0.11 | 1.32E-03 | 0.70 | 0.56 | 0.87 | IVW | Multivariable |
| Adult body size | BMI in nulliparous women between 20 and <30 years | Triple-negative subtype | 3.2 | 116 | 0.11 | 0.15 | 4.53E-01 | 1.12 | 0.84 | 1.48 | IVW | Multivariable |
| BMI in nulliparous women between 30 and <40 years | Prepubertal body size | Triple-negative subtype | 1.7 | 3 | -0.01 | 0.08 | 8.66E-01 | 0.99 | 0.84 | 1.16 | IVW | Multivariable |
| Prepubertal body size | BMI in nulliparous women between 30 and <40 years | Triple-negative subtype | 3.2 | 75 | -0.32 | 0.12 | 8.91E-03 | 0.72 | 0.57 | 0.92 | IVW | Multivariable |
| BMI in nulliparous women between 30 and <40 years | Adult body size | Triple-negative subtype | 1.8 | 3 | -0.15 | 0.09 | 1.00E-01 | 0.86 | 0.72 | 1.03 | IVW | Multivariable |
| BMI in nulliparous women between menarche and <40 years | -- | Triple-negative subtype | 4.3 | 114 | -0.12 | 0.12 | 3.16E-01 | 0.89 | 0.70 | 1.12 | IVW | Multivariable |

*Table S6. Univariable Mendelian randomization analyses parity onto BMI in nulliparous women between menarche and <40 years and overall breast cancer and subtypes.*

nSNP - number of single nucleotide polymorphism identifiers, Beta - effect estimate coefficient for SNP on BMI, SE- standard error of the effect estimate, LCI - lower confidence interval, UCI - upper confidence interval, P - corresponding p-value, IVW - inverse variance weighted

| Exposure | Outcome | nSNP | beta | SE | P | Odds ratio | LCI | UCI | Method |
|---|---|---|---|---|---|---|---|---|---|
| Number of live births | BMI in nulliparous women between menarche and <40 years | 10 | -0.01 | 0.03 | 0.58 | 0.99 | 0.94 | 1.04 | IVW |
| Number of live births | BMI in nulliparous women between menarche and <40 years | 10 | -0.02 | 0.02 | 0.36 | 0.98 | 0.94 | 1.02 | Weighted median |
| Number of live births | BMI in nulliparous women between menarche and <40 years | 10 | 0.21 | 0.24 | 0.41 | 1.23 | 0.77 | 1.95 | MR Egger |
| Number of live births | BMI in nulliparous women between menarche and <20 years | 10 | 0.01 | 0.04 | 0.75 | 1.01 | 0.94 | 1.09 | IVW |
| Number of live births | BMI in nulliparous women between menarche and <20 years | 10 | -0.03 | 0.04 | 0.52 | 0.97 | 0.90 | 1.06 | Weighted median |
| Number of live births | BMI in nulliparous women between menarche and <20 years | 10 | -0.27 | 0.34 | 0.45 | 0.76 | 0.39 | 1.48 | MR Egger |
| Number of live births | BMI in nulliparous women between 20 and <30 years | 10 | 0.00 | 0.03 | 0.94 | 1.00 | 0.94 | 1.06 | IVW |
| Number of live births | BMI in nulliparous women between 20 and <30 years | 10 | -0.01 | 0.03 | 0.63 | 0.99 | 0.93 | 1.05 | Weighted median |
| Number of live births | BMI in nulliparous women between 20 and <30 years | 10 | 0.30 | 0.29 | 0.32 | 1.35 | 0.77 | 2.37 | MR Egger |
| Number of live births | BMI in nulliparous women between 30 and <40 years | 10 | -0.07 | 0.03 | 0.05 | 0.93 | 0.87 | 1.00 | IVW |
| Number of live births | BMI in nulliparous women between 30 and <40 years | 10 | -0.07 | 0.04 | 0.07 | 0.94 | 0.87 | 1.00 | Weighted median |
| Number of live births | BMI in nulliparous women between 30 and <40 years | 10 | 0.08 | 0.33 | 0.81 | 1.09 | 0.57 | 2.07 | MR Egger |

*Table S7. Multivariable Mendelian randomization analyses for BMI in nulliparous women between menarche and <40 years onto overall breast cancer accounting for age at menarche.*

nSNP - number of single nucleotide polymorphism identifiers, Beta - effect estimate coefficient for SNP on BMI, SE- standard error of the effect estimate, LCI - lower confidence interval, UCI - upper confidence interval, P - corresponding p-value, IVW - inverse variance weighted, MR - Mendelian randomization

| Exposure | Accounting for | Outcome | nSNP | beta | SE | P | Odds ratio | LCI | UCI | Method | MR |
|---|---|---|---|---|---|---|---|---|---|---|---|
| BMI in nulliparous women between menarche and <40 years | Age at menarche | Overall breast cancer | 13 | -0.32 | 0.05 | 4.04E-11 | 0.72 | 0.66 | 0.79 | IVW | Multivariable |
| Age at menarche | BMI in nulliparous women between menarche and <40 years | Overall breast cancer | 70 | -0.05 | 0.03 | 1.37E-01 | 0.95 | 0.90 | 1.01 | IVW | Multivariable |
| BMI in nulliparous women between menarche and <20 years | Age at menarche | Overall breast cancer | 3 | -0.31 | 0.05 | 4.56E-09 | 0.74 | 0.67 | 0.81 | IVW | Multivariable |
| Age at menarche | BMI in nulliparous women between menarche and <20 years | Overall breast cancer | 68 | -0.05 | 0.03 | 1.35E-01 | 0.95 | 0.90 | 1.01 | IVW | Multivariable |
| BMI in nulliparous women between 20 and <30 years | Age at menarche | Overall breast cancer | 7 | -0.25 | 0.06 | 2.10E-05 | 0.78 | 0.70 | 0.87 | IVW | Multivariable |
| Age at menarche | BMI in nulliparous women between 20 and <30 years | Overall breast cancer | 69 | -0.04 | 0.03 | 2.08E-01 | 0.96 | 0.91 | 1.02 | IVW | Multivariable |
| BMI in nulliparous women between 30 and <40 years | Age at menarche | Overall breast cancer | 5 | -0.28 | 0.06 | 1.14E-06 | 0.76 | 0.68 | 0.85 | IVW | Multivariable |
| Age at menarche | BMI in nulliparous women between menarche and <40 years | Overall breast cancer | 69 | -0.05 | 0.03 | 1.56E-01 | 0.95 | 0.90 | 1.01 | IVW | Multivariable |
| BMI in nulliparous women between menarche and <40 years | Age at menarche | ER+ breast cancer | 13 | -0.33 | 0.06 | 2.24E-10 | 0.72 | 0.64 | 0.80 | IVW | Multivariable |
| Age at menarche | BMI in nulliparous women between menarche and <40 years | ER+ breast cancer | 70 | -0.06 | 0.03 | 6.47E-02 | 0.94 | 0.89 | 1.00 | IVW | Multivariable |
| BMI in nulliparous women between menarche and <20 years | Age at menarche | ER+ breast cancer | 3 | -0.32 | 0.05 | 8.32E-09 | 0.73 | 0.66 | 0.81 | IVW | Multivariable |
| Age at menarche | BMI in nulliparous women between menarche and <20 years | ER+ breast cancer | 68 | -0.07 | 0.03 | 5.33E-02 | 0.93 | 0.88 | 0.99 | IVW | Multivariable |
| BMI in nulliparous women between 20 and <30 years | Age at menarche | ER+ breast cancer | 7 | -0.27 | 0.06 | 6.14E-06 | 0.76 | 0.68 | 0.85 | IVW | Multivariable |
| Age at menarche | BMI in nulliparous women between 20 and <30 years | ER+ breast cancer | 69 | -0.06 | 0.04 | 9.07E-02 | 0.94 | 0.87 | 1.02 | IVW | Multivariable |
| BMI in nulliparous women between 30 and <40 years | Age at menarche | ER+ breast cancer | 5 | -0.28 | 0.06 | 3.72E-06 | 0.76 | 0.68 | 0.85 | IVW | Multivariable |
| Age at menarche | BMI in nulliparous women between menarche and <40 years | ER+ breast cancer | 69 | -0.06 | 0.04 | 7.58E-02 | 0.94 | 0.87 | 1.02 | IVW | Multivariable |
| BMI in nulliparous women between menarche and <40 years | Age at menarche | ER- breast cancer | 13 | -0.37 | 0.08 | 2.97E-06 | 0.69 | 0.59 | 0.80 | IVW | Multivariable |
| Age at menarche | BMI in nulliparous women between menarche and <40 years | ER- breast cancer | 70 | 0.00 | 0.05 | 9.34E-01 | 1.00 | 0.91 | 1.10 | IVW | Multivariable |

| | | | | | | | | | | | |
|---|---|---|---|---|---|---|---|---|---|---|---|
| BMI in nulliparous women between menarche and <20 years | Age at menarche | ER- breast cancer | 3 | -0.36 | 0.08 | 3.35E-05 | 0.70 | 0.60 | 0.82 | IVW | Multivariable |
| Age at menarche | BMI in nulliparous women between menarche and <20 years | ER- breast cancer | 68 | 0.01 | 0.05 | 9.07E-01 | 1.01 | 0.92 | 1.11 | IVW | Multivariable |
| BMI in nulliparous women between 20 and <30 years | Age at menarche | ER- breast cancer | 7 | -0.33 | 0.09 | 2.41E-04 | 0.72 | 0.60 | 0.86 | IVW | Multivariable |
| Age at menarche | BMI in nulliparous women between 20 and <30 years | ER- breast cancer | 69 | 0.01 | 0.05 | 9.20E-01 | 1.01 | 0.92 | 1.11 | IVW | Multivariable |
| BMI in nulliparous women between 30 and <40 years | Age at menarche | ER- breast cancer | 5 | -0.30 | 0.10 | 1.45E-03 | 0.74 | 0.61 | 0.89 | IVW | Multivariable |
| Age at menarche | BMI in nulliparous women between menarche and <40 years | ER- breast cancer | 69 | 0.02 | 0.06 | 7.81E-01 | 1.02 | 0.91 | 1.15 | IVW | Multivariable |
| BMI in nulliparous women between menarche and <40 years | Age at menarche | Luminal A subtype | 13 | -0.29 | 0.06 | 1.77E-07 | 0.75 | 0.67 | 0.83 | IVW | Multivariable |
| Age at menarche | BMI in nulliparous women between menarche and <40 years | Luminal A subtype | 70 | -0.05 | 0.04 | 1.20E-01 | 0.95 | 0.88 | 1.03 | IVW | Multivariable |
| BMI in nulliparous women between menarche and <20 years | Age at menarche | Luminal A subtype | 3 | -0.29 | 0.06 | 1.08E-06 | 0.75 | 0.67 | 0.85 | IVW | Multivariable |
| Age at menarche | BMI in nulliparous women between menarche and <20 years | Luminal A subtype | 68 | -0.06 | 0.04 | 1.02E-01 | 0.94 | 0.87 | 1.02 | IVW | Multivariable |
| BMI in nulliparous women between 20 and <30 years | Age at menarche | Luminal A subtype | 7 | -0.24 | 0.07 | 2.19E-04 | 0.79 | 0.69 | 0.90 | IVW | Multivariable |
| Age at menarche | BMI in nulliparous women between 20 and <30 years | Luminal A subtype | 69 | -0.06 | 0.04 | 1.25E-01 | 0.94 | 0.87 | 1.02 | IVW | Multivariable |
| BMI in nulliparous women between 30 and <40 years | Age at menarche | Luminal A subtype | 5 | -0.24 | 0.06 | 1.46E-04 | 0.79 | 0.69 | 0.89 | IVW | Multivariable |
| Age at menarche | BMI in nulliparous women between 30 and <40 years | Luminal A subtype | 69 | -0.06 | 0.04 | 1.35E-01 | 0.94 | 0.87 | 1.02 | IVW | Multivariable |
| BMI in nulliparous women between menarche and <40 years | Age at menarche | Luminal B1 subtype | 13 | -0.40 | 0.09 | 2.84E-06 | 0.67 | 0.56 | 0.79 | IVW | Multivariable |
| Age at menarche | BMI in nulliparous women between menarche and <40 years | Luminal B1 subtype | 70 | -0.06 | 0.05 | 2.70E-01 | 0.94 | 0.85 | 1.04 | IVW | Multivariable |
| BMI in nulliparous women between menarche and <20 years | Age at menarche | Luminal B1 subtype | 3 | -0.40 | 0.09 | 6.38E-06 | 0.67 | 0.56 | 0.80 | IVW | Multivariable |
| Age at menarche | BMI in nulliparous women between menarche and <20 years | Luminal B1 subtype | 68 | -0.06 | 0.05 | 2.54E-01 | 0.94 | 0.85 | 1.04 | IVW | Multivariable |
| BMI in nulliparous women between 20 and <30 years | Age at menarche | Luminal B1 subtype | 7 | -0.32 | 0.09 | 1.13E-03 | 0.73 | 0.60 | 0.87 | IVW | Multivariable |
| Age at menarche | BMI in nulliparous women between 20 and <30 years | Luminal B1 subtype | 69 | -0.05 | 0.06 | 4.16E-01 | 0.95 | 0.85 | 1.07 | IVW | Multivariable |
| BMI in nulliparous women between 30 and <40 years | Age at menarche | Luminal B1 subtype | 5 | -0.28 | 0.10 | 4.30E-03 | 0.76 | 0.63 | 0.92 | IVW | Multivariable |
| Age at menarche | BMI in nulliparous women between 30 and <40 years | Luminal B1 subtype | 69 | -0.05 | 0.06 | 3.57E-01 | 0.95 | 0.85 | 1.07 | IVW | Multivariable |

| | | | | | | | | | | | |
|---|---|---|---|---|---|---|---|---|---|---|---|
| BMI in nulliparous women between menarche and <40 years | Age at menarche | Luminal B2 subtype | 13 | -0.32 | 0.08 | 4.64E-05 | 0.73 | 0.62 | 0.85 | IVW | Multivariable |
| Age at menarche | BMI in nulliparous women between menarche and <40 years | Luminal B2 subtype | 70 | -0.09 | 0.05 | 5.86E-02 | 0.91 | 0.83 | 1.01 | IVW | Multivariable |
| BMI in nulliparous women between menarche and <20 years | Age at menarche | Luminal B2 subtype | 3 | -0.34 | 0.08 | 3.26E-05 | 0.71 | 0.61 | 0.84 | IVW | Multivariable |
| Age at menarche | BMI in nulliparous women between menarche and <20 years | Luminal B2 subtype | 68 | -0.10 | 0.05 | 4.58E-02 | 0.90 | 0.82 | 1.00 | IVW | Multivariable |
| BMI in nulliparous women between 20 and <30 years | Age at menarche | Luminal B2 subtype | 7 | -0.26 | 0.09 | 3.94E-03 | 0.77 | 0.64 | 0.92 | IVW | Multivariable |
| Age at menarche | BMI in nulliparous women between 20 and <30 years | Luminal B2 subtype | 69 | -0.09 | 0.05 | 8.97E-02 | 0.91 | 0.83 | 1.01 | IVW | Multivariable |
| BMI in nulliparous women between 30 and <40 years | Age at menarche | Luminal B2 subtype | 5 | -0.36 | 0.08 | 2.95E-05 | 0.70 | 0.59 | 0.82 | IVW | Multivariable |
| Age at menarche | BMI in nulliparous women between 30 and <40 years | Luminal B2 subtype | 69 | -0.11 | 0.05 | 3.58E-02 | 0.90 | 0.81 | 0.99 | IVW | Multivariable |
| BMI in nulliparous women between menarche and <40 years | Age at menarche | HER2-enriched subtype | 13 | -0.19 | 0.13 | 1.24E-01 | 0.83 | 0.64 | 1.06 | IVW | Multivariable |
| Age at menarche | BMI in nulliparous women between menarche and <40 years | HER2-enriched subtype | 70 | 0.11 | 0.08 | 1.63E-01 | 1.12 | 0.95 | 1.31 | IVW | Multivariable |
| BMI in nulliparous women between menarche and <20 years | Age at menarche | HER2-enriched subtype | 3 | -0.27 | 0.13 | 3.50E-02 | 0.76 | 0.59 | 0.98 | IVW | Multivariable |
| Age at menarche | BMI in nulliparous women between menarche and <20 years | HER2-enriched subtype | 68 | 0.11 | 0.08 | 1.98E-01 | 1.12 | 0.95 | 1.31 | IVW | Multivariable |
| BMI in nulliparous women between 20 and <30 years | Age at menarche | HER2-enriched subtype | 7 | -0.16 | 0.14 | 2.65E-01 | 0.85 | 0.65 | 1.12 | IVW | Multivariable |
| Age at menarche | BMI in nulliparous women between 20 and <30 years | HER2-enriched subtype | 69 | 0.11 | 0.08 | 1.91E-01 | 1.12 | 0.95 | 1.31 | IVW | Multivariable |
| BMI in nulliparous women between 30 and <40 years | Age at menarche | HER2-enriched subtype | 5 | -0.28 | 0.14 | 4.52E-02 | 0.75 | 0.57 | 0.99 | IVW | Multivariable |
| Age at menarche | BMI in nulliparous women between 30 and <40 years | HER2-enriched subtype | 69 | 0.11 | 0.08 | 1.77E-01 | 1.12 | 0.95 | 1.31 | IVW | Multivariable |
| BMI in nulliparous women between menarche and <40 years | Age at menarche | Triple-negative subtype | 13 | -0.31 | 0.08 | 2.00E-04 | 0.73 | 0.63 | 0.86 | IVW | Multivariable |
| Age at menarche | BMI in nulliparous women between menarche and <40 years | Triple-negative subtype | 70 | -0.03 | 0.05 | 5.94E-01 | 0.97 | 0.88 | 1.07 | IVW | Multivariable |
| BMI in nulliparous women between menarche and <20 years | Age at menarche | Triple-negative subtype | 3 | -0.31 | 0.09 | 5.09E-04 | 0.74 | 0.62 | 0.88 | IVW | Multivariable |
| Age at menarche | BMI in nulliparous women between menarche and <20 years | Triple-negative subtype | 68 | -0.03 | 0.05 | 6.10E-01 | 0.97 | 0.88 | 1.07 | IVW | Multivariable |
| BMI in nulliparous women between 20 and <30 years | Age at menarche | Triple-negative subtype | 7 | -0.28 | 0.09 | 1.70E-03 | 0.75 | 0.63 | 0.91 | IVW | Multivariable |
| Age at menarche | BMI in nulliparous women between 20 and <30 years | Triple-negative subtype | 69 | -0.03 | 0.05 | 5.96E-01 | 0.97 | 0.88 | 1.07 | IVW | Multivariable |

| | | | | | | | | | | | |
|---|---|---|---|---|---|---|---|---|---|---|---|
| BMI in nulliparous women between 30 and <40 years | Age at menarche | Triple-negative subtype | 5 | -0.24 | 0.09 | 7.28E-03 | 0.78 | 0.66 | 0.93 | IVW | Multivariable |
| Age at menarche | BMI in nulliparous women between 30 and <40 years | Triple-negative subtype | 69 | -0.02 | 0.05 | 6.74E-01 | 0.98 | 0.89 | 1.08 | IVW | Multivariable |

References.

39. Prince C, Howe LD, Sharp GC, Fraser A, Richmond RC. Establishing the relationships between adiposity and reproductive factors: a multivariable Mendelian randomization analysis. BMC Med. 2023;21(1):350.
40. Willer CJ, Li Y, Abecasis GR. METAL: fast and efficient meta-analysis of genomewide association scans. Bioinformatics. 2010;26(17):2190-1.
41. Abecasis GR, Auton A, Brooks LD, DePristo MA, Durbin RM, Handsaker RE, et al. An integrated map of genetic variation from 1,092 human genomes. Nature. 2012;491(7422):56-65.
42. R Core Team. R: A language and environment for statistical computing. Vienna, Austria: R Foundation for Statistical Computing; 2020.
43. Vabistsevits M, Davey Smith G, Richardson TG, Richmond RC, Sieh W, Rothstein JH, et al. Mammographic density mediates the protective effect of early-life body size on breast cancer risk. Nat Commun. 2024;15(1):4021.
44. Tran TXM, Chang Y, Choi HR, Kwon R, Lim G-Y, Kim EY, et al. Adiposity, Body Composition Measures, and Breast Cancer Risk in Korean Premenopausal Women. JAMA Network Open. 2024;7(4):e245423-e.
45. van den Brandt PA, Spiegelman D, Yaun SS, Adami HO, Beeson L, Folsom AR, et al. Pooled analysis of prospective cohort studies on height, weight, and breast cancer risk. Am J Epidemiol. 2000;152(6):514-27.
46. van den Brandt PA, Ziegler RG, Wang M, Hou T, Li R, Adami HO, et al. Body size and weight change over adulthood and risk of breast cancer by menopausal and hormone receptor status: a pooled analysis of 20 prospective cohort studies. Eur J Epidemiol. 2021;36(1):37-55.
47. Dehesh T, Fadaghi S, Seyedi M, Abolhadi E, Ilaghi M, Shams P, et al. The relation between obesity and breast cancer risk in women by considering menstruation status and geographical variations: a systematic review and meta-analysis. BMC Women's Health. 2023;23(1):392.
48. Friedenreich CM. Review of anthropometric factors and breast cancer risk. Eur J Cancer Prev. 2001;10(1):15-32.
49. Neuhouser ML, Aragaki AK, Prentice RL, Manson JE, Chlebowski R, Carty CL, et al. Overweight, Obesity, and Postmenopausal Invasive Breast Cancer Risk: A Secondary Analysis of the Women's Health Initiative Randomized Clinical Trials. JAMA Oncol. 2015;1(5):611-21.
50. Glassman I, Le N, Asif A, Goulding A, Alcantara CA, Vu A, et al. The Role of Obesity in Breast Cancer Pathogenesis. Cells. 2023;12(16).
51. Cecchini RS, Costantino JP, Cauley JA, Cronin WM, Wickerham DL, Land SR, et al. Body mass index and the risk for developing invasive breast cancer among high-risk women in NSABP P-1 and STAR breast cancer prevention trials. Cancer Prev Res (Phila). 2012;5(4):583-92.
52. Zhao P, Xia N, Zhang H, Deng T. The Metabolic Syndrome Is a Risk Factor for Breast Cancer: A Systematic Review and Meta-Analysis. Obes Facts. 2020;13(4):384-96.
